# Association between national action and trends in antibiotic resistance: an analysis of 73 countries from 2000 to 2023

**DOI:** 10.1101/2024.09.25.24313966

**Authors:** Peter Søgaard Jørgensen, Luong Nguyen Thanh, Ege Pehlivanoglu, Franziska Klein, Didier Wernli, Dusan Jasovsky, Athena Aktipis, Robert R. Dunn, Yrjo Gröhn, Guillaume Lhermie, H. Morgan Scott, Eili Y. Klein

**Affiliations:** Global Economic Dynamics and the Biosphere, The Royal Swedish Academy of Sciences, Stockholm, Sweden; Stockholm Resilience Centre, Stockholm University, Stockholm, Sweden; Uppsala Antibiotic Centre and Department of Women’s and Children’s Health, Uppsala University, Uppsala, Sweden; University of Geneva, Global Studies Institute, Transformative Governance Lab, Geneva, Switzerland; Uppsala University, ReAct Europe, Uppsala, Sweden; Arizona State University, AZ, USA; North Carolina State University, Raleigh, NC, USA; Cornell University, Ithaca, NY, USA; Texas A&M University, College Station, TX, USA; One Health Trust, Washington, D.C. 20015, USA; Hopkins School of Medicine, Department of Emergency Medicine, Baltimore, MD, USA

**Keywords:** TrACSS, national policy, antibiotic use, DRI, DPSEEA

## Abstract

**Background:** The world’s governments have agreed both global and national actions to address the challenge of antimicrobial resistance. This raises the importance of understanding to what degree national action so far has been effective. Answering this question is challenged by variation in data availability and quality as well as disruptive events such as the COVID-19 pandemic. We investigate the association between a survey of self-reported action based on the first Global Database for Tracking Antimicrobial Resistance (TrACSS) survey and trends in multiple indicators related to the DPSEEA framework leading up to the survey.

**Methods and findings:** We apply regression methods across 73 countries between actions in 2016 and the trend in indicators of health system development (drivers), antibiotic use (pressures, ABU), absolute rates of resistance (state, ABR) and relative rates of resistance (exposure, Drug Resistance Index, DRI) from 2000 to 2016. We find that action is consistently associated with improved linear and categorical trend in health systems, ABU, ABR and DRI. Reductions are associated with relatively high levels of action (0-4) for ABU (median 2.8, 25-75% quartile 2.6-3.3), ABR (3.0, 2.4-3.4), and DRI (3.5, 3.1-3.6). These associations are robust to the inclusion of other contextual factors such as health system and socio-economic status, human population density, animal production and climate. Since 2016, a majority of both Low-Middle Income Countries (LMICs) and High-Income Countries (HICs) report increased action on repeated questions, while one third of countries report reduced action. The main limitations in interpretation are heterogeneity in data availability and the recency of action.

**Conclusions:** Our findings highlight the importance of national action to address the domestic situation related to antibiotic resistance and indicate the value of both incremental changes in reducing adverse outcomes and the need for high levels of action in delivering improvements.

## Introduction

Antibiotic resistance (ABR) is a global public health challenge that in 2019 was estimated to contribute to around 1.27 million deaths per year and associated with a total of 4.95 million deaths [1]. In 2015 countries agreed a global action plan (GAP) to address the growing challenge of ABR, which was followed up in 2016 at a UN high-level meeting with commitments to develop national action plans (NAPs). In 2024 countries are meeting again at another high-level meeting of the UN to agree the next steps for global and national governance. New goals currently in discussion include the 10-20-30 by 2030 goal to reduce mortality by 10 %, inappropriate human antibiotic use (ABU) by 20 % and inappropriate animal ABU by 30 % by 2030 [2].

Given the severe burden of ABR and the increasing focus on national action, it is important to understand what association there is between action taken so far and improvements in national conditions. The fact that ABR can be limited through multiple points of intervention, including by addressing upstream drivers of ABU complicates assessment [3,4]. Variation in data availability across countries and the recent disruption of the COVID-19 pandemic further adds to these challenges [5]. Instead of associating action taken since the agreement of the GAP with recent trends in ABR, another approach is to see whether countries that were already taking action when the GAP was agreed experienced improved trends in key indicators. Such information will help provide a conservative baseline for the impact of national action on ABR going forwards.

Here we apply a multi-indicator approach inspired by the DPSEEA family of frameworks to assess the association between action reported in the TrACSS survey in 2016 and preceding national trends in indicators related to drivers (health system), pressures (ABU), state (ABR) and exposure to antibiotic resistant infections in humans (Drug Resistance Index, DRI, Fig 1). We apply cross-sectional regression across 73 countries (S1 Table) and model selection to assess the strength of association with linear as well as categorical trends. Finally, we provide assessment of the extent to which countries have increased action from 2016 up until 2023 and assess the prevalence of positively and negatively reinforcing governance responses (Fig 1B).

**Fig 1.**
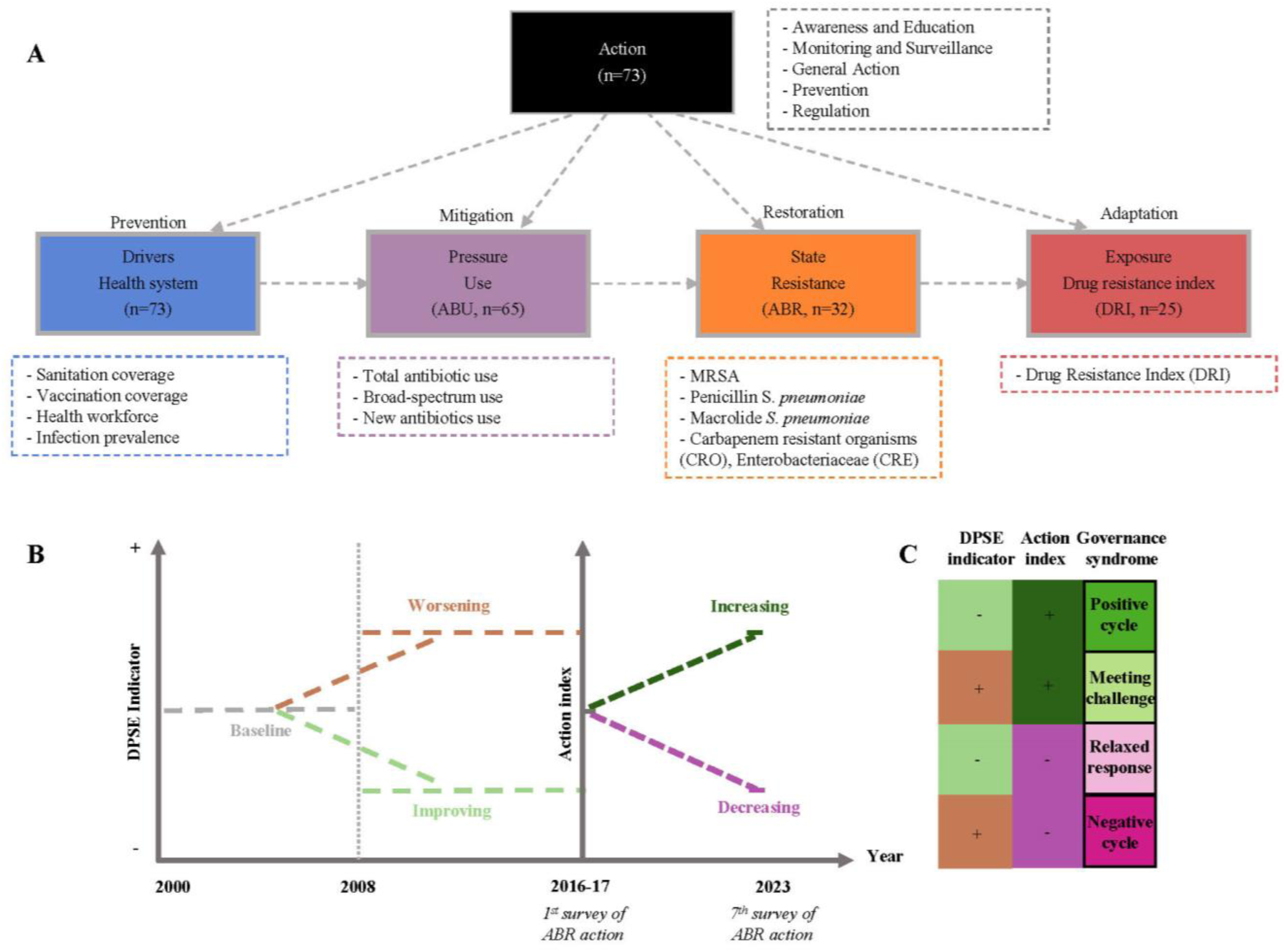
Study design. (A) DPSE indicators (tier 1) and their tier 2 components (boxes with dashed-lines). “n” refers to number of countries, for detailed information on indicators see S2 Table. (B) Temporal overview of analysis showing the two periods across which the trend in DPSE indicators is calculated and the timing of the first (2016-17) and seventh (2023) TrACSS survey. (C) Governance syndrome. Categorical trends in DPSE indicators and the action index combine to form four types of governance syndrome.

## Methods

### Study design

We investigate national trends in indicators related to antibiotic resistance (ABR) by using a multi-indicator assessment with indicators organized according to a modified version of the DPSEEA (Drivers-Pressure-State-Exposure-Effect-Action) family of frameworks [6] (Fig 1, S2 Table). Such frameworks aim to assess how policies affects changes at multiple points in a system [7] and similar frameworks, have been used to evaluate national progress on health and biodiversity issues [8], but also to understand the co-evolutionary dynamics of pesticide resistance [9].

Our main purpose is to understand to what degree self-reported policy action (Action) can help explain temporal trends in national indicators of drivers, pressures, state and exposure (DPSE) indicators (Fig 1A). The DPSE categories capture indicators of health system development (Drivers), antibiotic use (Pressures), antibiotic resistance (State), and antibiotic resistance relative to use of an antibiotic as measured by the Drug Resistance Index (DRI, Exposure, *see Indicators and Data sources for details*). To construct the action index, we use the Global Database for TrACSS survey first conducted in 2016-17 (Fig 1B). As 2016 was the first survey since the agreement of the global action plan (GAP), the actions reported here are assumed to mainly reflect measures implemented during the first decade and a half of the 21^st^ century as countries will have had little time to take new large-scale action since 2015. We therefore investigate the association of these actions with the change in the DPSE categories between the first and second half of the period 2000-2016. We also investigate the relative importance of action compared to covariates related to health systems, economy, human population, livestock production and climate (S2 Table).

In a second set of analyses, we investigate the association between the 16-year trends in DPSE-indicators and the linear trend in subsequent governance responses with the purpose of identifying the prevalence of positive and negative cycles of increasing and decreasing policy ambition, respectively (Fig 1B). A positive cycle is defined as a country experiencing decreases in DPSE indicators during the 2000-2016 period and subsequent increased action during the period of 2016-2022 and a negative cycle if DPSE indicator increased and action decreased. Increases in DPSE indicators and increased action were categorized as countries aiming to “Meet the challenge” and decreases in DPSE indicators and decreased action were categorized as indications of “Relaxed responses” (Fig 1B). Overall data are compiled across 73 countries (S1 Table). For detailed calculations and data sources, see the Supplementary Materials.

### Indicators and data sources

Composite DPSE indicators vary in the number of levels (tiers) at which they can be (dis)aggregated, from one (Exposure) over two (Pressure and State) to three (Drivers). Indicator selection and tiers are mentioned in detail in S2 Table.

#### Drivers – health system

Driving forces behind human antibiotic resistance is captured by analysing the trends in time series data for fifteen variables relating to the general state of the human health system, across four tier 2 indicators including infection prevalence, sanitation standards, vaccination coverage, and health care workforce, which each consist of between two to four tier-3 indicators.

#### Pressure – antibiotic use

We use antibiotic consumption data for humans obtained from the IQVIA database [10] for the years 2000 to 2015. Data is obtained for three tier-2 indicators namely use of broad-spectrum antibiotics [11], newly available antibiotic use [12], and the Daily Defined Dose (DDD) per 1000 inhabitants per day.

#### State – absolute rates of resistance

Three groups of tier-2 indicators of the state of antibiotic resistance were obtained from ResistanceMap [13], namely Methicillin-resistant Staphylococcus aureus, Carbapenem resistance in Enterobacteriaceae and other bacteria, and Streptococcal resistance to macrolides and penicillin.

#### Exposure – relative rates of resistance

We used the Drug Resistance Index (DRI) as an indicator of exposure as it takes into account resistance relative to the use of antibiotics in a country, both of which are important factors in determining likely ABR exposures in the form of ineffective antibiotics [14]. Although the DRI index has been critiqued previously as a standalone indicator [15], we here use it as a part of a multi-indicator analysis to complement patterns in absolute rates of use and resistance.

#### Action – self-assessment survey

Selected questions from the AMR TrACSS dataset (S3 Table) were divided into five thematic categories: awareness & education, monitoring & surveillance, prevention, regulation, and general responses, like progress on NAP implementation (S4 Table). For each country, action scores within categories were calculated by a simple average with individual responses ranging from 0 to 4, in order of increasing ambition. Then the action index was calculated as the average of all five categories equally weighted. Action scores used for the governance syndrome analysis include questions that were asked repeatedly over the years 2016-17 to 2022-23 (S3 Table). Here we calculated the difference between the last year (2022-23, but 2021-22 for Romania) and first year of response (2016-17 for all).

#### Linear and categorical trends

We calculated the average for each DPSE indicator at tier 1 and tier 2 (S2 Table) across the years 2000-2008 (henceforth baseline) and 2008-2016. First, raw data were standardized by standard deviation (SD=1), which allow for comparing indicators that use different units of measurement. Then the national changes in means between two periods was calculated as:

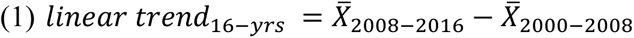

Henceforth referred to as the *linear trend*. We also grouped countries based on the sign of change of the linear trend (increasing vs. decreasing), henceforth referred to as the *categorical trend*.

#### Country selection

Countries included in the various analyses were filtered based on two criteria. First, answering the TrACSS (1.1) survey in 2016-17 (S3 Table). Second, reporting enough years of ABU or ABR indicators to calculate the 16-year trend for at least one of them, meaning at minimum three years in the period 2000-2008 as well as in 2008-2016. Applying these criteria, we are able to analyse 73 countries in total (drivers n=73, ABU n=65, ABR n=35, DRI n=25).

### Statistical analyses

#### Model formulation

We fitted gaussian mixed effect regressions models with the linear trend (equation 2) and the action index as response variable (equation 3), the latter to assess the explanatory ability of the categorical trend for different indicators (S5 Table). We also fitted binomial mixed models with the proportion of declining tier 2 indicators as the response variable weighted by the availability of indicators (equation 4, S6 Table). All models included baselines (*Baseline*) as covariates and a random grouping variable indicating (*Income*) High-Income Countries (HICs) vs. Low-and-Middle Income Countries (LMICs).

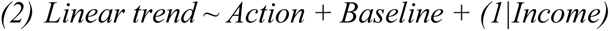

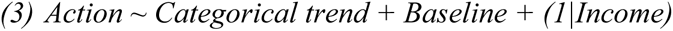

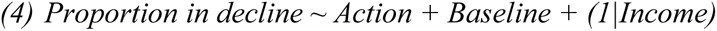

To explore how action various with size of animal production we fit a model with action as response variable and action in the human and animal health sector as explanatory variables.

#### Model selection

We applied model selection to assess the relative importance of action variables compared to other covariates (S7 Table). Here we fit gaussian models to the linear trend and binomial models to the categorical trend and we included countries as random effects, the type of DPSE indicator and country income level as factorial variables and covariates relating to the health system, economic condition, human population, animal production mass, and annual mean temperature (S8 Table). We also allow for pairwise interactions between factorial variables and covariates. We tested the sensitivity to various combinations of country and indicator subsets, resulting in a total of 17 model selection procedures for the linear trend and 16 for the categorial trend where we did not fit models to the Exposure data as there was too little variation in the response variable outcome. We use the R package “MuMIn” [16] and apply the Akaike’s information criterion corrected for small sample sizes (AICc). We report averaged effects for the 95% Akaike weighted subset as well as coefficients from the models with lowest AICc. All statistical analyses were carried out in R 4.3.1 [17]. All global model formulas are detailed in S8 Table.

## Results

### Trends in DPSE indicators

The four tier-1 DPSE indicators exhibit mixed 16-year trends with health system drivers improving (linear trend mean= -0.126 ± 0.017 s.d., p < .001, categorical trend = 6/73 countries increasing, p<0.001), pressures and exposure increasing (ABU: 0.289 ± 0.048, p < .001, 55/65, p < .001; DRI: 0.184 ± 0.065, p =0.01, 21/25, p=0.002), and non-significant trends in the state indicator (ABR: 0.019± 0.069, p =0.783, 16/32, p=1). The increase in ABU is also seen for three of four tier 2 variables (total per capita use 0.329±0.065 p<0.001, 50/65, p<0.001, broad-spectrum antibiotics 0.278 ± 0.069, p<0.001 47/65, p=0.001, newly available antibiotics 0.232 ± 0.061, p<0.001, 55/63, p<0.001). The only tier-2 ABR indicators that showed a consistent increase over time in countries was resistance to last resort carbapenems (20/28, p=0.028).

### Association between DPSE trends and action

At the tier-1 level, action is negatively correlated with three of four DPSE indicators for both linear and categorical trend (Fig 2, S9-S10 Tables). For the linear trend, action is associated with reductions in health system drivers, ABU and DRI (Fig 2 A-D, p<0.05). For the categorical trend, action is associated with ABU, ABR and DRI (Fig. 2 E-H, p<0.05).

**Fig 2.**
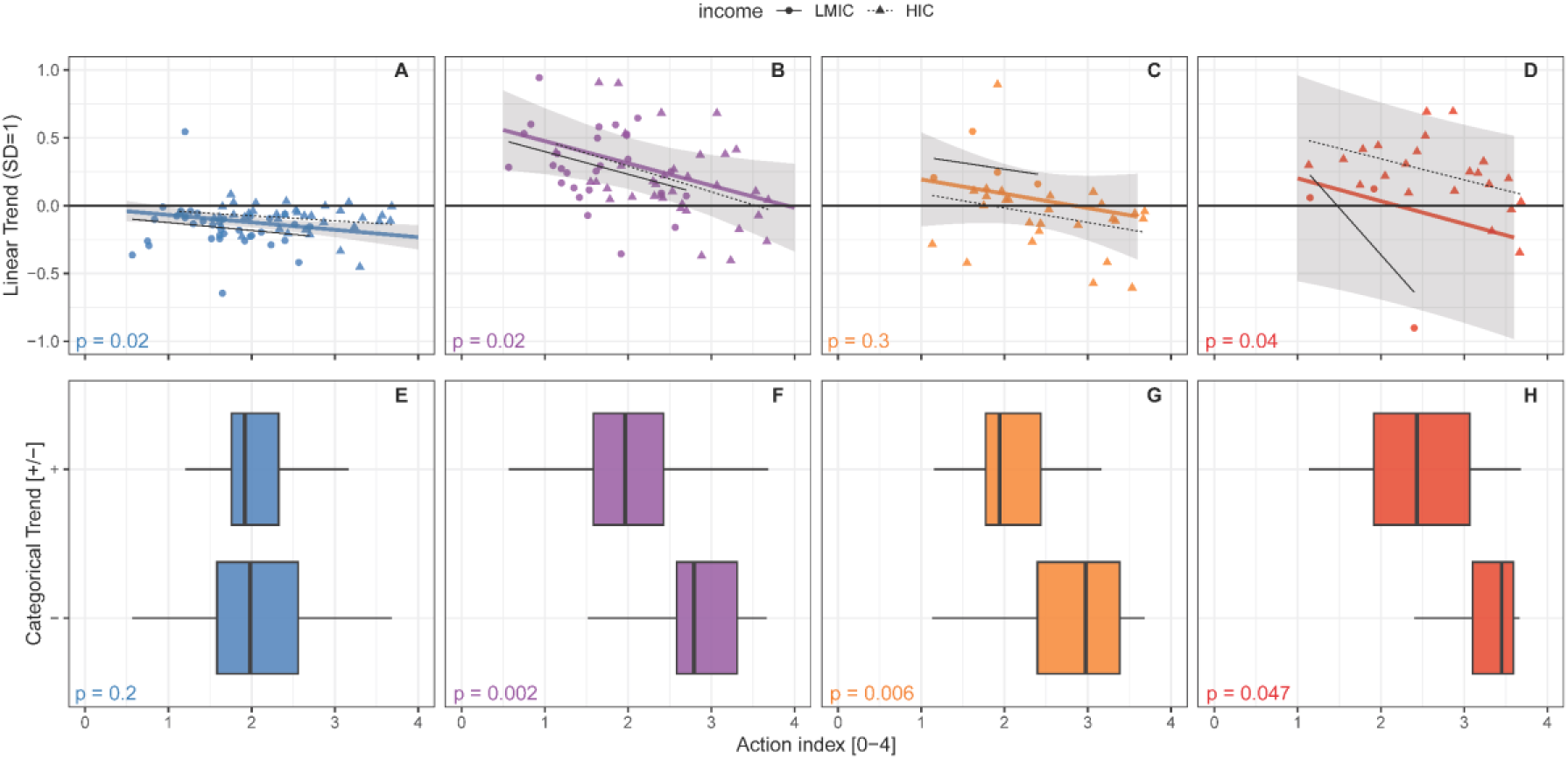
Association between stated action and linear trend (indicator change A-D), and sign of change (categorical trend E-H). Indicator p-values are from linear mixed models with country income group as random effect. For detailed indicators, please see S5 Table. Blue represents drivers of antibiotics resistance, purple represents antibiotics use, orange represents resistance, and red represents DRI.

Complementary analyses indicate that the overall explanatory ability of the action index is (a) not due to a correlation with the baseline state of the indicators (S11 Table) (b), is highest at the level of tier 1 (S9-S10 Tables), and (c) outperforms individual components of the action index (S12-S21 Tables). Thus, individual action components are on average associated with 1 (linear trend) and 1.4 (categorical trend) DPSE indicators, respectively. Of these, the Monitoring and Surveillance component is most commonly associated with DPSE indicators (two of four indicators in both types of models) while Prevention is not associated with any indicators.

The level of action required for improved outcomes (negative categorical trend) in multiple tier-2 indicators or in the case of DRI across multiple countries vary widely for the four DPSE categories (Fig 3, S22-S23 Tables). Here, DRI and ABU exhibits the largest needs for action, converging around an action index of 3.5-3.6 out of 4 for 50 % of variables (ABU) or countries (DRI) to decline (Fig 3). DRI showed signs of a threshold behaviour, with very low probabilities of improvement with action below 2 and 50 % chance of improvement with an action score above 3.5. For ABU, reduction in 25 % of variables was achieved at an action score of 2.5 and 50 % at 3.5. Reduction in resistance, showed a near-linear relationship with action, with 30 % chance at 1.5 and 50 % chance at 2.5. Drivers show overall high probabilities of improvement irrespective of action, but with a slight negative trend as action increases.

**Fig 3.**
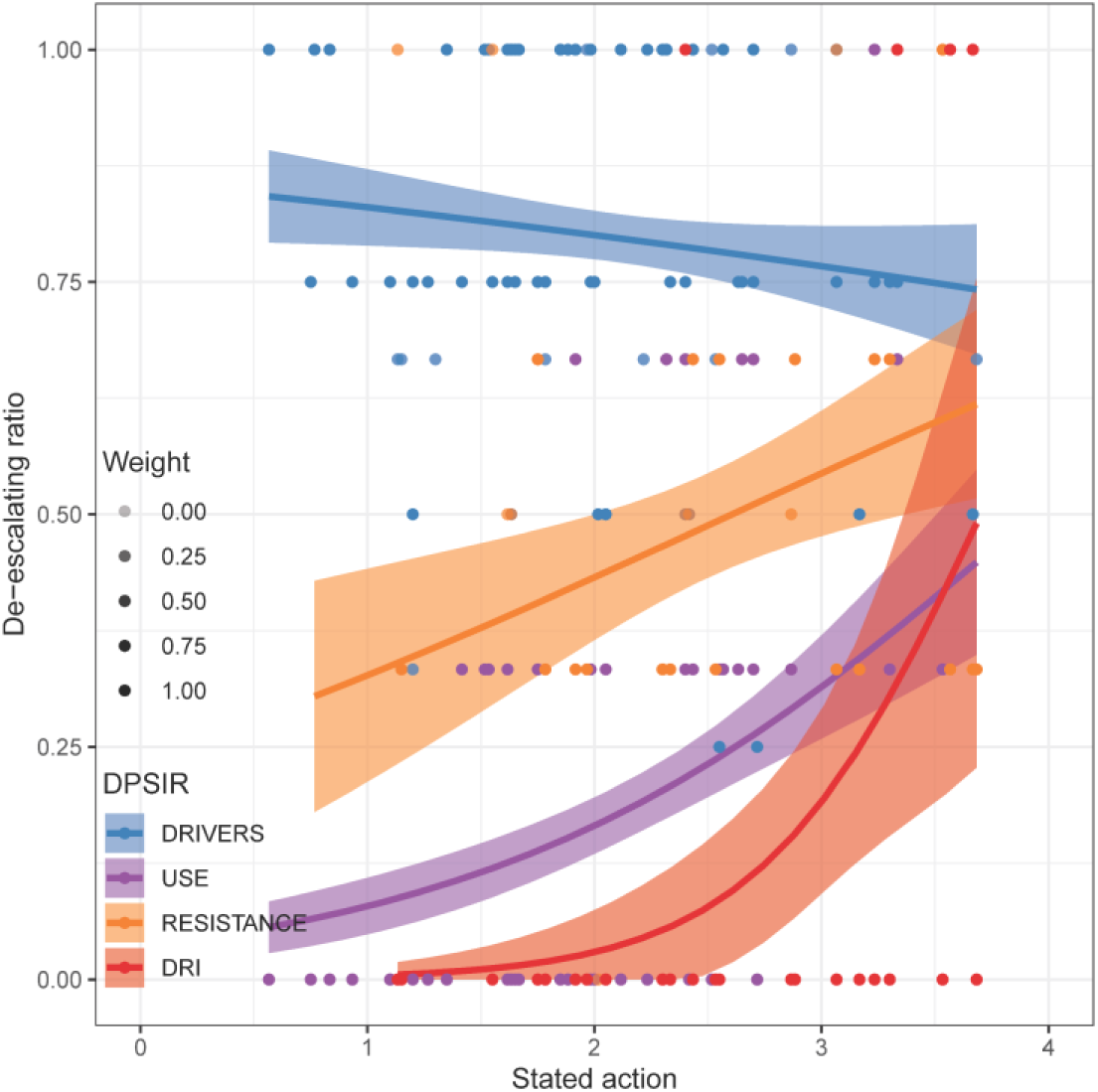
De-escalation of DPSE. De-escalation ratios for Drivers, Use, Resistance, and DRI. De-escalating ratio is defined as the proportion of available lower-level indicators withing a category that have witnessed a reduction from 2000 to 2016. Uncertainty bands indicate standard errors.

### Model selection

Action variables had relatively high levels of importance in model selection with a broader set of explanatory variables, usually ranking in top 3 (Fig 4, S1 Fig, S24 Table), and with consistent negative associations with worsened outcomes (S2-S3 Figs). For linear trends, general action often had a higher importance score than the action index. Here, health system indicators such as workforce were also important and associated with increasing trends for ABU, ABR and DRI. For categorical trends, the action index repeatedly featured in the best selected models (negative coefficients) along with animal production (positive coefficients) and mean temperature (interaction with DPSE) (S3 Fig). Similar effects were also visible when narrowing the analysis down to HICs, but almost absent within LMICs, potentially due to their larger heterogeneity and data scarcity (S4-S6 Figs).

**Fig 4.**
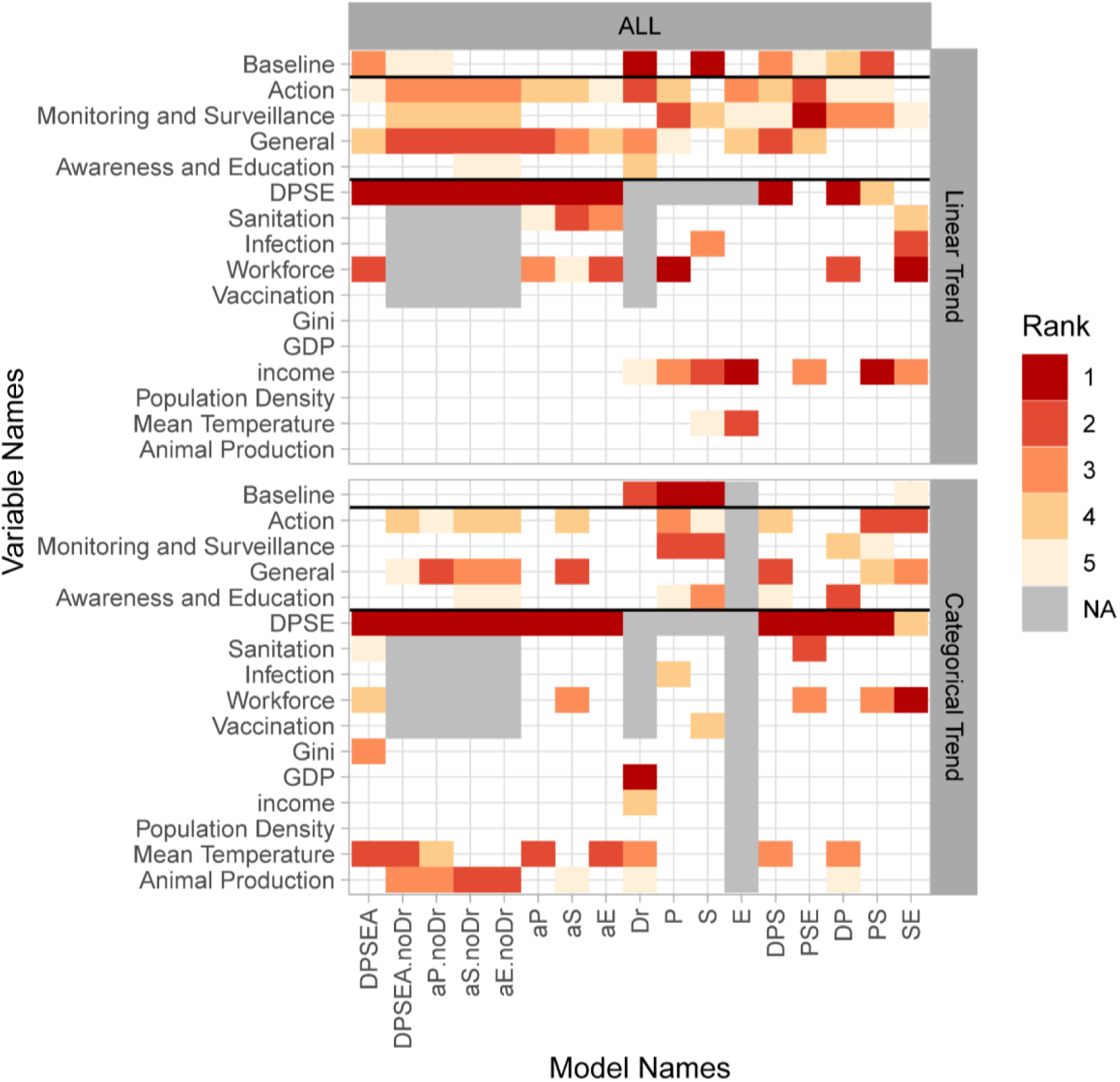
Most important variables in model selection. The rank of the five most important variables (rows) is shown using color coding. Each column represents a unique model selection procedure on the linear trend (change, 17 procedures) or the categorical trend (binomial, 16 procedures). Model names refers to the subset of the dataset with certain DPSE indicators including D (Driver), P (Pressure), S (State), E (Exposure). noDr indicates exclusion of health system variables as explanatory variables. a*X* refers to analysis of DPSE for country subsets with *X* variable available. See S7-S8 Tables for details on each model selection procedure.

### Action in the animal vs human health sector

Countries with larger animal production generally take more ambitious action on animal as well as human health specific issues (p<0.001, slope=0.60, se=0.1, t=6.1, Fig 5). This is true especially in HICs, where animal health specific action was more sensitive to tonnage of animal protein produced than in LMICs (slope diff=-0.61, p<0.001, F=12.6, df =129, Fig 5). Thus, in HICs, large producers of animal protein had higher action scores on specific animal health questions than they had for human health questions, whereas small producers had higher action scores for the human health questions (slope diff=-0.52, p=0.01, df=80, F=6.2, Fig 5A). In LMICs on the other hand, human action scores were generally higher than animal action scores (p diff=0.02, df=179, F=5.8, intercept diff = 0.25, Fig 5B) with both increasing for bigger producers (p slope<0.001, df=179, t=5, slope = 0.29, Fig 5B). The level of action in the largest LMIC producers is significantly lower than in the largest HIC producers pointing to a need for further strengthening action in several of the world’s largest producing countries (Fig 5).

**Fig 5.**
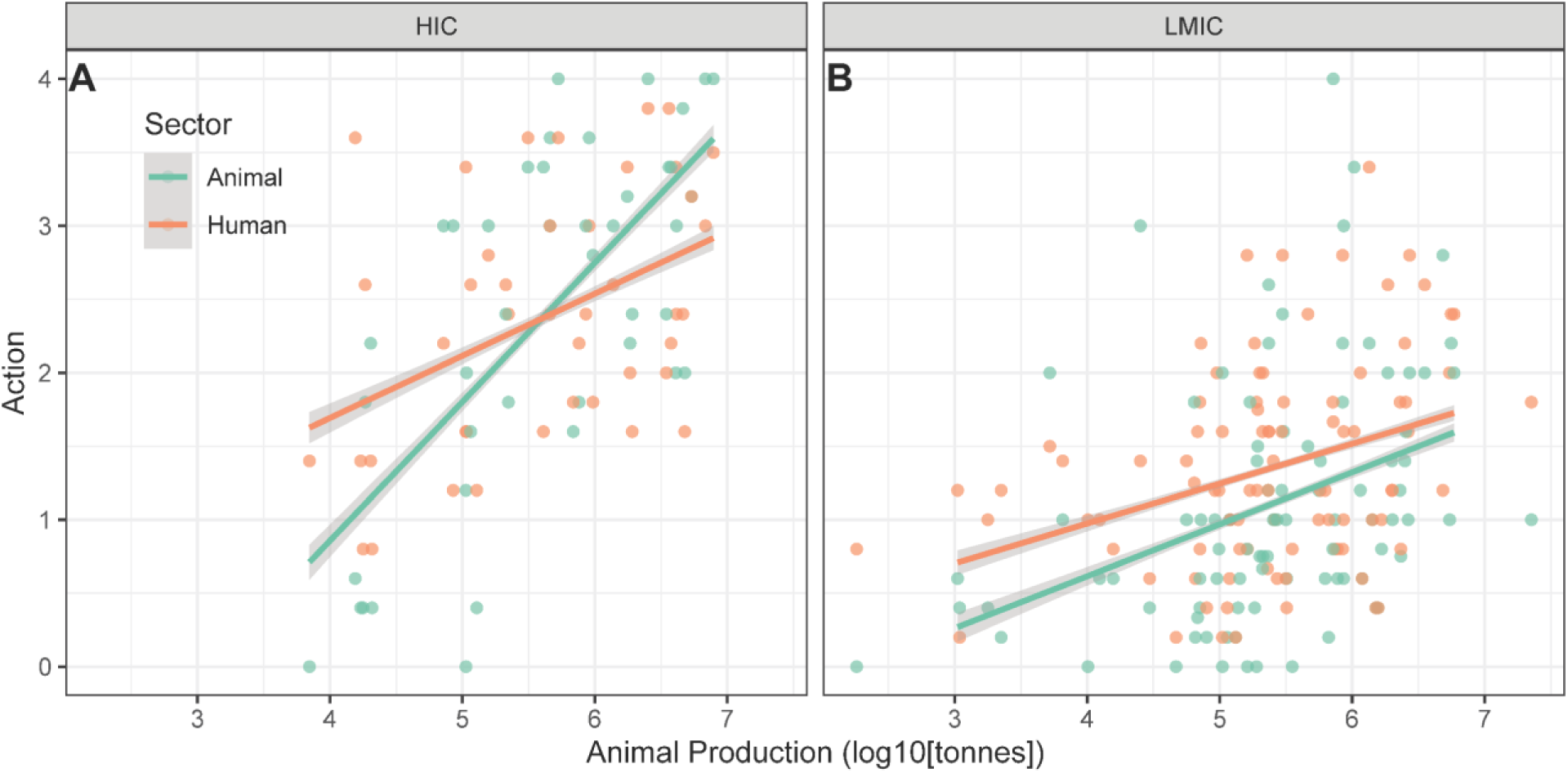
Actions levels in animal protein produced countries for animal and human related Antibitotic Indicators. Action level of governments belong to countries with large producers of animal protein. Stated government action animal and human health scores in relation to total production of vertebrate biomass (mammals, birds and fish) for high-income (HICs) and low- and middle-income countries (LMICs).

### Changes in action over time

Three quarters (76 %) of countries analysed for association with DPSE indicators in 2016 report increased action between 2016 and 2023. This is the case in both HICs (70 %) as well as in LMICs (83 %). However, almost 25 % of countries lowered their ambition including almost 30 % of HICs (all=23 %, HIC=30 %, LMIC=17 %, Fig 6). For drivers, we see that more than half of HICs and LMICs are in a potential positive cycle with improved conditions and subsequent increased policy ambition. However, this proportion drops markedly for ABU, ABR, and DRI. For ABU in LMICs, 3 out of 31 countries (10 %) are in a positive cycle and 6 in a negative cycle, but 70 % are meeting the challenge of increasing ABU with increased policy responses. For HICs, negative cycles are fairly frequent for ABU (24 %, n=34) and DRI (32 %, n=22) and positive cycles very rare for DRI (5 %, n=22).

**Fig 6.**
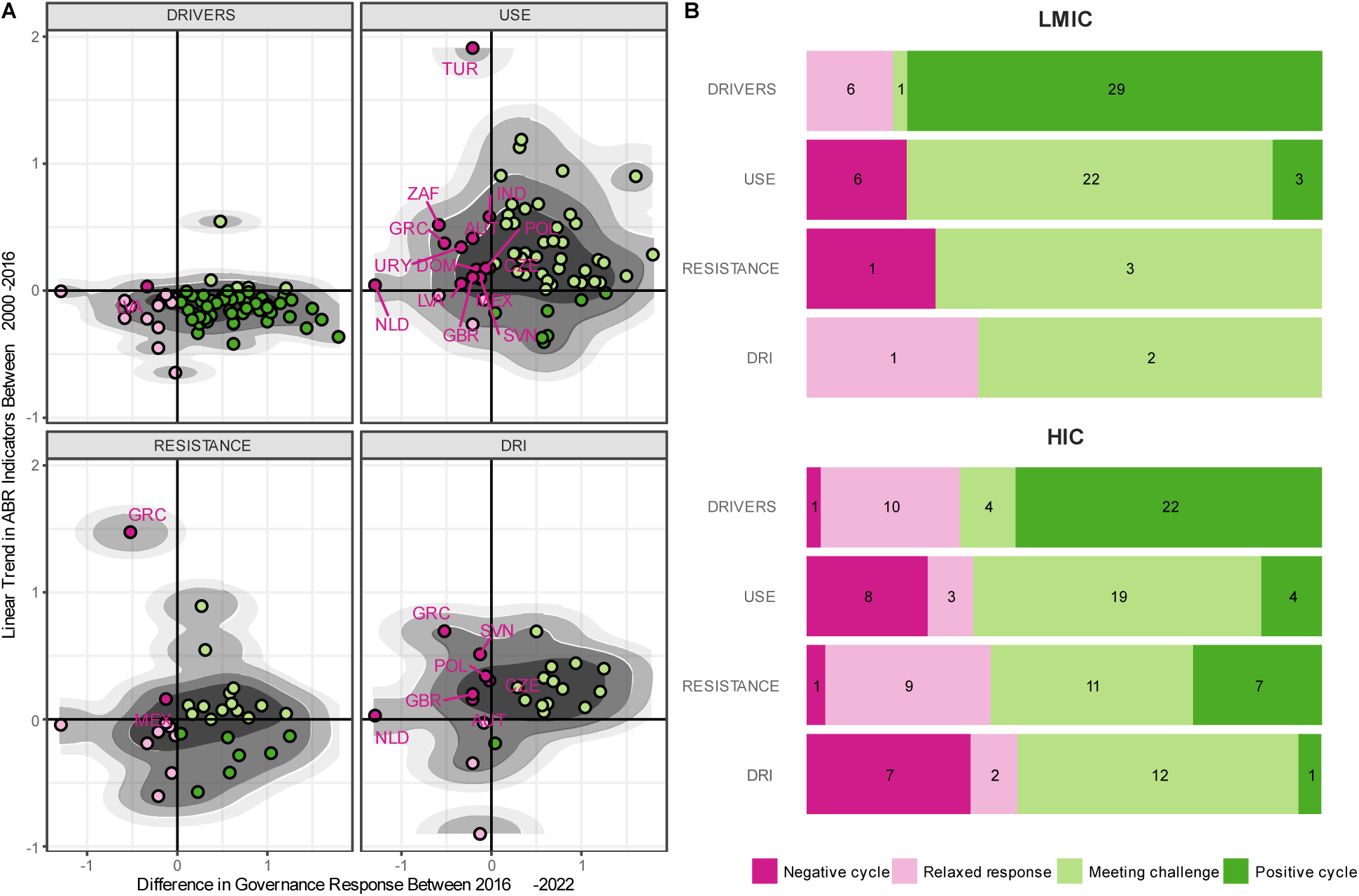
Classification of country ABR governance syndrome. (A) categorize country trajectory based on trend in ABR indicators and governmental action. Countries in negative cycle are displayed with name ISO3 code (see S1 Table). (B) Comparison of country governance syndrome according to DPSE indicators and income level.

## Discussion

Our results show associations between self-reported action in 2016 and trends in the preceding 16 years in health system drivers, ABU, ABR and DRI. These results are robust to inclusion of other contextual variables in multivariate models, strongest at the aggregate scale of both DPSE and action indicators, and not due to a correlation with the baseline state of indicators. The results indicate that every step of improved action is important to reduce the magnitude of adverse outcomes of increasing ABU and DRI (Fig 2 B, D), and that high levels of action can help achieve reductions in ABU, ABR and DRI (Fig 2 F-H). Here we discuss the interpretation of the results, extent of some potential limitations as well as consequences for policy.

### National action and DPSE indicators

National action on antibiotic resistance has been critiqued for not being well enough funded, national action plans (NAP) for not being fully developed and operationalized, with some indications of adoption of standard templates [18–21]. These concerns about the state of national action increase the uncertainty between data reported and action taken on the ground. For example, we would expect template responses to make it harder to detect an association between reported actions and DPSE indicator trends as it makes it harder to distinguish between countries implementing contextually adapted and well-funded policies and countries reporting action that is not adapted to national needs or not well funded. As we compare trends in DPSE indicators leading up to the 2016 survey, our study design implies a conservative estimate of the association between action and trends in DPSE indicators as some recently taken action will likely not have had an effect on the trends. That we detect these associations even with the above-mentioned caveats indicates that well implemented action probably has fairly large and measurable effects. It also means that the level of action needed for reductions in ABU, ABR, and DRI estimated here are likely safe levels and that reductions might be achieved at lower levels.

We find that the action index more consistently explains univariate changes in the four composite DPSE indicators than any of its individual components and that the explanatory ability of the action index is most consistent at the highest level of aggregation of the DPSE indicators (S9-S21 Tables). These results indicate the importance of assessing aggregate policy responses and evaluating their potential impacts on combined indices of multiple indicators. This approach helps account for cross-country variation in social, economic and environmental context and related variation in needs for action. Monitoring and surveillance and general action (such as early progress on NAP development) also explain variation in trends in several multivariate models. Here, general action likely indicates the importance of responding systemically through coordination across sectors e.g., through NAPs [20–22]. Monitoring and Surveillance performs well in multivariate model selection for models limited to ABU, ABR and DRI as well as models limited to HICs. The importance of this variable could indicate its importance in enabling informed decision making and adaptive management of both the use of antibiotics and containment of resistant variants [23].

DRI is the hardest of the indicators to achieve reductions in, followed by ABU and ABR. Taken together, we interpret these patterns as indications that absolute rates of resistance can more easily be mitigated by e.g., cycling antibiotics, while reducing absolutes rates of use requires a wider infection prevention and control strategy. Finally relative rates of resistance may also require improving use of antibiotics across the board to mitigate selection pressures on resistance e.g., when a new antibiotic is adopted. The linear trend in ABR shows fairly weak correlation with action, whereas the categorical trend is consistently correlated. The former pattern could be an indication of (1) a non-linear association between action and ABR; (2) that bacteria targeted for management vary by country and potentially go beyond the three indicators available for this analysis, or; (3) that absolute rates of resistance can be circumvented through various aforementioned strategies. The latter pattern could indicate a threshold of action above which such reductions are more likely to be achieved. Overall, national action has a fairly small effect size on the linear trend in health system drivers at ca. 0.25 s.d. across the range of the action index compared to 0.5 s.d. for ABU and DRI. This could be because health system improvements are driven by other policy instruments rather than AMR specific policy. The weakly negative association between action and trends in drivers might be due to improvements in drivers being much harder to achieve for countries with more developed health systems where almost the entirely population has access to sanitation and vaccines, infection rates are low and where health care workforces are already high.

### Health systems, climate and animal production

Model selection reveals that multiple contextual factors help explain variation in DPSE indicator trends, mainly additional to that associated with national action. These include variables related to health system development, climate and animal production. The relationships between health systems on antibiotic resistance is multifaceted. While the highest burden of antibiotic resistance is often due to lack of access to healthcare and antibiotics, health systems are also associated with higher levels of regulated use [2,24,25]. These consequences of equitable access might be driving the positive association between health workforce and the linear trend in ABU, ABR and DRI in multivariate models.

A positive association between warmer mean temperatures and the categorical trend of DPSE indicators is likely not due to collinearity with GDP per capita, inequality, or country income level as these are all considered as covariates (Fig 4). Possible explanations include a potential non-linear effect of climate biological factors such as bacterial growth rates [26] or horizontal gene transfer [27] or alternatively, effects of one or more social, economic or environmental factors that covary with climate that we were not able to include in this analysis.

Animal production is positively associated with the categorical trend of DPSE indicators in four models that do not include health system variables (Fig 4). While the importance of animal production is lower when health system variables are included, the pattern is still worth examining given the potential concern about any spill over effect from the animal sector to the human health sector. Multiple explanations can be hypothesized for such patterns, including spill over of residues, resistance genes, or cultural factors relating to antibiotic use, to name a few [28]. Whatever any causal explanations might be, these patterns indicate that additional action in both the animal and human sectors may be needed to achieve indicator reductions in large animal producing countries. This challenge should not be underestimated as countries with large animal production are already reporting higher levels of actions in both the human and animal health sector (Fig 5). Reducing the spread and elevating the lower level of action in large producing countries could help reduce any spill over effects from animal production on DPSE indicators in human health.

### Changes in action and conclusions

While most countries increase their level of action in 2023 compared with 2016-17, around a quarter reduce their levels of action. Reductions in the action index might reflect reduced prioritisation of the policies in the ABR area e.g., during the pandemic and economic disruption [5], but might also be due to enhanced accuracy of the data reported in the TrACSS survey compared to the first years. Improved accuracy in reporting would influence findings regarding governance syndromes by inflating the number of countries in the ‘relaxed response’ or ‘negative cycle’ categories. Given the high levels of action associated with reductions of ABU and DRI, it is expected that positive cycles are the least common in these two categories. That is in part due to countries with high levels of action that reduced ABU and DRI having less options to improve their score in 2023. Going forward it will be important to find governance mechanisms, that enable countries to progress in taking new action while also focusing on implementing and securing sustainable funding for currently planned actions.

In conclusion, our analysis indicates the importance for governments to take ambitious and comprehensive action on AMR across sectors to improve the situation with regard to antibiotic resistance while maintaining or improving access to the services of highly functioning health systems. At the same time, in the event reductions cannot yet be achieved, each additionally implemented action is likely to help reduce the adversity of trends in antibiotic use and relative rates of resistance. Future studies should assess the association between national action and trends in indicators since the agreement of the GAP.

## Data Availability

All data produced willlbe available online via Github.

https://github.com/PSJorgensen/

## Acknowledgments

We acknowledge funding from the Erling-Persson Family Foundation (L.N.T., P.S.J), the European Union (ERC, INFLUX, 101039376, P.S.J.), the IKEA Foundation (P.S.J.), the Marianne and Marcus Wallenberg Foundation (P.S.J) and the Uppsala Antibiotic Centre (UAC, L.N.T.). The funders had no role in the design, analysis, or writing of this paper. Views and opinions expressed are, however, those of the author(s) only and do not necessarily reflect those of the European Union or the European Research Council. Neither the European Union nor the granting authorities can be held responsible for them. We thank all participants of the SESYNC Living with Resistance pursuit for their participation.

