## Supplementary materials for "Association between national action and trends in antibiotic resistance: an analysis of 73 countries from 2000 to 2023"

[6] Arizona State University, Department of Psychology, Tempe, AZ 85281, USA

[7] North Carolina State University, Department of Applied Ecology, Raleigh, NC 27695-7617, USA

### Supplementary Materials

### Supplementary Methods

#### DPSEA Indicators

**Drivers – health system:** Driving forces behind human antibiotic use is captured by analyzing the trends in time series data for fifteen variables across four tier 2 indicators including infection prevalence (primary driver), sanitation standards, vaccination coverage, and health care workforce. Total data for drivers are compiled for a total of 219 countries from the United Nations (UN), World Bank database and the World Health Organization (WHO). Data availability varies by indicator as detailed in S2 Table.

**Pressure - antibiotic use (ABU):** Data from QuintilesIMS are estimates of the total volume of sales of each antibiotic molecule (or combination of molecules) based on national sample surveys of antibiotic sales. Antibiotic consumption data are in kilograms and converted into defined daily doses (DDDs) using the Anatomical Therapeutic Chemical Classification System (ATC/DDD, 2016) developed by the WHO Collaborating Centre for Drug Statistics Methodology as in Klein et al. [1].

**State - resistance:** Data obtained from ResistanceMap [2] which is a repository of global antimicrobial resistance data. ResistanceMap obtains data from public and private sources, including lab networks, hospitals, and government agencies. Data include resistance rates for eight high-priority pathogens isolated from blood and cerebrospinal fluid of patients and are aggregated at the country level on an annual basis. Data on ResistanceMap has been harmonized to present similar definitions of resistance across countries and regions to enable comparisons between countries.

**Exposure – Drug resistance index:** The Drug Resistance Index (DRI) combines use and resistance rates into a single value that provides measures of antibiotic effectiveness relative to their use [3]. While DRI has been critiqued when used as a single indicator of antibiotic effectiveness [4], we here use it as part of a multi-indicator framework. We calculated the adaptive Drug Resistance Index for countries for which data on resistance and use is available over the time period following the methodology outlined in [3] and [5]. Briefly, the annual DRI was estimated for each country by the following equation:

$$DRI = \sum_k \rho_k^{i,t} q_k^{i,t} \qquad (1)$$

where, for country $i$ at time $t$, $\rho_k^{i,t}$ is the proportion of resistance among all included organisms to drug $k$ and $q_k^{i,t}$ is the proportion of drug $k$ used for their treatment in all drugs included in the index. Pathogens included in the analysis were *E. coli*, *K. pneumoniae*, *P. aeruginosa*, *S. aureus*, *E. faecium*, and *E. faecalis*. Antibiotics included in the analysis were aminoglycosides, broad-spectrum penicillin, carbapenems, cephalosporins, narrow-spectrum penicillin, and quinolones. Because not all countries had data for all combinations, we included a country if they had at least four of the six organisms, and 10 of the 17 total combinations possible (S2 Table).

**Action – TrACSS:** All action indicators are self-reported data from the Global Database for Tracking Antimicrobial Resistance Country Self-Assessment Survey (TrACSS) spanning the period of 2016-2023 (https://amrcountryprogress.org/). The survey responses are publicly available with the yearly updated version providing information about countries ongoing actions to live up to the global action plan on antimicrobial resistance All answers are given on an ordinal scale from A to E (0-4).

**Figure S1. Importance Scores of Variables in Model Selection from Averaged Models.** For model name descriptions and formulas see S7 and S8 Table. Interaction terms shown with ":", variables excluded from the explanatory variables shown as NA. It includes all countries regardless of income.

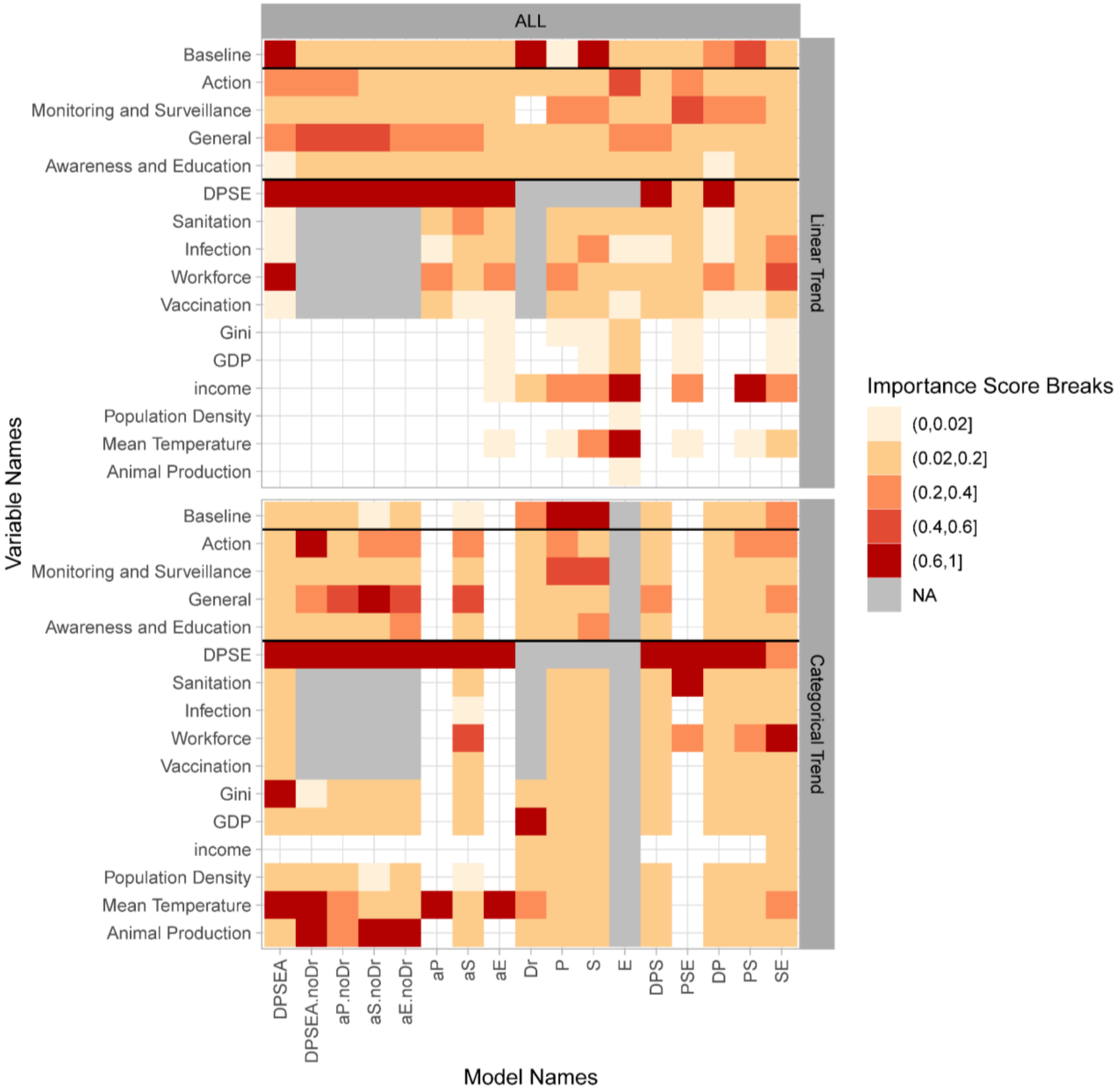

**Figure S2. Coefficient Estimates of Averaged Models for All Countries in Linear Trend and Categorical Trend Models.**
For model name descriptions and formulas see S5 Table. Variables excluded from the explanatory variables shown as NA. It includes all countries regardless of income.

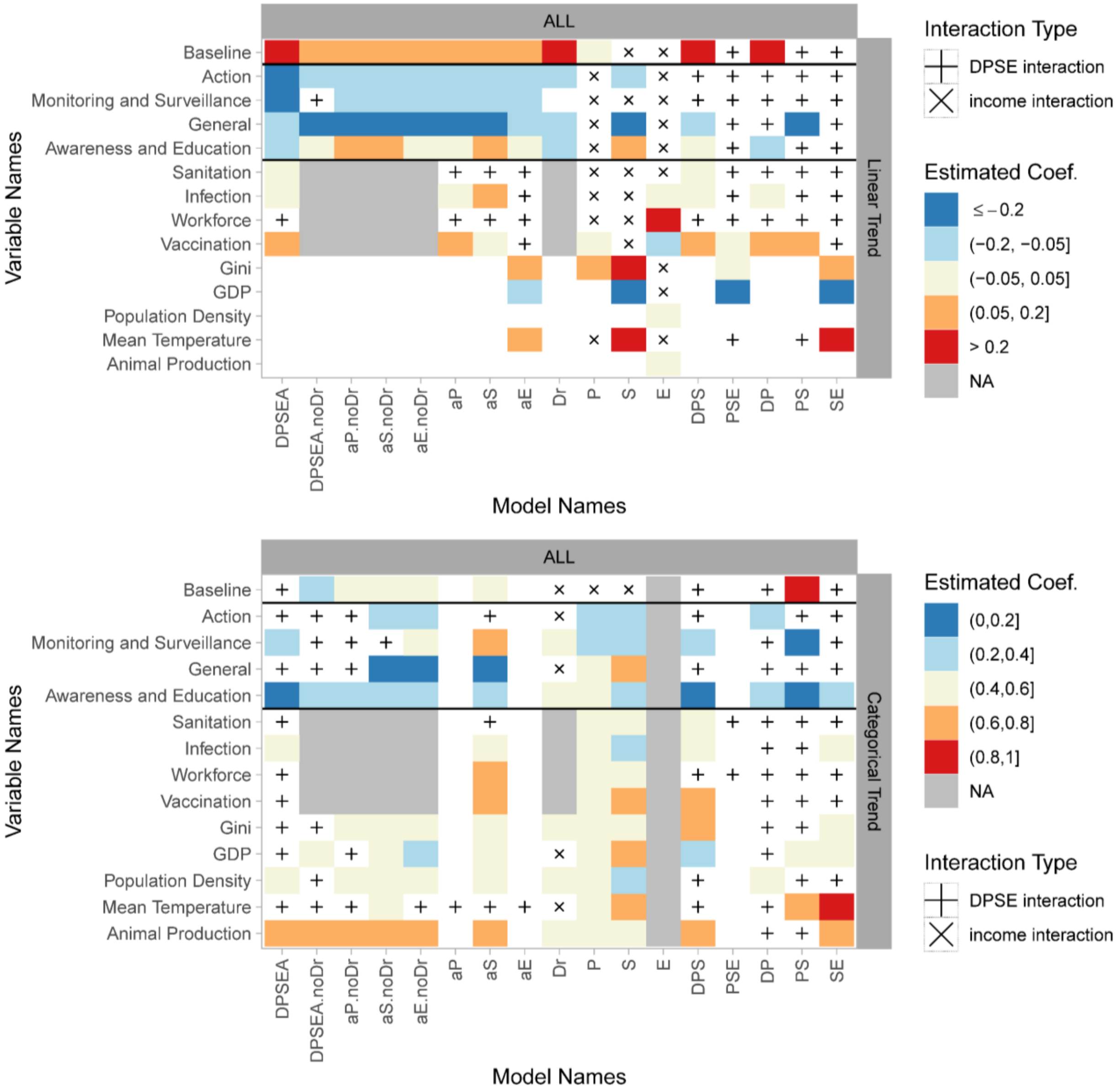

**Figure S3. Coefficient Estimates of Variables from Best Selected Models from Model Selection.**
For model names see and formulas see S7 and S8 Table. Figure only included main variables. Variables excluded from the explanatory variables shown as NA. It includes all countries regardless of income.

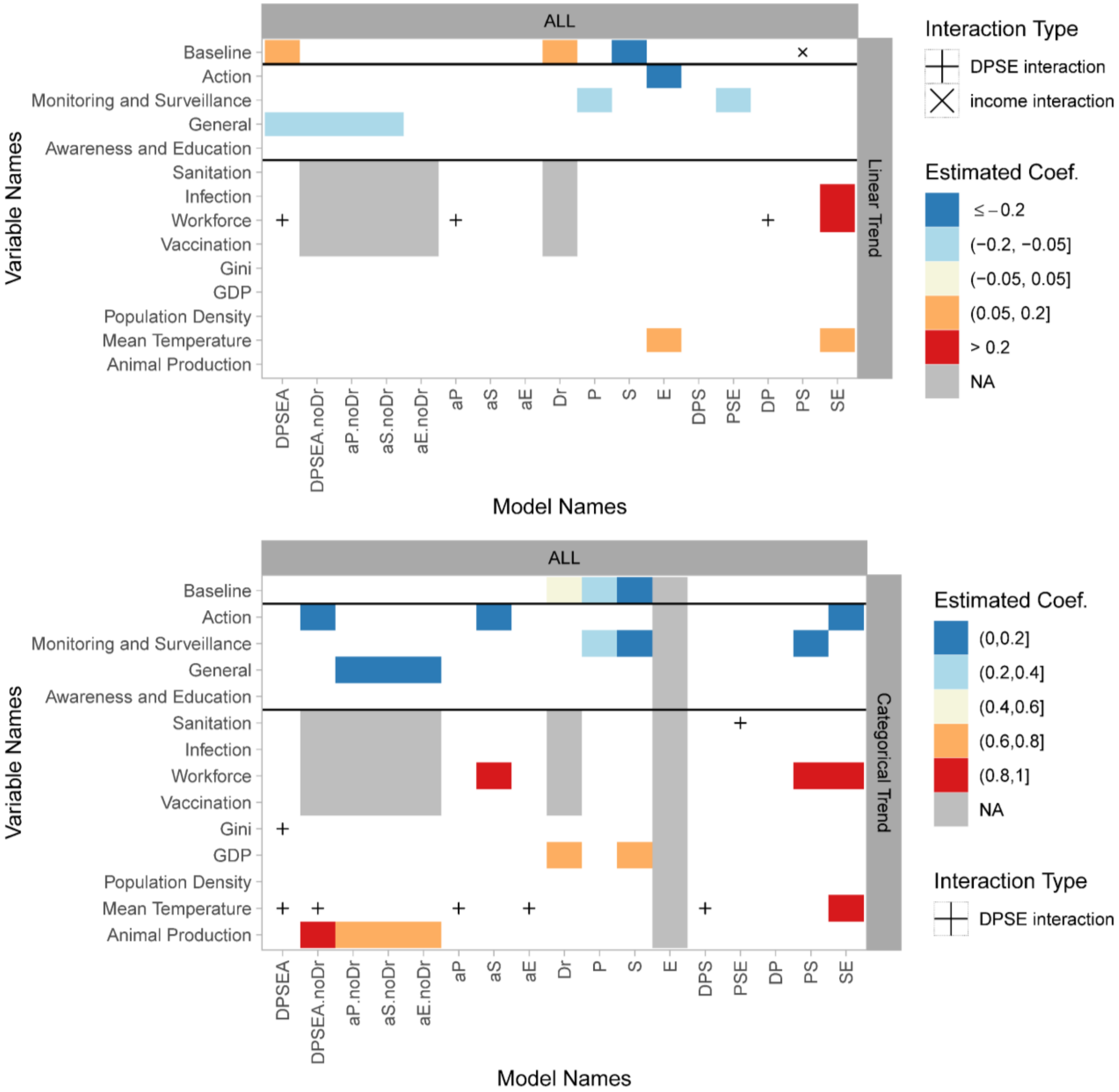

**Figure S4. Rank of Variables in Averaged Models for Different Income**
For model name descriptions and formulas see S7 and S8 Table. Interaction terms shown with ":". Variables excluded from the explanatory variables shown as NA. HIC refers to High Income Countries, LMIC refers to Low-and-Middle Income Countries.

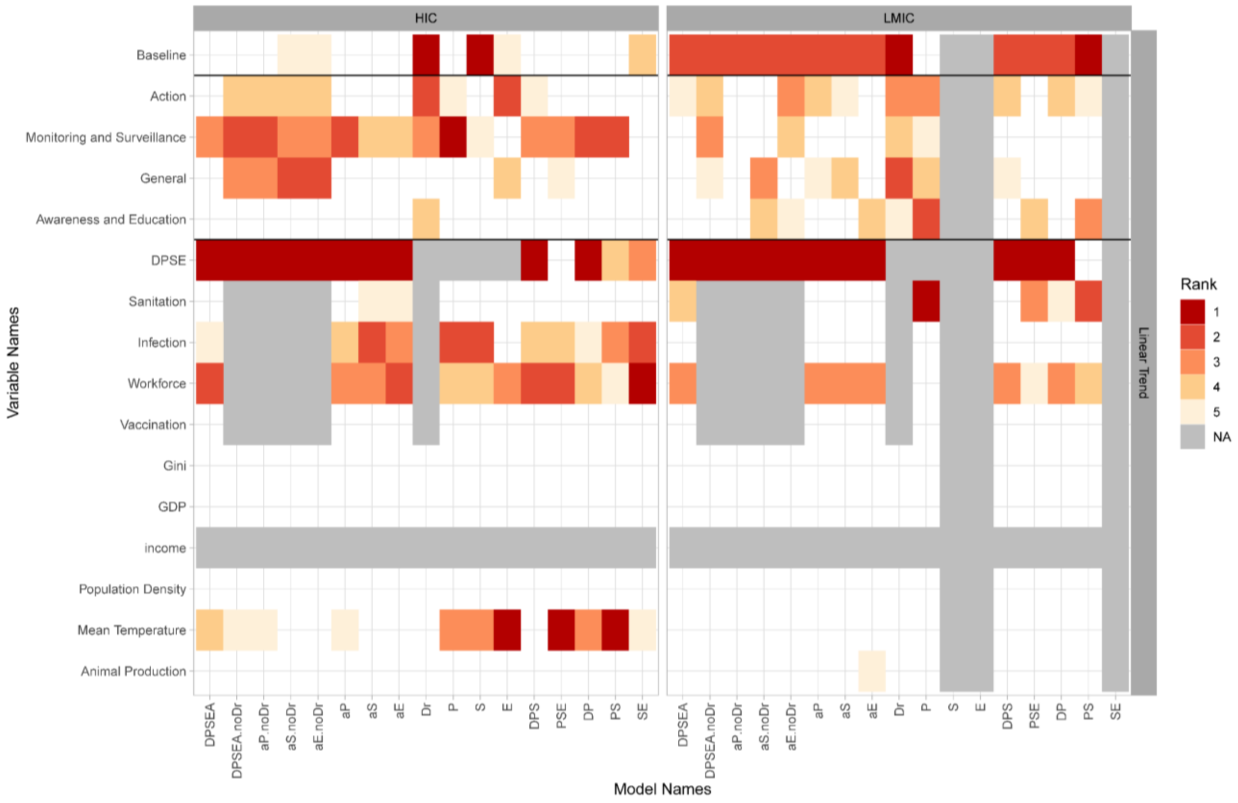

**Figure S5. Importance Scores of Variables in Model Selection from Averaged Models for Different income Groups.**
For model name descriptions and formulas see S7 and S8 Table. Interaction terms shown with ":". Variables excluded from the explanatory variables shown as NA. HIC refers to High Income Countries, LMIC refers to Low-and-Middle Income Countries.

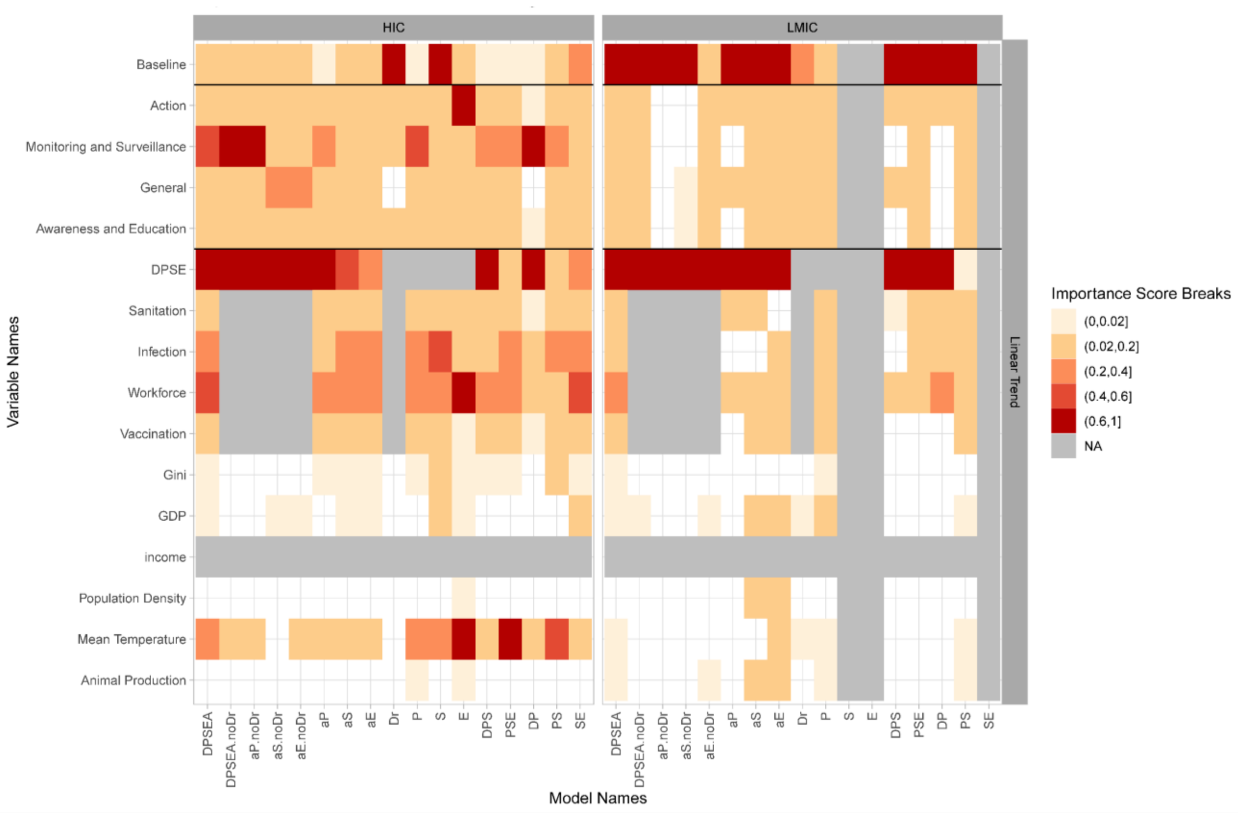

**Figure S6. Coefficient Estimates of Averaged Models for Countries with Different Income Groups**

For model name descriptions and formulas see S7 and S8 Table. Variables excluded from the explanatory variables shown as NA. HIC refers to High Income Countries, LMIC refers to Low-and-Middle Income Countries.

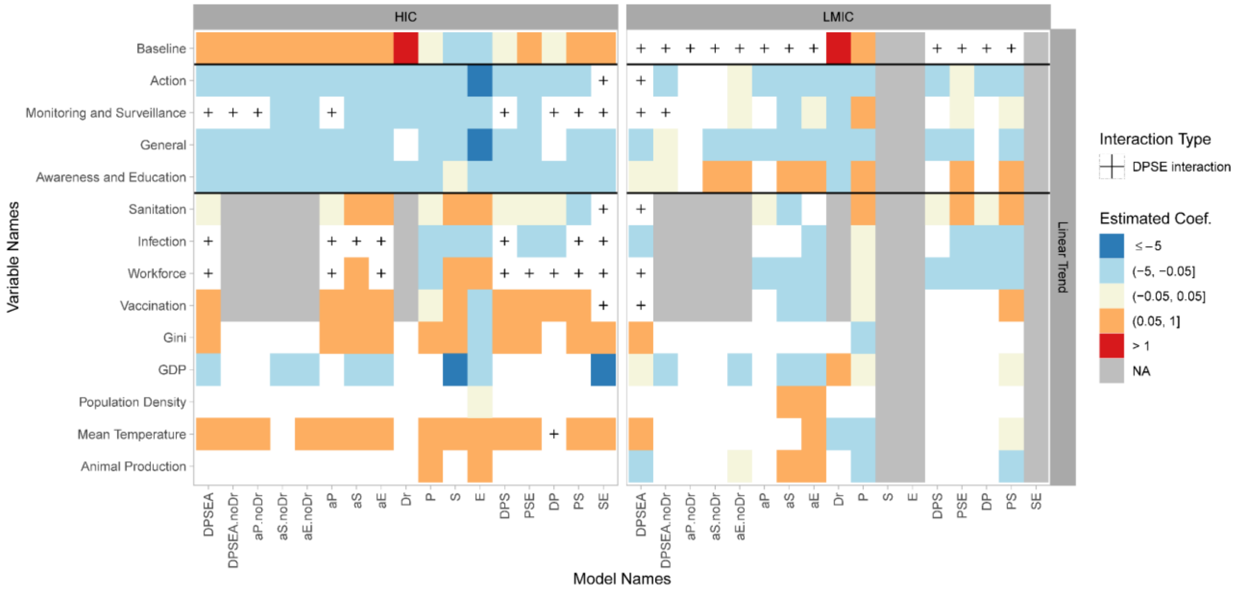

### TABLES

**S1 Table. List of countries included in the study.**
The total number of countries included in the study is 73. ISO3 codes refers to three-letter country codes according to ISO 3166-1. Income levels: HIC refers for High Income Countries, LMIC refers for Low- and Middle-Income Countries.

| ISO3 | Country | Income |
|---|---|---|
| **Asia** | | |
| ARE | United Arab Emirates | HIC |
| BGD | Bangladesh | LMIC |
| CHN | China | LMIC |
| IDN | Indonesia | LMIC |
| IND | India | LMIC |
| JOR | Jordan | LMIC |
| JPN | Japan | HIC |
| KAZ | Kazakhstan | LMIC |
| KOR | Republic of Korea | HIC |
| LBN | Lebanon | LMIC |
| LKA | Sri Lanka | LMIC |
| MYS | Malaysia | LMIC |
| PAK | Pakistan | LMIC |
| PHL | Philippines | LMIC |
| SAU | Saudi Arabia | HIC |
| SGP | Singapore | HIC |
| THA | Thailand | LMIC |
| TUR | Turkey | LMIC |
| UKR | Ukraine | LMIC |
| VNM | Vietnam | LMIC |
| **South America** | | |
| ARG | Argentina | LMIC |
| BRA | Brazil | LMIC |
| CHL | Chile | LMIC |
| COL | Colombia | LMIC |
| ECU | Ecuador | LMIC |
| PER | Peru | LMIC |
| URY | Uruguay | LMIC |
| **Oceania** | | |
| AUS | Australia | HIC |
| NZL | New Zealand | HIC |
| **Europe** | | |
| AUT | Austria | HIC |

| ISO3 | Country | Income |
|---|---|---|
| BEL | Belgium | HIC |
| BGR | Bulgaria | LMIC |
| BLR | Belarus | LMIC |
| CHE | Switzerland | HIC |
| CYP | Cyprus | HIC |
| CZE | Czech Republic | HIC |
| DEU | Germany | HIC |
| DNK | Denmark | HIC |
| ESP | Spain | HIC |
| EST | Estonia | HIC |
| FIN | Finland | HIC |
| FRA | France | HIC |
| GBR | United Kingdom | HIC |
| GRC | Greece | HIC |
| HRV | Croatia | HIC |
| HUN | Hungary | HIC |
| IRL | Ireland | HIC |
| ISL | Iceland | HIC |
| ITA | Italy | HIC |
| LTU | Lithuania | LMIC |
| LUX | Luxembourg | HIC |
| LVA | Latvia | HIC |
| MKD | North Macedonia | LMIC |
| MLT | Malta | HIC |
| NLD | Netherlands | HIC |
| NOR | Norway | HIC |
| POL | Poland | HIC |
| PRT | Portugal | HIC |
| ROU | Romania | LMIC |
| RUS | Russian Federation | LMIC |
| SRB | Serbia | LMIC |
| SVK | Slovakia | HIC |
| SVN | Slovenia | HIC |
| SWE | Sweden | HIC |
| **North America** | | |
| CAN | Canada | HIC |
| DOM | Dominican Republic | LMIC |
| MEX | Mexico | LMIC |
| USA | United States of America | HIC |
| **Africa** | | |
| EGY | Egypt | LMIC |
| GHA | Ghana | LMIC |
| MAR | Morocco | LMIC |

| ISO3 | Country | Income |
|---|---|---|
| TUN | Tunisia | LMIC |
| ZAF | South Africa | LMIC |

**S2 Table. Indicator selection for Driver, Use, Resistance, and DRI categories.**
DPSE indicators (Driver, Pressure, State, and Exposure) and tiers included for the calculation of each DPSE indicator variable. Driver data was available for 148 countries, while use or resistance data were available for 73 countries with ABU (Antibiotic Usage), ABR (Antibiotic Resistance) and DRI (Drug-Resistance Index) data available for 65, 32, and 25 countries respectively. Data for governmental responses to ABR were collected from a recent survey of stated levels of national government action to limit antimicrobial resistance. Additionally, countries which had driver data only included if they had one of the other indicator categories (Use, Resistance, and DRI), hence only 73 countries included in drivers (data is standardised SD=1 by denoting the ratio of standard deviation to mean). Drivers data included 3 tiers, use and resistance included 2 tiers of indicators. Average of tier 3 indicators are taken from different data sources.

| **DRIVERS: FACTORS INFLUENCING ANTIBIOTIC USE** | | | | | | |
|---|---|---|---|---|---|---|
| **TIER 1** | **TIER 2** | **DESCRIPTION** | **TIER 3** | **DESCRIPTION** | **DESCRIPTION** | **UNIT** |
| **Drivers (Total)** | **Sanitation** | **Hygiene and Sanitation Standards (+)** | Drinking Water Source | Proportion of population using improved drinking water sources | United Nations (UN) | pct (%) |
| | | | Drinking Water Source | Population using improved drinking-water sources (%) | United Nations (UN) | pct (%) |
| | | | Overall Sanitation | Population using improved sanitation facilities (%) | United Nations | pct (%) |
| | | | Water Source Access | Improved water source (% of population with access) | United Nations | pct (%) |
| | **Infection** | **Infection Prevalence (-)** | HIV Prevalence | UN Prevalence of HIV Total (% of population ages 15-49) | United Nations (UN) | pct (%) |
| | | | HIV Prevalence | WB Prevalence of HIV Total (% of population ages 15-49) | World Bank (WB) | pct (%) |
| | | | Tuberculosis Prevalence (Tub) | UN Prevalence of tuberculosis (per 100 000 population | United Nations (UN) | Per 100 000 |

| | | | | | | |
|---|---|---|---|---|---|---|
| | | | Incidence of Tuberculosis (Tub) | WB Incidence of tuberculosis (per 100,000 people) | World Bank (WB) | Per 100 000 |
| | **Vaccination** | **Vaccination Coverage (+)** | DTP3 | Diphtheria, Tetanus, and Pertussis) immunization coverage among 1-year-olds (%) | World Health Organization (WHO) | pct (%) |
| | | | HepB3 | Hepatitis B ) immunization coverage among 1-year-olds (%) | WHO | pct (%) |
| | | | Hib3 | Haemophilus influenzae type b) immunization coverage among 1-year-olds (%) | WHO | pct (%) |
| | | | PCV3 | Pneumococcal conjugate vaccines immunization coverage among 1-year-olds (%) | WHO | pct (%) |
| | | | Pol3 | Polio immunization coverage among 1-year-olds (%) | WHO | pct (%) |
| | | | Measles | Immunization, measles (% of children ages 12-23 months) | United Nations and World Bank data average | pct (%) |
| | | | RCV1 | Rubella vaccination coverage | Rubella | pct (%) |
| | **Workforce** | **Health Care Workforce (+)** | Physicians per capita | Physicians density (per 1000 population) | UN | per 1000 |
| | | | Physicians per capita | Physicians density (per 1000 population) | WHO | per 1000 |
| | | | Nursing and midwifery | Nursing and midwifery personnel density per 1000 population | UN | per 1000 |
| | | | Nursing and midwifery | Nursing and midwifery | WHO | per 1000 |

| | | | | |
|---|---|---|---|---|
| | | | | personnel density per 1000 population |

**USE: PRESSURES FOR RESISTANCE TO SPREAD**

| TIER 1 | TIER 2 | Details | DESCRIPTION | UNIT |
|---|---|---|---|---|
| **Use (Total)** | BroadPerTotalABXUse | Broad spectrum antibiotics including Fluoroquinolones, Macrolides, | Use of broad-spectrum antibiotics. | Defined Daily Doses (DDD) |
| | | third-generation cephalosporins, | | |
| | | co-amoxiclav, clindamycin, oral vancomycin | | |
| | NewABXUse | New Antibiotics and their year of introduction: dalfopristin/ quinupristin (2005), | Newly Available Antibiotic Use (defined as antibiotics first introduced in 1999 or later [1]) | DDD |
| | | gatifloxacin (1999), | | |
| | | moxifloxacin (1999), | | |
| | | linezolid (2000), | | |
| | | telithromycin (2001), | | |
| | | balofloxacin (2006), | | |
| | | Biapenem (2002), | | |
| | | ertapenem (2002), | | |
| | | pazufloxacin (2002), | | |
| | | prulifloxacin (2002), | | |
| | | daptomycin (2003), | | |
| | | gemifloxacin (2003), | | |
| | | doripenem (2005), | | |
| | | tigecycline (2005), | | |
| | | garenoxacin (2007), | | |
| | | ceftobiprole (2008), | | |

| | | | | |
|---|---|---|---|---|
| | | sitafloxacin (2008), | | |
| | | tebipenem (2009), | | |
| | | telavancin (2009), | | |
| | | antofloxacin (2010), | | |
| | | ceftaroline (2011), | | |
| | | ceftolozane/tazobactam (2014), | | |
| | | dalbavancin (2014), | | |
| | | oritavancin (2014), | | |
| | | tedizolid (2014) | | |
| | TotalDDDPer1000Persons | Total Per Capita Use | ResistanceMap | DDD per 1000 |
| **RESISTANCE: CURRENT STATE OF ANTBIOTIC RESISTANCE** | | | | |
| **TIER 1** | **TIER 2** | **DESCRIPTION** | | **UNIT** |
| **Resistance (Total)** | MRSA | Methicillin-resistant Staphylococcus aureus | | pct (%) |
| | CR | Carbapenems resistance in Enterobacteriaceae | | pct (%) |
| | STR | Streptococcal resistance to macrolides and penicillin (average) | | pct (%) |
| | **DRUG RESISTANCE INDEX (DRI): PATHOGEN-ANTIBIOTIC COMBINATIONS** | | | |
| **TIER 1** | **PATHOGEN** | **ANTIBIOTICS** | | **UNIT** |
| **DRI** | *Enterococcus faecalis/faecium* | Aminopenicillins | | Unit Free |
| | *Escherichia coli* | Aminoglycosides | | Unit Free |
| | | Aminopenicillins | | |
| | | Carbapenems | | |
| | | Cephalosporins (3rd gen) | | |
| | | Fluoroquinolones | | |
| | *Klebsiella pneumonia* | Aminoglycosides | | Unit Free |
| | | Carbapenems | | |
| | | Cephalosporins (3rd gen) | | |
| | | Fluoroquinolones | | |
| | *Pseudomonas aeruginosa* | Aminoglycosides | | Unit Free |
| | | Carbapenems | | |
| | | Cephalosporins (3rd gen) | | |
| | | Fluoroquinolones | | |
| | | Piperacillin-tazobactam | | |
| | *Staphylococcus aureus* | Oxacillin/Cefoxitin (MRSA) | | Unit Free |
| | **ECOLOGICAL VARIABLES** | | | |
| **CONTEXT** | **NAME** | **DESCRIPTION** | **DATABASE** | **UNIT** |

| | | | | |
|---|---|---|---|---|
| Economy | **GDP** | GDP per capita (log transformed) | EORA | Unit Free |
| | **Gini** | Gini index | Inequality – proportion of the lowest 20 % of the national income distribution. | Unit Free |
| Climate | **Mean Temperature** | Mean temperature by population density of grid cells (weighted) | SEDAC, BIOCLIM | degree C |
| Livestock | **Animal Production** | Animal production in mass per country area | GLW | Unit Free |
| Population | **Population Density** | Human population density (log transformed) | SEDAC | $1/km^2$ |

**S3 Table. Governance Syndrome questions.**
Questions used and their sectors included for the calculation of governance syndrome for the years between 2016-2022 according to TrACSS (1.1) 2016-2017 version. All surveys are available online (https://amrcountryprogress.org/#/download-responses). Only questions asked in all surveys were selected for the analysis. In case a question is separated in two questions, they were still kept for the analysis. Answers converted from the scale of A-E to 0-4. Categories' scores were averaged from their constituted questions, and the overall governance score was calculated as the mean score of all categories. NAP stands for National Action Plan.

| ACTION SUB-CATEGORIES | | AMR governance response survey | | | | | | |
|---|---|---|---|---|---|---|---|---|
| **Category** | **Question title** | 2016–2017 | 2017–2018 | 2018–2019 | 2019–2020 | 2020–2021 | 2021–2022 | 2022–2023 |
| **General** | Multi-sector and One Health working arrangements | 4.1 | 4.1 | 4.1 | 4.1 | 4.1 | 2.1 | 2.1 |
| | Country progress with development of a national action plan on AMR | 5.1 | 5.1 | 5.1 | 5.1 | 5.1 | 2.3 | 2.3 |
| **Awareness and Education** | Raising awareness and understanding of AMR risks and response | 6.1, 6.2 | 6.1, 6.2 | 6.1 | 6.1 | 6.1 | 2.9 | 2.9 |
| | Training and professional education on AMR in the human health sector | 6.3 | 6.3 | 6.2 | 6.2 | 6.2 | 3.1 | 3.1 |
| | Training and professional education on AMR in the veterinary sector | 6.4 | 6.4 | 6.3 | 6.3 | 6.3 | 4.1 | 4.1 |
| | Progress with strengthening veterinary services | 6.5 | 6.5 | 6.5 | 6.5 | 6.5 | 4.3 | 4.3 |
| **Monitoring and Surveillance** | National monitoring system for consumption and rational use of antimicrobials in human health | 7.1 | 7.1 | 7.1 | 7.1 | 7.1 | 3.2 | 3.2 |
| | National surveillance system for antimicrobial resistance (AMR) in humans | 7.3 | 7.4 | 7.4 | 7.4 | 7.4 | 3.3 | 3.3 |
| **Prevention** | Infection Prevention and Control (IPC) in human health care | 8.1 | 8.1 | 8.1 | 8.1 | 8.1 | 3.5 | 3.5 |
| | Good health, management and hygiene practices to reduce the use of antimicrobials and minimize development and transmission of AMR in animal production (terrestrial and aquatic) | 8.2 | 8.2 | 8.2 | 8.2 | 8.2 | 4.9, 4.10 | 4.9, 4.10 |
| **Regulation** | Antimicrobial Stewardship & regulation in human health | 9.1 | 9.1 | 9.1 | 9.1 | 9.1 | 3.6 | 3.6 |
| | Antimicrobial stewardship & regulation in terrestrial animal health | 9.2 | 9.2 | 9.2 | 9.2 | 9.2 | 4.11 | 4.11 |

**S4 Table. Questions used for calculating the action index.**

Topics and titles of questions from the TrACSS survey in 2016-17 used to calculate the action index as well as the sub-categories used to group questions within a similar theme. Answers were answered on a scale of A-E and converted to 0-4.

| Topic | Survey Questions |
|---|---|
| **Awareness and Education** | |
| AMR Training (Human Health) * | 6.3 Training and professional education on AMR in the human health sector |
| AMR Training (Animal Health and Food Production) * | 6.4 Training and professional education on AMR in the veterinary sector |
| AMR Awareness (Human Health) * | 6.1 Raising awareness and understanding of AMR risks and response in human health |
| AMR Awareness (Animal Health and Food Production) * | 6.2 Raising Awareness and understanding of AMR risks and response in animal health and food production. |
| **Monitoring and Surveillance** | |
| Monitoring System for AMU (Animals and Crop) * | 7.2 National monitoring system for antimicrobial use in animals and crop production |
| AMR Surveillance System (Humans)* | 7.3 National surveillance system for antimicrobial resistance (AMR) in humans |
| Monitoring System for AMU (Human Health) * | 7.1 National monitoring system for consumption and rational use of antimicrobials in human health |
| AMR Surveillance System (Animals and Foods) * | 7.4 National surveillance system for antimicrobial resistance (AMR) in animals and foods |
| **General** | |
| Veterinary Services | 6.5 Progress with strengthening veterinary services |
| One Health Arrangements | 4.1 Multi-sector and One Health working arrangement |
| NAP progress | 5.1 Country progress with development of a national action plan on AMR |
| **Regulation** | |
| AMS and Regulation (Human)* | 9.1 Antimicrobial Stewardship & regulation in human health |
| AMS and Regulation (Animal & Crop) * | 9.2 Antimicrobial stewardship & regulation in animal and crop production |
| Contamination Prevention | 9.3 Legislation and/or regulations to prevent contamination of the environment with antimicrobials |
| **Prevention** | |
| IPC | 8.1 Infection Prevention and Control (IPC) in human health care |
| AMU Prevention | 8.2 Good animal health and management practices and good hygiene to prevent infections in order to reduce the use of antimicrobials in animals and AMR transmission in food production |
| **Subcategories included in Animal Data Analysis and Figure 4* | |

**S5 Table. Model Formulas for Association between Action and Indicator Linear Trend and Categorical Trend**

Formulas for generalized linear mixed models to investigate the association between linear trend of indicators in 16 years and action. First type of model included linear trend of indicators as response variable. The second type of model included Action as a response variable and categorical trend as an explanatory variable to investigate the action difference between the countries with positive vs negative change. Results shown in Figure 2 for tier 1 indicators.

| Indicators | Formula |
|---|---|
| **Drivers** | Linear trend ~ Action + Baseline + (1\|Income) |
| | Action ~ Categorical trend + Baseline + (1\|Income) |
| **Use** | Linear trend ~ Action + Baseline + (1\|Income) |
| | Action ~ Categorical trend + Baseline + (1\|Income) |
| **Resistance** | Linear trend ~ Action + Baseline + (1\|Income) |
| | Action ~ Categorical trend + Baseline + (1\|Income) |
| **DRI** | Linear trend ~ Action + Baseline + (1\|Income) |
| | Action ~ Categorical trend + Baseline + (1\|Income) |

**S6 Table. De-escalation plot formulas for univariate models.**

Formulas used for binomial general linear model to investigate the association between de-escalation of categories and action level. Declining proportion refers to declining numbers of tier 2 indicators divided by the total number of tier 2 indicators for countries, Baseline Mean refers to mean of baseline for tier 2 indicators for each DPSE indicator (See S2 Table). Models weighted by total number of tier 2 indicators within each DPSE indicator for specific country.

| **Indicators** | **Formula** | **Weight** |
|---|---|---|
| **Drivers** | Declining proportion ~ Action+ Baseline Mean | (1-4)/4 |
| **Use** | Declining proportion ~ Action+ Baseline Mean | (1-3)/3 |
| **Resistance** | Declining proportion ~ Action+ Baseline Mean | (1-3)/3 |
| **DRI** | Declining proportion ~ Action+ Baseline Mean | 1/1 |

**S7 Table. Global Models Data Subset Formulas for The Model Selection**
Global Models for multivariate model selection data subsets.

| Model Name | Description |
|---|---|
| DPSEA | All DPSE indicators as response variables. |
| DPSEA.noDr | All DPSE indicators as response variables, explanatory variables relating to health system excluded. |
| aP.noDr | Only countries reporting use (P) included, all DPSE indicators as response variable, explanatory variables relating to health system excluded. |
| aS.noDr | Only countries reporting resistance (S) included, all DPSE indicators as response variable, explanatory variables relating to health system excluded. |
| aE.noDr | Only countries reporting DRI (E) included, all DPSE indicators as response variable, explanatory variables relating to health system excluded. |
| aP | Only countries reporting use (P) included, all DPSE indicators as response variable. |
| aS | Only countries reporting resistance (S) included, all DPSE indicators as response variable. |
| aE | Only countries reporting DRI (E), all DPSE indicators as response variable. |
| Dr | Only countries with tier 2 driver components and at least one of Use (P), Resistance (S) or DRI (E) included. Only driver indicators as response variable. |
| P | Only countries with tier 2 use (P) components included. Only use indicators as response variable. |
| S | Only countries with tier 2 resistance (S) components included. Only resistance indicators as response variable. |
| E | Only countries with DRI (E) included. Only DRI indicators as response variable. |
| DPS | Only countries with reporting driver, use (P), or resistance (S) included. Driver, use, and resistance as a response variable included. |
| PSE | Only countries with reporting use (P), resistance (S) or DRI (E) included. use, resistance, and DRI as a response variable included. |
| DP | Only countries with reporting driver or use (P) included. Driver and use as a response variable included. |
| PS | Only countries with reporting, use (P), or resistance (S) included. Use and resistance as a response variable included. |
| SE | Only countries with reporting resistance (S) or DRI (E) included. Resistance and DRI as a response variable included. |

**S8 Table. Model selection global model formulas**

Global models as starting points for creating and evaluating model subsets.

| Model Name | Global Models with DPSE interaction | Global Models with income interaction |
|---|---|---|
| DPSEA<br>aP<br>aS<br>aE<br>DPS<br>PSE<br>DP<br>PS<br>SE | Linear trend ~ Action * DPSE + General * DPSE + Monitoring and Surveillance * DPSE + Awareness and Education * DPSE + Sanitation * DPSE + Infection * DPSE + Workforce * DPSE + Vaccination * DPSE + GDP * DPSE + Pop. density * DPSE + Gini * DPSE + Animal prod. * DPSE + Mean temperature * DPSE + Baseline * DPSE + (1 \| ISO3) | Linear trend ~ Action * income + General * income + Monitoring and Surveillance * income + Awareness and Education * income + Sanitation * income + Infection * income + Workforce * income + Vaccination * income + GDP * income + Pop. density * income + Gini * income + Animal prod. * income + Mean temperature * income + Baseline * income + (1 \| ISO3) |
| DPSEA.noDr<br>aP.noDr<br>aS.noDr<br>aE.noDr | Linear trend ~ Action * DPSE + General * DPSE + Monitoring and Surveillance * DPSE + Awareness and Education * DPSE + GDP * DPSE + Pop. density * DPSE + Gini * DPSE + Animal prod. * DPSE + Mean temperature * DPSE + Baseline * DPSE + (1 \| ISO3) | Linear trend ~ Action * income + General * income + Monitoring and Surveillance * income + Awareness and Education * income + GDP * income + Pop. density * income + Gini * income + Animal prod. * income + Mean temperature * income + Baseline * income + (1 \| ISO3) |
| Dr | No model | Linear trend ~ Action*income + General*income + Monitoring and Surveillance*income + Awareness and Education*income + GDP*income + Pop. density*income + Gini*income + Animal prod.*income + Mean temperature*income + Baseline*income + (1\|ISO3) + (1\|Subcategory) |
| P<br>S | No model | Linear trend ~ Action*income + General*income + Monitoring and Surveillance*income + Awareness and Education*income + Sanitation * income + Infection * income + Workforce * income + Vaccination * income + GDP*income + Pop. density*income + Gini*income + Animal prod.*income + Mean temperature*income + Baseline*income + (1\|ISO3) + (1\|Subcategory) |
| E | No model | Linear trend ~ Action * income + General * income + Monitoring and Surveillance * income + Awareness and Education * income + Sanitation * income + Infection * income + Workforce * income + Vaccination * income + GDP * income + Pop. density * income + Gini * income + Animal prod. * income + Mean temperature * income + Baseline * income |
| **HIC** | | |
| DPSEA<br>aP<br>aS<br>aE<br>DPS<br>PSE<br>DP<br>PS<br>SE | Linear trend ~ Action * DPSE + General * DPSE + Monitoring and Surveillance * DPSE + Awareness and Education * DPSE + Sanitation * DPSE + Infection * DPSE + Workforce * DPSE + Vaccination * DPSE + GDP * DPSE + Pop. density * DPSE + Gini * DPSE + Animal prod. * DPSE + Mean temperature * DPSE + Baseline * DPSE + (1 \| ISO3) | No model |

| Model Name | Global Models with DPSE interaction | Global Models with income interaction |
|---|---|---|
| DPSEA.noDr<br>aP.noDr<br>aS.noDr<br>aE.noDr | Linear trend ~ Action * DPSE + General * DPSE + Monitoring and Surveillance * DPSE + Awareness and Education * DPSE + GDP * DPSE + Pop. density * DPSE + Gini * DPSE + Animal prod. * DPSE + Mean temperature * DPSE + Baseline * DPSE + (1 \| ISO3) | No model |
| Dr | Linear trend ~ Action + General + Monitoring and Surveillance + Awareness and Education + GDP + Pop. density + Gini + Animal prod. + Mean temperature + Baseline + (1 \| ISO3) + (1 \| Subcategory) | No model |
| P<br>S<br>E | Linear trend ~ Action + General + Monitoring and Surveillance + Awareness and Education + Sanitation + Infection + Workforce + Vaccination + GDP + Pop. density + Gini + Animal prod. + Mean temperature + Baseline + (1 \| ISO3) + (1 \| Subcategory) | No model |
| **LMIC** | | No model |
| DPSEA<br>aP<br>aS<br>aE<br>DPS<br>PSE<br>DP<br>PS | Linear trend ~ Action * DPSE + General * DPSE + Monitoring and Surveillance * DPSE + Awareness and Education * DPSE + Sanitation * DPSE + Infection * DPSE + Workforce * DPSE + Vaccination * DPSE + GDP * DPSE + Pop. density * DPSE + Gini * DPSE + Animal prod. * DPSE + Mean temperature * DPSE + Baseline * DPSE + (1 \| ISO3) | No model |
| DPSEA.noDr<br>aP.noDr<br>aS.noDr<br>aE.noDr | Linear trend ~ Action * DPSE + General * DPSE + Monitoring and Surveillance * DPSE + Awareness and Education * DPSE + GDP * DPSE + Pop. density * DPSE + Gini * DPSE + Animal prod. * DPSE + Mean temperature * DPSE + Baseline * DPSE + (1 \| ISO3) | No model |
| Dr | Linear trend ~ Action + General + Monitoring and Surveillance + Awareness and Education + GDP + Pop. density + Gini + Animal prod. + Mean temperature + Baseline + (1 \| ISO3) + (1 \| Subcategory) | No model |
| P | Linear trend ~ Action + General + Monitoring and Surveillance + Awareness and Education + Sanitation + Infection + Workforce + Vaccination + GDP + Pop. density + Gini + Animal prod. + Mean temperature + Baseline + (1 \| ISO3) + (1 \| Subcategory) | No model |
| E | No model | No model |

| Model Name | Global Models with DPSE interaction | Global Models with income interaction |
|---|---|---|
| SE | No model | No model |
| **BINOMIAL** | | |
| DPSEA<br>aP<br>aS<br>aE | Categorical trend ~ Action * DPSE + General * DPSE + Monitoring and Surveillance * DPSE + Awareness and Education * DPSE + Sanitation * DPSE + Infection * DPSE + Workforce * DPSE + Vaccination * DPSE + GDP * DPSE + Pop. density * DPSE + Gini * DPSE + Animal prod. * DPSE + Mean temperature * DPSE + Baseline * DPSE + (1 \| ISO3) | Categorical trend ~ Action * income + General * income + Monitoring and Surveillance * income + Awareness and Education * income + Sanitation * income + Infection * income + Workforce * income + Vaccination * income + GDP * income + Pop. density * income + Gini * income + Animal prod. * income + Mean temperature * income + Baseline * income + (1 \| ISO3) |
| DPSEA.noDr<br>aP.noDr<br>aE.noDr | Categorical trend ~ Action * DPSE + General * DPSE + Monitoring and Surveillance * DPSE + Awareness and Education * DPSE + GDP * DPSE + Pop. density * DPSE + Gini * DPSE + Animal prod. * DPSE + Mean temperature * DPSE + Baseline * DPSE + (1 \| ISO3) | Categorical trend ~ Action * income + General * income + Monitoring and Surveillance * income + Awareness and Education * income + GDP * income + Pop. density * income + Gini * income + Animal prod. * income + Mean temperature * income + Baseline * income + (1 \| ISO3) |
| aS.noDr | Categorical trend ~ Action * DPSE + General * DPSE + Monitoring and Surveillance * DPSE + Awareness and Education * DPSE + GDP * DPSE + Pop. density * DPSE + Gini * DPSE + Animal prod. * DPSE + Baseline * DPSE + (1 \| ISO3); Categorical trend ~ Action * DPSE + General * DPSE + Monitoring and Surveillance * DPSE + Awareness and Education * DPSE + GDP * DPSE + Pop. density * DPSE + Gini * DPSE + Mean temperature * DPSE + Baseline * DPSE + (1 \| ISO3); Categorical trend ~ Action * DPSE + General * DPSE + Monitoring and Surveillance * DPSE + Awareness and Education * DPSE + GDP * DPSE + Gini * DPSE + Animal prod. * DPSE + Mean temperature * DPSE + Baseline * DPSE + (1 \| ISO3) | Categorical trend ~ Action * income + General * income + Monitoring and Surveillance * income + Awareness and Education * income + GDP * income + Pop. density * income + Gini * income + Animal prod. * income + Mean temperature * income + Baseline * income + (1 \| ISO3) |
| Dr | | Categorical trend ~ Action * income + General * income + Monitoring and Surveillance * income + Awareness and Education * income + GDP * income + Pop. density * income + Gini * income + Animal prod. * income + Mean temperature * income + Baseline * income + (1 \| ISO3) |
| P<br>S | | Categorical trend ~ Action * income + General * income + Monitoring and Surveillance * income + Awareness and Education * income + Sanitation * income + Infection * income + Workforce * income + Vaccination * income + GDP * income + Pop. density * income + Gini * income + Animal prod. * income + Mean temperature * income + Baseline * income + (1 \| ISO3) + (1 \| Subcategory) |

| Model Name | Global Models with DPSE interaction | Global Models with income interaction |
|---|---|---|
| E | No model | No model |
| DPS<br>PSE<br>DP<br>SE | Categorical trend ~ Action * DPSE + General * DPSE + Monitoring and Surveillance * DPSE + Awareness and Education * DPSE + Sanitation * DPSE + Infection * DPSE + Workforce * DPSE + Vaccination * DPSE + GDP * DPSE + Pop. density * DPSE + Gini * DPSE + Animal prod. * DPSE + Mean temperature * DPSE + Baseline * DPSE + (1 \| ISO3) | Categorical trend ~ Action * income + General * income + Monitoring and Surveillance * income + Awareness and Education * income + Sanitation * income + Infection * income + Workforce * income + Vaccination * income + GDP * income + Pop. density * income + Gini * income + Animal prod. * income + Mean temperature * income + Baseline * income + (1 \| ISO3) |
| PS | Categorical trend ~ Action * DPSE + General * DPSE + Monitoring and Surveillance * DPSE + Awareness and Education * DPSE + Sanitation * DPSE + Infection * DPSE + Workforce * DPSE + Vaccination * DPSE + GDP * DPSE + Pop. density * DPSE + Gini * DPSE + Animal prod. * DPSE + Mean temperature * DPSE + Baseline * DPSE + (1 \| ISO3) | Categorical trend ~ Action * income + General * income + Monitoring and Surveillance * income + Awareness and Education * income + Sanitation * income + Infection * income + Workforce * income + Vaccination * income + Pop. density * income + Gini * income + Animal prod. * income + Mean temperature * income + Baseline * income + (1 \| ISO3)**;** Categorical trend ~ General * income + Monitoring and Surveillance * income + Awareness and Education * income + Sanitation * income + Infection * income + Workforce * income + Vaccination * income + GDP * income + Pop. density * income + Gini * income + Animal prod. * income + Mean temperature * income + Baseline * income + (1 \| ISO3)**;** Categorical trend ~ Action * income + Monitoring and Surveillance * income + Awareness and Education * income + Sanitation * income + Infection * income + Workforce * income + Vaccination * income + GDP * income + Pop. density * income + Gini * income + Animal prod. * income + Mean temperature * income + Baseline * income + (1 \| ISO3) |

**S9 Table. Association Between Linear Trend and Action**

Association is determined with linear mixed model between linear trend of variables as a response variable and action and baseline as fixed effects and income level as a random effect.

| Indicators | DPSE | Coefficient | t-value | std.error | df | p.value | Number of Countries with Increase | Sample Size |
|---|---|---|---|---|---|---|---|---|
| level 1 | | | | | | | | |
| Drivers Total | Drivers | -0.05 | -2.6 | 0.02 | 21.4 | **0.017** | 6 | 73 |
| Use Total | Use | -0.16 | -2.3 | 0.07 | 39.0 | **0.024** | 55 | 65 |
| Resistance Total | Resistance | -0.11 | -1.1 | 0.10 | 26.3 | 0.291 | 16 | 32 |
| DRI | DRI | -0.17 | -2.1 | 0.08 | 21.1 | **0.044** | 21 | 25 |
| level 2 | | | | | | | | |
| Infections | Drivers | 0.01 | 0.8 | 0.01 | 48.1 | 0.402 | 12 | 73 |
| Sanitation | Drivers | -0.01 | -0.6 | 0.02 | 66.1 | 0.548 | 27 | 73 |
| Vaccination | Drivers | -0.09 | -1.6 | 0.05 | 70.0 | 0.108 | 11 | 73 |
| Workforce | Drivers | -0.21 | -3.9 | 0.05 | 29.8 | **0.001** | 9 | 55 |
| TotalDDDPer1000Persons | Use | 0.04 | 0.4 | 0.10 | 61.4 | 0.659 | 50 | 65 |
| BroadPerTotalABXUse | Use | -0.32 | -3.3 | 0.10 | 43.7 | **0.002** | 47 | 65 |
| NewABXUse | Use | -0.15 | -1.8 | 0.08 | 60.0 | 0.076 | 55 | 63 |
| MRSA | Resistance | -0.17 | -1.6 | 0.11 | 29.0 | 0.129 | 11 | 32 |
| CR | Resistance | -0.11 | -0.6 | 0.20 | 18.7 | 0.587 | 20 | 28 |
| STR | Resistance | -0.13 | -1.7 | 0.08 | 22.0 | 0.113 | 13 | 25 |
| level 3 | | | | | | | | |
| HIV | Drivers/infections | 0.01 | 1.6 | 0.01 | 28.0 | 0.127 | 22 | 31 |
| TB | Drivers/infections | 0.02 | 1.1 | 0.02 | 27.9 | 0.29 | 11 | 73 |
| Drinking Water Source | Drivers/Sanitation | 0.01 | 0.6 | 0.02 | 69.0 | 0.577 | 65 | 72 |
| Water Source Access | Drivers/Sanitation | 0.01 | 0.6 | 0.02 | 69.0 | 0.533 | 65 | 72 |
| Overall Sanitation | Drivers/Sanitation | 0.01 | 0.8 | 0.02 | 62.4 | 0.435 | 63 | 66 |
| DTP3 | Drivers/Vaccination | 0.11 | 1.8 | 0.06 | 69.0 | 0.081 | 51 | 72 |
| HepB3 | Drivers/Vaccination | 0.13 | 1.5 | 0.09 | 57.0 | 0.148 | 48 | 60 |
| Hib3 | Drivers/Vaccination | 0.02 | 0.3 | 0.08 | 41.9 | 0.796 | 45 | 53 |
| Pol3 | Drivers/Vaccination | 0.13 | 2.3 | 0.06 | 69.0 | **0.027** | 49 | 72 |
| Measles | Drivers/Vaccination | 0.11 | 2.3 | 0.05 | 70.0 | **0.022** | 53 | 73 |
| RCV1 | Drivers/Vaccination | 0.18 | 2.2 | 0.08 | 59.0 | **0.033** | 43 | 62 |
| Nursing | Drivers/Workforce | 0.19 | 2.6 | 0.07 | 39.0 | **0.014** | 35 | 42 |
| Physicians | Drivers/Workforce | 0.19 | 3.6 | 0.05 | 52.0 | **0.001** | 44 | 55 |

lmer(Linear trend ~ Action + Baseline + (1|income))

#### S10 Table. Association Between Categorical Trend and Mean Action Score

| Indicators | DPSE | Coefficient | t-value | std.error | df | p.value | Number of Countries with Increase | Sample Size |
|---|---|---|---|---|---|---|---|---|
| level 1 | | | | | | | | |
| Drivers Total | Drivers | -0.37 | -1.4 | 0.27 | 69.1 | 0.169 | 6 | 73 |
| Use Total | Use | -0.65 | -3.2 | 0.21 | 61.0 | **0.002** | 55 | 65 |
| Resistance Total | Resistance | -0.69 | -2.9 | 0.24 | 29.0 | **0.006** | 16 | 32 |
| DRI | DRI | -0.80 | -2.1 | 0.38 | 22.0 | **0.047** | 21 | 25 |
| level 2 | | | | | | | | |
| Infections | Drivers | 0.18 | 0.9 | 0.20 | 69.2 | 0.364 | 12 | 73 |
| Sanitation | Drivers | 0.24 | 1.4 | 0.17 | 69.0 | 0.156 | 27 | 73 |
| Vaccination | Drivers | -0.08 | -0.4 | 0.21 | 69.1 | 0.693 | 11 | 73 |
| Workforce | Drivers | -0.59 | -2.8 | 0.21 | 51.2 | **0.007** | 9 | 55 |
| TotalDDDPer1000Persons | Use | -0.01 | -0.1 | 0.21 | 61.1 | 0.951 | 50 | 65 |
| BroadPerTotalABXUse | Use | -0.55 | -3.3 | 0.17 | 61.0 | **0.001** | 47 | 65 |
| NewABXUse | Use | -0.60 | -2.4 | 0.25 | 59.0 | **0.02** | 55 | 63 |
| MRSA | Resistance | 0.01 | 0.0 | 0.32 | 28.5 | 0.981 | 11 | 32 |
| CR | Resistance | -0.38 | -1.1 | 0.35 | 24.8 | 0.279 | 20 | 28 |
| STR | Resistance | -0.62 | -2.1 | 0.30 | 21.9 | **0.049** | 13 | 25 |
| level 3 | | | | | | | | |
| HIV | Drivers/infections | -0.16 | -0.7 | 0.23 | 27.0 | 0.486 | 22 | 31 |
| TB | Drivers/infections | 0.19 | 0.9 | 0.21 | 69.1 | 0.351 | 11 | 73 |
| Drinking Water Source | Drivers/Sanitation | 0.53 | 2.1 | 0.25 | 68.8 | **0.038** | 65 | 72 |
| Water Source Access | Drivers/Sanitation | 0.55 | 2.2 | 0.25 | 68.8 | **0.033** | 65 | 72 |
| Overall Sanitation | Drivers/Sanitation | -0.29 | -0.7 | 0.39 | 62.5 | 0.459 | 63 | 66 |
| DTP3 | Drivers/Vaccination | 0.00 | 0.0 | 0.17 | 68.1 | 0.978 | 51 | 72 |
| HepB3 | Drivers/Vaccination | 0.14 | 0.7 | 0.20 | 56.0 | 0.492 | 48 | 60 |
| Hib3 | Drivers/Vaccination | -0.26 | -1.0 | 0.25 | 49.2 | 0.306 | 45 | 53 |
| Pol3 | Drivers/Vaccination | 0.25 | 1.5 | 0.16 | 68.0 | 0.133 | 49 | 72 |
| Measles | Drivers/Vaccination | 0.11 | 0.7 | 0.17 | 69.1 | 0.503 | 53 | 73 |
| RCV1 | Drivers/Vaccination | 0.15 | 0.8 | 0.19 | 58.3 | 0.428 | 43 | 62 |
| Nursing | Drivers/Workforce | 0.85 | 4.0 | 0.21 | 38.0 | **< .001** | 35 | 42 |
| Physicians | Drivers/Workforce | 0.54 | 2.7 | 0.20 | 51.0 | **0.008** | 44 | 55 |

lmer(Action ~ Sign + Baseline + (1|income))

**S11 Table. Association Between Baseline and Action Score.**
Action Score calculated from the mean of all action categories (See S3 Table). Association is determined with linear mixed model between baseline values for variables as a response variable and action as a fixed effect and income level as a random effect.

| Indicators | DPSE | Coefficient | t-value | std.error | df | p.value | Number of Countries with Increase | Sample Size |
|---|---|---|---|---|---|---|---|---|
| level 1 | | | | | | | | |
| Drivers Total | Drivers | 0.12 | 1.5 | 0.08 | 67.6 | 0.131 | 6 | 73 |
| Use Total | Use | 0.00 | 0.0 | 0.13 | 63.0 | 0.982 | 55 | 65 |
| Resistance Total | Resistance | -0.16 | -1.1 | 0.14 | 30.0 | 0.265 | 16 | 32 |
| DRI | DRI | -0.30 | -1.6 | 0.19 | 22.9 | 0.126 | 21 | 25 |
| level 2 | | | | | | | | |
| Infections | Drivers | 0.02 | 0.2 | 0.09 | 69.8 | 0.835 | 12 | 73 |
| Sanitation | Drivers | 0.09 | 1.2 | 0.08 | 71.0 | 0.244 | 27 | 73 |
| Vaccination | Drivers | 0.18 | 1.3 | 0.14 | 41.1 | 0.216 | 11 | 73 |
| Workforce | Drivers | 0.19 | 1.4 | 0.14 | 52.8 | 0.182 | 9 | 55 |
| TotalDDDPer1000Persons | Use | -0.12 | -0.8 | 0.15 | 63.0 | 0.442 | 50 | 65 |
| BroadPerTotalABXUse | Use | 0.03 | 0.2 | 0.17 | 62.4 | 0.859 | 47 | 65 |
| NewABXUse | Use | 0.14 | 0.8 | 0.17 | 57.0 | 0.405 | 55 | 63 |
| MRSA | Resistance | -0.21 | -0.8 | 0.26 | 26.4 | 0.423 | 11 | 32 |
| CR | Resistance | -0.06 | -0.6 | 0.11 | 26.0 | 0.569 | 20 | 28 |
| STR | Resistance | -0.13 | -0.6 | 0.20 | 23.0 | 0.529 | 13 | 25 |
| level 3 | | | | | | | | |
| HIV | Drivers/infections | 0.03 | 0.1 | 0.19 | 29.0 | 0.887 | 22 | 31 |
| TB | Drivers/infections | -0.01 | -0.1 | 0.11 | 70.8 | 0.919 | 11 | 73 |
| Drinking Water Source | Drivers/Sanitation | 0.04 | 0.7 | 0.06 | 69.8 | 0.46 | 65 | 72 |
| Water Source Access | Drivers/Sanitation | 0.07 | 1.2 | 0.06 | 69.9 | 0.25 | 65 | 72 |
| Overall Sanitation | Drivers/Sanitation | 0.10 | 1.0 | 0.10 | 64.0 | 0.323 | 63 | 66 |
| DTP3 | Drivers/Vaccination | 0.06 | 0.6 | 0.09 | 56.8 | 0.529 | 51 | 72 |
| HepB3 | Drivers/Vaccination | 0.06 | 0.3 | 0.23 | 58.0 | 0.802 | 48 | 60 |
| Hib3 | Drivers/Vaccination | 0.43 | 2.6 | 0.17 | 11.3 | **0.026** | 45 | 53 |
| Pol3 | Drivers/Vaccination | 0.07 | 0.7 | 0.10 | 51.4 | 0.494 | 49 | 72 |
| Measles | Drivers/Vaccination | 0.09 | 1.0 | 0.09 | 71.0 | 0.335 | 53 | 73 |
| RCV1 | Drivers/Vaccination | -0.03 | -0.2 | 0.14 | 60.0 | 0.82 | 43 | 62 |
| Nursing | Drivers/Workforce | 0.27 | 1.4 | 0.19 | 39.7 | 0.163 | 35 | 42 |
| Physicians | Drivers/Workforce | 0.18 | 1.1 | 0.16 | 52.9 | 0.273 | 44 | 55 |

lmer(Baseline ~ Action + (1|income))

#### S12 Table. Linear Trend and Awareness and Education

| Indicators | DPSE | Coefficient | t-value | std.error | df | p.value | Number of Countries with Increase | Sample Size |
|---|---|---|---|---|---|---|---|---|
| level 1 | | | | | | | | |
| Drivers Total | Drivers | -0.04 | -2.0 | 0.02 | 70.0 | **0.046** | 6 | 73 |
| Use Total | Use | -0.08 | -1.3 | 0.06 | 62.0 | 0.205 | 55 | 65 |
| Resistance Total | Resistance | -0.04 | -0.4 | 0.09 | 29.0 | 0.666 | 16 | 32 |
| DRI | DRI | -0.12 | -1.7 | 0.07 | 21.2 | 0.113 | 21 | 25 |
| level 2 | | | | | | | | |
| Infections | Drivers | 0.01 | 0.5 | 0.01 | 69.9 | 0.593 | 12 | 73 |
| Sanitation | Drivers | -0.02 | -1.6 | 0.01 | 69.8 | 0.12 | 27 | 73 |
| Vaccination | Drivers | -0.06 | -1.3 | 0.05 | 70.0 | 0.201 | 11 | 73 |
| Workforce | Drivers | -0.10 | -2.1 | 0.05 | 52.0 | **0.04** | 9 | 55 |
| TotalDDDPer1000Persons | Use | 0.08 | 1.0 | 0.08 | 61.6 | 0.305 | 50 | 65 |
| BroadPerTotalABXUse | Use | -0.17 | -2.1 | 0.08 | 62.0 | **0.039** | 47 | 65 |
| NewABXUse | Use | -0.15 | -2.1 | 0.07 | 60.0 | **0.043** | 55 | 63 |
| MRSA | Resistance | -0.08 | -0.8 | 0.10 | 29.0 | 0.433 | 11 | 32 |
| CR | Resistance | 0.04 | 0.2 | 0.19 | 24.8 | 0.831 | 20 | 28 |
| STR | Resistance | -0.11 | -1.7 | 0.07 | 21.7 | 0.106 | 13 | 25 |
| level 3 | | | | | | | | |
| HIV | Drivers/infections | 0.02 | 2.2 | 0.01 | 28.0 | **0.034** | 22 | 31 |
| TB | Drivers/infections | 0.00 | 0.2 | 0.02 | 69.3 | 0.82 | 11 | 73 |
| Drinking Water Source | Drivers/Sanitation | 0.02 | 1.4 | 0.02 | 69.0 | 0.181 | 65 | 72 |
| Water Source Access | Drivers/Sanitation | 0.02 | 1.4 | 0.01 | 69.0 | 0.168 | 65 | 72 |
| Overall Sanitation | Drivers/Sanitation | 0.02 | 1.2 | 0.01 | 62.6 | 0.234 | 63 | 66 |
| DTP3 | Drivers/Vaccination | 0.07 | 1.2 | 0.06 | 54.9 | 0.251 | 51 | 72 |
| HepB3 | Drivers/Vaccination | 0.11 | 1.5 | 0.07 | 57.0 | 0.151 | 48 | 60 |
| Hib3 | Drivers/Vaccination | 0.07 | 1.2 | 0.06 | 47.6 | 0.245 | 45 | 53 |
| Pol3 | Drivers/Vaccination | 0.08 | 1.6 | 0.05 | 47.3 | 0.124 | 49 | 72 |
| Measles | Drivers/Vaccination | 0.08 | 1.8 | 0.05 | 70.0 | 0.082 | 53 | 73 |
| RCV1 | Drivers/Vaccination | 0.13 | 1.7 | 0.07 | 59.0 | 0.094 | 43 | 62 |
| Nursing | Drivers/Workforce | 0.06 | 1.0 | 0.06 | 39.0 | 0.307 | 35 | 42 |
| Physicians | Drivers/Workforce | 0.11 | 2.4 | 0.05 | 52.0 | **0.019** | 44 | 55 |

lmer(Linear Trend ~ Awareness and Education + Baseline + (1|income))

#### S13 Table. Categorical Trend and Awareness and Education

| Indicators | DPSE | Coefficient | t-value | std.error | df | p.value | Number of Countries with Increase | Sample Size |
|---|---|---|---|---|---|---|---|---|
| level 1 | | | | | | | | |
| Drivers Total | Drivers | -0.43 | -1.3 | 0.34 | 69.3 | 0.211 | 6 | 73 |
| Use Total | Use | -0.66 | -2.6 | 0.26 | 61.0 | **0.013** | 55 | 65 |
| Resistance Total | Resistance | -0.66 | -2.4 | 0.27 | 29.0 | **0.022** | 16 | 32 |
| DRI | DRI | -0.79 | -1.8 | 0.43 | 21.9 | 0.08 | 21 | 25 |
| level 2 | | | | | | | | |
| Infections | Drivers | 0.22 | 0.9 | 0.25 | 69.6 | 0.388 | 12 | 73 |
| Sanitation | Drivers | 0.24 | 1.1 | 0.21 | 69.2 | 0.27 | 27 | 73 |
| Vaccination | Drivers | -0.27 | -1.0 | 0.26 | 69.2 | 0.312 | 11 | 73 |
| Workforce | Drivers | -0.62 | -2.2 | 0.29 | 51.8 | **0.033** | 9 | 55 |
| TotalDDDPer1000Persons | Use | 0.15 | 0.6 | 0.25 | 61.5 | 0.563 | 50 | 65 |
| BroadPerTotalABXUse | Use | -0.53 | -2.5 | 0.21 | 61.0 | **0.016** | 47 | 65 |
| NewABXUse | Use | -0.63 | -2.1 | 0.31 | 59.0 | **0.045** | 55 | 63 |
| MRSA | Resistance | -0.03 | -0.1 | 0.36 | 28.7 | 0.924 | 11 | 32 |
| CR | Resistance | -0.53 | -1.4 | 0.37 | 25.0 | 0.168 | 20 | 28 |
| STR | Resistance | -0.82 | -2.3 | 0.35 | 21.5 | **0.029** | 13 | 25 |
| level 3 | | | | | | | | |
| HIV | Drivers/infections | 0.14 | 0.4 | 0.34 | 27.0 | 0.681 | 22 | 31 |
| TB | Drivers/infections | 0.19 | 0.8 | 0.26 | 69.3 | 0.453 | 11 | 73 |
| Drinking Water Source | Drivers/Sanitation | 0.78 | 2.6 | 0.30 | 69.0 | **0.011** | 65 | 72 |
| Water Source Access | Drivers/Sanitation | 0.69 | 2.2 | 0.31 | 63.2 | **0.03** | 65 | 72 |
| Overall Sanitation | Drivers/Sanitation | -0.14 | -0.3 | 0.49 | 62.8 | 0.768 | 63 | 66 |
| DTP3 | Drivers/Vaccination | 0.08 | 0.4 | 0.21 | 68.4 | 0.687 | 51 | 72 |
| HepB3 | Drivers/Vaccination | 0.18 | 0.7 | 0.25 | 56.0 | 0.468 | 48 | 60 |
| Hib3 | Drivers/Vaccination | -0.04 | -0.1 | 0.33 | 49.4 | 0.891 | 45 | 53 |
| Pol3 | Drivers/Vaccination | 0.15 | 0.7 | 0.20 | 68.1 | 0.458 | 49 | 72 |
| Measles | Drivers/Vaccination | 0.03 | 0.1 | 0.21 | 69.4 | 0.886 | 53 | 73 |
| RCV1 | Drivers/Vaccination | 0.02 | 0.1 | 0.24 | 58.8 | 0.925 | 43 | 62 |
| Nursing | Drivers/Workforce | 0.89 | 2.9 | 0.30 | 38.0 | **0.006** | 35 | 42 |
| Physicians | Drivers/Workforce | 0.41 | 1.5 | 0.27 | 51.0 | 0.135 | 44 | 55 |

lmer(Awareness and Education ~ Sign + Baseline + (1|income))

#### S14 Table. Linear Trend and General

| Indicators | DPSE | Coefficient | t-value | std.error | df | p.value | Number of Countries with Increase | Sample Size |
|---|---|---|---|---|---|---|---|---|
| level 1 | | | | | | | | |
| Drivers Total | Drivers | -0.03 | -1.8 | 0.02 | 70.0 | 0.078 | 6 | 73 |
| Use Total | Use | -0.09 | -1.9 | 0.05 | 61.7 | 0.066 | 55 | 65 |
| Resistance Total | Resistance | -0.11 | -1.7 | 0.06 | 29.0 | 0.097 | 16 | 32 |
| DRI | DRI | -0.12 | -2.4 | 0.05 | 21.1 | **0.026** | 21 | 25 |
| level 2 | | | | | | | | |
| Infections | Drivers | 0.02 | 2.0 | 0.01 | 62.6 | **0.045** | 12 | 73 |
| Sanitation | Drivers | -0.01 | -0.4 | 0.01 | 69.2 | 0.674 | 27 | 73 |
| Vaccination | Drivers | -0.03 | -0.8 | 0.04 | 38.2 | 0.432 | 11 | 73 |
| Workforce | Drivers | -0.14 | -3.4 | 0.04 | 52.0 | **0.001** | 9 | 55 |
| TotalDDDPer1000Persons | Use | 0.04 | 0.7 | 0.07 | 61.9 | 0.512 | 50 | 65 |
| BroadPerTotalABXUse | Use | -0.19 | -2.9 | 0.07 | 62.0 | **0.005** | 47 | 65 |
| NewABXUse | Use | -0.11 | -1.7 | 0.06 | 60.0 | 0.091 | 55 | 63 |
| MRSA | Resistance | -0.14 | -2.0 | 0.07 | 29.0 | 0.06 | 11 | 32 |
| CR | Resistance | -0.13 | -1.0 | 0.13 | 25.0 | 0.309 | 20 | 28 |
| STR | Resistance | -0.06 | -1.2 | 0.05 | 22.0 | 0.229 | 13 | 25 |
| level 3 | | | | | | | | |
| HIV | Drivers/infections | 0.02 | 2.2 | 0.01 | 28.0 | **0.036** | 22 | 31 |
| TB | Drivers/infections | 0.03 | 1.9 | 0.02 | 43.4 | 0.061 | 11 | 73 |
| Drinking Water Source | Drivers/Sanitation | 0.01 | 0.6 | 0.01 | 69.0 | 0.582 | 65 | 72 |
| Water Source Access | Drivers/Sanitation | 0.00 | 0.4 | 0.01 | 69.0 | 0.713 | 65 | 72 |
| Overall Sanitation | Drivers/Sanitation | 0.01 | 0.8 | 0.01 | 63.0 | 0.413 | 63 | 66 |
| DTP3 | Drivers/Vaccination | 0.05 | 1.0 | 0.05 | 43.1 | 0.334 | 51 | 72 |
| HepB3 | Drivers/Vaccination | 0.10 | 1.4 | 0.07 | 57.0 | 0.168 | 48 | 60 |
| Hib3 | Drivers/Vaccination | -0.07 | -1.2 | 0.06 | 49.8 | 0.228 | 45 | 53 |
| Pol3 | Drivers/Vaccination | 0.07 | 1.4 | 0.05 | 32.5 | 0.163 | 49 | 72 |
| Measles | Drivers/Vaccination | 0.07 | 1.7 | 0.04 | 70.0 | 0.094 | 53 | 73 |
| RCV1 | Drivers/Vaccination | 0.11 | 1.9 | 0.06 | 59.0 | 0.068 | 43 | 62 |
| Nursing | Drivers/Workforce | 0.15 | 2.7 | 0.06 | 39.0 | **0.011** | 35 | 42 |
| Physicians | Drivers/Workforce | 0.13 | 3.1 | 0.04 | 52.0 | **0.003** | 44 | 55 |

lmer(Linear Trend ~ General + Baseline + (1|income))

#### S15 Table. Categorical Trend and General

| Indicators | DPSE | Coefficient | t-value | std.error | df | p.value | Number of Countries with Increase | Sample Size |
|---|---|---|---|---|---|---|---|---|
| level 1 | | | | | | | | |
| Drivers Total | Drivers | -0.59 | -1.5 | 0.39 | 69.2 | 0.133 | 6 | 73 |
| Use Total | Use | -0.84 | -2.8 | 0.30 | 61.0 | **0.008** | 55 | 65 |
| Resistance Total | Resistance | -1.13 | -3.2 | 0.35 | 29.0 | **0.003** | 16 | 32 |
| DRI | DRI | -1.08 | -1.8 | 0.58 | 22.0 | 0.078 | 21 | 25 |
| level 2 | | | | | | | | |
| Infections | Drivers | 0.55 | 2.0 | 0.28 | 69.4 | 0.05 | 12 | 73 |
| Sanitation | Drivers | 0.40 | 1.6 | 0.24 | 69.1 | 0.107 | 27 | 73 |
| Vaccination | Drivers | -0.01 | 0.0 | 0.30 | 69.2 | 0.982 | 11 | 73 |
| Workforce | Drivers | -0.99 | -3.4 | 0.29 | 51.4 | **0.001** | 9 | 55 |
| TotalDDDPer1000Persons | Use | 0.23 | 0.8 | 0.30 | 61.3 | 0.435 | 50 | 65 |
| BroadPerTotalABXUse | Use | -0.67 | -2.7 | 0.25 | 61.0 | **0.009** | 47 | 65 |
| NewABXUse | Use | -0.79 | -2.1 | 0.37 | 59.0 | **0.036** | 55 | 63 |
| MRSA | Resistance | 0.29 | 0.6 | 0.48 | 28.5 | 0.551 | 11 | 32 |
| CR | Resistance | -0.52 | -0.9 | 0.55 | 24.8 | 0.356 | 20 | 28 |
| STR | Resistance | -0.76 | -1.6 | 0.48 | 21.9 | 0.129 | 13 | 25 |
| level 3 | | | | | | | | |
| HIV | Drivers/infections | 0.19 | 0.6 | 0.30 | 27.0 | 0.533 | 22 | 31 |
| TB | Drivers/infections | 0.67 | 2.4 | 0.28 | 69.2 | **0.02** | 11 | 73 |
| Drinking Water Source | Drivers/Sanitation | 1.00 | 2.8 | 0.35 | 68.8 | **0.006** | 65 | 72 |
| Water Source Access | Drivers/Sanitation | 0.96 | 2.7 | 0.36 | 68.9 | **0.009** | 65 | 72 |
| Overall Sanitation | Drivers/Sanitation | -0.32 | -0.6 | 0.56 | 62.8 | 0.573 | 63 | 66 |
| DTP3 | Drivers/Vaccination | 0.05 | 0.2 | 0.24 | 68.3 | 0.841 | 51 | 72 |
| HepB3 | Drivers/Vaccination | 0.20 | 0.8 | 0.26 | 56.0 | 0.436 | 48 | 60 |
| Hib3 | Drivers/Vaccination | -0.73 | -2.1 | 0.36 | 49.6 | **0.045** | 45 | 53 |
| Pol3 | Drivers/Vaccination | 0.41 | 1.8 | 0.23 | 68.0 | 0.076 | 49 | 72 |
| Measles | Drivers/Vaccination | 0.26 | 1.1 | 0.24 | 69.4 | 0.294 | 53 | 73 |
| RCV1 | Drivers/Vaccination | 0.42 | 1.5 | 0.28 | 58.7 | 0.135 | 43 | 62 |
| Nursing | Drivers/Workforce | 1.24 | 4.3 | 0.29 | 39.0 | **< .001** | 35 | 42 |
| Physicians | Drivers/Workforce | 0.61 | 2.2 | 0.28 | 51.0 | **0.036** | 44 | 55 |

lmer(General ~ Sign + Baseline + (1|income))

#### S16 Table. Linear Trend and Monitoring and Surveillance

| Indicators | DPSE | Coefficient | t-value | std.error | df | p.value | Number of Countries with Increase | Sample Size |
|---|---|---|---|---|---|---|---|---|
| level 1 | | | | | | | | |
| Drivers Total | Drivers | -0.06 | -3.1 | 0.02 | 39.1 | **0.003** | 6 | 73 |
| Use Total | Use | -0.19 | -4.1 | 0.05 | 62.0 | **< .001** | 55 | 65 |
| Resistance Total | Resistance | -0.15 | -1.1 | 0.13 | 18.7 | 0.281 | 16 | 32 |
| DRI | DRI | -0.14 | -1.1 | 0.13 | 21.1 | 0.278 | 21 | 25 |
| level 2 | | | | | | | | |
| Infections | Drivers | 0.00 | 0.1 | 0.01 | 39.2 | 0.896 | 12 | 73 |
| Sanitation | Drivers | 0.00 | -0.2 | 0.02 | 52.7 | 0.862 | 27 | 73 |
| Vaccination | Drivers | -0.07 | -1.6 | 0.04 | 70.0 | 0.112 | 11 | 73 |
| Workforce | Drivers | -0.15 | -2.9 | 0.05 | 52.0 | **0.006** | 9 | 55 |
| TotalDDDPer1000Persons | Use | -0.09 | -1.1 | 0.08 | 49.8 | 0.286 | 50 | 65 |
| BroadPerTotalABXUse | Use | -0.25 | -3.1 | 0.08 | 39.1 | **0.004** | 47 | 65 |
| NewABXUse | Use | -0.14 | -2.3 | 0.06 | 60.0 | **0.023** | 55 | 63 |
| MRSA | Resistance | -0.13 | -0.9 | 0.15 | 29.0 | 0.386 | 11 | 32 |
| CR | Resistance | -0.30 | -1.3 | 0.23 | 25.0 | 0.214 | 20 | 28 |
| STR | Resistance | -0.30 | -3.5 | 0.09 | 22.0 | **0.002** | 13 | 25 |
| level 3 | | | | | | | | |
| HIV | Drivers/infections | 0.01 | 0.8 | 0.01 | 28.0 | 0.419 | 22 | 31 |
| TB | Drivers/infections | 0.02 | 1.3 | 0.02 | 6.4 | 0.228 | 11 | 73 |
| Drinking Water Source | Drivers/Sanitation | 0.01 | 0.4 | 0.02 | 69.0 | 0.71 | 65 | 72 |
| Water Source Access | Drivers/Sanitation | 0.01 | 0.5 | 0.01 | 69.0 | 0.601 | 65 | 72 |
| Overall Sanitation | Drivers/Sanitation | 0.00 | 0.1 | 0.01 | 55.5 | 0.955 | 63 | 66 |
| DTP3 | Drivers/Vaccination | 0.07 | 1.6 | 0.05 | 69.0 | 0.124 | 51 | 72 |
| HepB3 | Drivers/Vaccination | 0.06 | 0.9 | 0.07 | 57.0 | 0.379 | 48 | 60 |
| Hib3 | Drivers/Vaccination | 0.11 | 2.1 | 0.05 | 50.0 | **0.042** | 45 | 53 |
| Pol3 | Drivers/Vaccination | 0.08 | 1.8 | 0.04 | 69.0 | 0.071 | 49 | 72 |
| Measles | Drivers/Vaccination | 0.07 | 1.8 | 0.04 | 70.0 | 0.08 | 53 | 73 |
| RCV1 | Drivers/Vaccination | 0.12 | 1.9 | 0.06 | 59.0 | 0.064 | 43 | 62 |
| Nursing | Drivers/Workforce | 0.13 | 1.8 | 0.07 | 39.0 | 0.074 | 35 | 42 |
| Physicians | Drivers/Workforce | 0.12 | 2.6 | 0.05 | 52.0 | **0.011** | 44 | 55 |

lmer(Linear Trend ~ Monitoring and Surveillance + Baseline + (1|income))

#### S17 Table. Categorical Trend and Monitoring and Surveillance

| Indicators | DPSE | Coefficient | t-value | std.error | df | p.value | Number of Countries with Increase | Sample Size |
|---|---|---|---|---|---|---|---|---|
| level 1 | | | | | | | | |
| Drivers Total | Drivers | -0.09 | -0.3 | 0.30 | 69.0 | 0.752 | 6 | 73 |
| Use Total | Use | -0.61 | -2.4 | 0.25 | 61.0 | **0.019** | 55 | 65 |
| Resistance Total | Resistance | -0.47 | -2.6 | 0.18 | 28.2 | **0.013** | 16 | 32 |
| DRI | DRI | -0.12 | -0.4 | 0.27 | 20.5 | 0.671 | 21 | 25 |
| level 2 | | | | | | | | |
| Infections | Drivers | -0.05 | -0.2 | 0.24 | 69.1 | 0.83 | 12 | 73 |
| Sanitation | Drivers | 0.45 | 2.3 | 0.19 | 69.0 | **0.022** | 27 | 73 |
| Vaccination | Drivers | -0.06 | -0.2 | 0.24 | 69.0 | 0.822 | 11 | 73 |
| Workforce | Drivers | -0.57 | -2.4 | 0.24 | 51.2 | **0.019** | 9 | 55 |
| TotalDDDPer1000Persons | Use | -0.28 | -1.2 | 0.25 | 61.1 | 0.253 | 50 | 65 |
| BroadPerTotalABXUse | Use | -0.58 | -2.8 | 0.20 | 61.0 | **0.006** | 47 | 65 |
| NewABXUse | Use | -0.43 | -1.4 | 0.31 | 59.0 | 0.164 | 55 | 63 |
| MRSA | Resistance | 0.03 | 0.1 | 0.23 | 28.4 | 0.902 | 11 | 32 |
| CR | Resistance | -0.53 | -2.0 | 0.27 | 24.6 | 0.058 | 20 | 28 |
| STR | Resistance | -0.54 | -2.4 | 0.23 | 22.0 | **0.025** | 13 | 25 |
| level 3 | | | | | | | | |
| HIV | Drivers/infections | -0.30 | -1.1 | 0.27 | 27.0 | 0.282 | 22 | 31 |
| TB | Drivers/infections | -0.02 | -0.1 | 0.25 | 69.0 | 0.939 | 11 | 73 |
| Drinking Water Source | Drivers/Sanitation | 0.32 | 1.1 | 0.30 | 68.5 | 0.285 | 65 | 72 |
| Water Source Access | Drivers/Sanitation | 0.35 | 1.2 | 0.30 | 68.5 | 0.243 | 65 | 72 |
| Overall Sanitation | Drivers/Sanitation | 0.02 | 0.1 | 0.44 | 62.3 | 0.957 | 63 | 66 |
| DTP3 | Drivers/Vaccination | -0.19 | -1.0 | 0.19 | 68.1 | 0.34 | 51 | 72 |
| HepB3 | Drivers/Vaccination | -0.14 | -0.6 | 0.24 | 56.0 | 0.557 | 48 | 60 |
| Hib3 | Drivers/Vaccination | -0.26 | -0.8 | 0.31 | 49.1 | 0.401 | 45 | 53 |
| Pol3 | Drivers/Vaccination | -0.05 | -0.3 | 0.19 | 68.0 | 0.785 | 49 | 72 |
| Measles | Drivers/Vaccination | 0.00 | 0.0 | 0.20 | 69.1 | 1 | 53 | 73 |
| RCV1 | Drivers/Vaccination | 0.13 | 0.6 | 0.22 | 58.2 | 0.564 | 43 | 62 |
| Nursing | Drivers/Workforce | 0.75 | 2.9 | 0.26 | 38.0 | **0.006** | 35 | 42 |
| Physicians | Drivers/Workforce | 0.48 | 2.0 | 0.24 | 51.0 | **0.048** | 44 | 55 |

lmer(Monitoring and Surveillance ~ Sign + Baseline + (1|income))

#### S18 Table. Linear Trend and Prevention

| Indicators | DPSE | Coefficient | t-value | std.error | df | p.value | Number of Countries with Increase | Sample Size |
|---|---|---|---|---|---|---|---|---|
| level 1 | | | | | | | | |
| Drivers Total | Drivers | -0.01 | -0.4 | 0.02 | 70.0 | 0.661 | 6 | 73 |
| Use Total | Use | -0.02 | -0.5 | 0.05 | 62.0 | 0.642 | 55 | 65 |
| Resistance Total | Resistance | -0.06 | -1.0 | 0.06 | 27.7 | 0.339 | 16 | 32 |
| DRI | DRI | -0.09 | -1.9 | 0.05 | 21.0 | 0.078 | 21 | 25 |
| level 2 | | | | | | | | |
| Infections | Drivers | 0.01 | 1.0 | 0.01 | 70.0 | 0.324 | 12 | 73 |
| Sanitation | Drivers | 0.02 | 1.7 | 0.01 | 70.0 | 0.096 | 27 | 73 |
| Vaccination | Drivers | -0.04 | -0.9 | 0.04 | 63.3 | 0.383 | 11 | 73 |
| Workforce | Drivers | -0.08 | -2.2 | 0.04 | 52.0 | **0.035** | 9 | 55 |
| TotalDDDPer1000Persons | Use | 0.03 | 0.5 | 0.07 | 62.0 | 0.65 | 50 | 65 |
| BroadPerTotalABXUse | Use | -0.09 | -1.3 | 0.07 | 62.0 | 0.211 | 47 | 65 |
| NewABXUse | Use | 0.01 | 0.2 | 0.06 | 58.4 | 0.822 | 55 | 63 |
| MRSA | Resistance | -0.10 | -1.4 | 0.07 | 29.0 | 0.173 | 11 | 32 |
| CR | Resistance | -0.07 | -0.6 | 0.13 | 21.4 | 0.583 | 20 | 28 |
| STR | Resistance | -0.10 | -1.7 | 0.06 | 22.0 | 0.106 | 13 | 25 |
| level 3 | | | | | | | | |
| HIV | Drivers/infections | 0.01 | 0.7 | 0.01 | 28.0 | 0.486 | 22 | 31 |
| TB | Drivers/infections | 0.01 | 0.5 | 0.02 | 70.0 | 0.64 | 11 | 73 |
| Drinking Water Source | Drivers/Sanitation | -0.02 | -1.4 | 0.01 | 69.0 | 0.153 | 65 | 72 |
| Water Source Access | Drivers/Sanitation | -0.02 | -1.4 | 0.01 | 69.0 | 0.158 | 65 | 72 |
| Overall Sanitation | Drivers/Sanitation | 0.01 | 0.7 | 0.01 | 62.3 | 0.506 | 63 | 66 |
| DTP3 | Drivers/Vaccination | 0.05 | 1.0 | 0.05 | 61.6 | 0.324 | 51 | 72 |
| HepB3 | Drivers/Vaccination | 0.08 | 1.3 | 0.06 | 57.0 | 0.212 | 48 | 60 |
| Hib3 | Drivers/Vaccination | -0.01 | -0.2 | 0.06 | 49.4 | 0.829 | 45 | 53 |
| Pol3 | Drivers/Vaccination | 0.08 | 1.8 | 0.04 | 50.4 | 0.084 | 49 | 72 |
| Measles | Drivers/Vaccination | 0.04 | 1.0 | 0.04 | 58.7 | 0.346 | 53 | 73 |
| RCV1 | Drivers/Vaccination | 0.06 | 0.9 | 0.06 | 56.7 | 0.385 | 43 | 62 |
| Nursing | Drivers/Workforce | 0.07 | 1.3 | 0.05 | 39.0 | 0.193 | 35 | 42 |
| Physicians | Drivers/Workforce | 0.08 | 2.0 | 0.04 | 52.0 | **0.046** | 44 | 55 |

lmer(Linear Trend ~ Prevention + Baseline + (1|income))

#### S19 Table. Categorical Trend and Prevention

| Indicators | DPSE | Coefficient | t-value | std.error | df | p.value | Number of Countries with Increase | Sample Size |
|---|---|---|---|---|---|---|---|---|
| level 1 | | | | | | | | |
| Drivers Total | Drivers | -0.11 | -0.3 | 0.42 | 69.3 | 0.796 | 6 | 73 |
| Use Total | Use | -0.54 | -1.6 | 0.33 | 61.0 | 0.107 | 55 | 65 |
| Resistance Total | Resistance | -0.67 | -1.7 | 0.39 | 24.1 | 0.096 | 16 | 32 |
| DRI | DRI | -0.80 | -1.3 | 0.62 | 22.0 | 0.209 | 21 | 25 |
| level 2 | | | | | | | | |
| Infections | Drivers | -0.03 | -0.1 | 0.30 | 69.7 | 0.926 | 12 | 73 |
| Sanitation | Drivers | 0.12 | 0.5 | 0.26 | 69.2 | 0.647 | 27 | 73 |
| Vaccination | Drivers | 0.08 | 0.3 | 0.31 | 69.3 | 0.802 | 11 | 73 |
| Workforce | Drivers | -0.35 | -1.0 | 0.36 | 51.3 | 0.335 | 9 | 55 |
| TotalDDDPer1000Persons | Use | -0.11 | -0.4 | 0.31 | 61.3 | 0.718 | 50 | 65 |
| BroadPerTotalABXUse | Use | -0.37 | -1.4 | 0.27 | 61.0 | 0.18 | 47 | 65 |
| NewABXUse | Use | -0.33 | -0.8 | 0.39 | 59.0 | 0.4 | 55 | 63 |
| MRSA | Resistance | 0.19 | 0.4 | 0.48 | 28.6 | 0.69 | 11 | 32 |
| CR | Resistance | -0.27 | -0.5 | 0.55 | 24.8 | 0.627 | 20 | 28 |
| STR | Resistance | -0.63 | -1.5 | 0.43 | 21.8 | 0.158 | 13 | 25 |
| level 3 | | | | | | | | |
| HIV | Drivers/infections | -0.42 | -1.3 | 0.32 | 27.0 | 0.202 | 22 | 31 |
| TB | Drivers/infections | -0.07 | -0.2 | 0.31 | 69.4 | 0.835 | 11 | 73 |
| Drinking Water Source | Drivers/Sanitation | 0.18 | 0.5 | 0.39 | 68.3 | 0.644 | 65 | 72 |
| Water Source Access | Drivers/Sanitation | 0.46 | 1.2 | 0.37 | 69.0 | 0.223 | 65 | 72 |
| Overall Sanitation | Drivers/Sanitation | -0.20 | -0.4 | 0.51 | 59.0 | 0.703 | 63 | 66 |
| DTP3 | Drivers/Vaccination | 0.02 | 0.1 | 0.25 | 68.4 | 0.952 | 51 | 72 |
| HepB3 | Drivers/Vaccination | 0.38 | 1.2 | 0.31 | 56.0 | 0.233 | 48 | 60 |
| Hib3 | Drivers/Vaccination | -0.22 | -0.6 | 0.36 | 49.7 | 0.544 | 45 | 53 |
| Pol3 | Drivers/Vaccination | 0.37 | 1.5 | 0.24 | 68.1 | 0.135 | 49 | 72 |
| Measles | Drivers/Vaccination | 0.02 | 0.1 | 0.26 | 69.5 | 0.931 | 53 | 73 |
| RCV1 | Drivers/Vaccination | -0.01 | 0.0 | 0.30 | 59.0 | 0.965 | 43 | 62 |
| Nursing | Drivers/Workforce | 0.61 | 1.7 | 0.36 | 38.0 | 0.101 | 35 | 42 |
| Physicians | Drivers/Workforce | 0.74 | 2.3 | 0.32 | 51.0 | **0.023** | 44 | 55 |

lmer(Prevention ~ Sign + Baseline + (1|income))

#### S20 Table. Linear Trend and Regulation

| Indicators | DPSE | Coefficient | t-value | std.error | df | p.value | Number of Countries with Increase | Sample Size |
|---|---|---|---|---|---|---|---|---|
| level 1 | | | | | | | | |
| Drivers Total | Drivers | -0.04 | -2.8 | 0.01 | 26.0 | **0.01** | 6 | 73 |
| Use Total | Use | -0.08 | -1.5 | 0.05 | 60.6 | 0.129 | 55 | 65 |
| Resistance Total | Resistance | -0.01 | -0.1 | 0.10 | 28.2 | 0.946 | 16 | 32 |
| DRI | DRI | -0.10 | -1.3 | 0.08 | 21.1 | 0.207 | 21 | 25 |
| level 2 | | | | | | | | |
| Infections | Drivers | -0.01 | -0.8 | 0.01 | 68.5 | 0.405 | 12 | 73 |
| Sanitation | Drivers | -0.02 | -1.9 | 0.01 | 69.8 | 0.058 | 27 | 73 |
| Vaccination | Drivers | -0.06 | -1.6 | 0.04 | 70.0 | 0.121 | 11 | 73 |
| Workforce | Drivers | -0.12 | -3.1 | 0.04 | 52.0 | **0.003** | 9 | 55 |
| TotalDDDPer1000Persons | Use | 0.02 | 0.3 | 0.07 | 62.0 | 0.758 | 50 | 65 |
| BroadPerTotalABXUse | Use | -0.17 | -2.4 | 0.07 | 20.4 | **0.026** | 47 | 65 |
| NewABXUse | Use | -0.07 | -1.1 | 0.06 | 60.0 | 0.258 | 55 | 63 |
| MRSA | Resistance | -0.13 | -1.2 | 0.11 | 29.0 | 0.255 | 11 | 32 |
| CR | Resistance | 0.09 | 0.4 | 0.21 | 24.5 | 0.677 | 20 | 28 |
| STR | Resistance | 0.04 | 0.5 | 0.08 | 21.4 | 0.636 | 13 | 25 |
| level 3 | | | | | | | | |
| HIV | Drivers/infections | 0.00 | 0.6 | 0.01 | 28.0 | 0.555 | 22 | 31 |
| TB | Drivers/infections | 0.00 | 0.0 | 0.02 | 61.8 | 0.97 | 11 | 73 |
| Drinking Water Source | Drivers/Sanitation | 0.02 | 1.6 | 0.01 | 49.5 | 0.106 | 65 | 72 |
| Water Source Access | Drivers/Sanitation | 0.02 | 1.8 | 0.01 | 44.4 | 0.086 | 65 | 72 |
| Overall Sanitation | Drivers/Sanitation | 0.00 | 0.1 | 0.01 | 61.5 | 0.933 | 63 | 66 |
| DTP3 | Drivers/Vaccination | 0.08 | 1.8 | 0.04 | 69.0 | 0.08 | 51 | 72 |
| HepB3 | Drivers/Vaccination | 0.04 | 0.6 | 0.06 | 57.0 | 0.529 | 48 | 60 |
| Hib3 | Drivers/Vaccination | 0.01 | 0.1 | 0.06 | 44.0 | 0.902 | 45 | 53 |
| Pol3 | Drivers/Vaccination | 0.08 | 2.0 | 0.04 | 69.0 | **0.046** | 49 | 72 |
| Measles | Drivers/Vaccination | 0.10 | 2.7 | 0.03 | 70.0 | **0.008** | 53 | 73 |
| RCV1 | Drivers/Vaccination | 0.11 | 1.9 | 0.06 | 59.0 | 0.057 | 43 | 62 |
| Nursing | Drivers/Workforce | 0.13 | 2.5 | 0.05 | 39.0 | **0.015** | 35 | 42 |
| Physicians | Drivers/Workforce | 0.11 | 2.7 | 0.04 | 52.0 | **0.009** | 44 | 55 |

lmer(Linear Trend ~ Regulation + Baseline + (1|income))

#### S21 Table. Categorical Trend and Regulation

| Indicators | DPSE | Coefficient | t-value | std.error | df | p.value | Number of Countries with Increase | Sample Size |
|---|---|---|---|---|---|---|---|---|
| level 1 | | | | | | | | |
| Drivers Total | Drivers | -0.61 | -1.6 | 0.39 | 69.1 | 0.122 | 6 | 73 |
| Use Total | Use | -0.60 | -2.0 | 0.30 | 61.0 | 0.051 | 55 | 65 |
| Resistance Total | Resistance | -0.43 | -1.6 | 0.26 | 29.0 | 0.115 | 16 | 32 |
| DRI | DRI | -1.04 | -2.8 | 0.38 | 21.9 | **0.012** | 21 | 25 |
| level 2 | | | | | | | | |
| Infections | Drivers | 0.27 | 0.9 | 0.29 | 69.2 | 0.363 | 12 | 73 |
| Sanitation | Drivers | 0.03 | 0.1 | 0.25 | 69.1 | 0.91 | 27 | 73 |
| Vaccination | Drivers | -0.19 | -0.6 | 0.30 | 69.1 | 0.532 | 11 | 73 |
| Workforce | Drivers | -0.37 | -1.1 | 0.34 | 51.4 | 0.285 | 9 | 55 |
| TotalDDDPer1000Persons | Use | -0.08 | -0.3 | 0.29 | 61.3 | 0.773 | 50 | 65 |
| BroadPerTotalABXUse | Use | -0.62 | -2.6 | 0.24 | 61.0 | **0.012** | 47 | 65 |
| NewABXUse | Use | -0.82 | -2.3 | 0.35 | 59.0 | **0.023** | 55 | 63 |
| MRSA | Resistance | -0.37 | -1.2 | 0.32 | 29.0 | 0.256 | 11 | 32 |
| CR | Resistance | -0.17 | -0.5 | 0.35 | 25.0 | 0.643 | 20 | 28 |
| STR | Resistance | -0.50 | -1.5 | 0.32 | 22.0 | 0.139 | 13 | 25 |
| level 3 | | | | | | | | |
| HIV | Drivers/infections | -0.41 | -1.2 | 0.35 | 27.0 | 0.246 | 22 | 31 |
| TB | Drivers/infections | 0.22 | 0.7 | 0.30 | 69.1 | 0.473 | 11 | 73 |
| Drinking Water Source | Drivers/Sanitation | 0.51 | 1.4 | 0.37 | 68.8 | 0.179 | 65 | 72 |
| Water Source Access | Drivers/Sanitation | 0.43 | 1.1 | 0.37 | 68.8 | 0.255 | 65 | 72 |
| Overall Sanitation | Drivers/Sanitation | -0.64 | -1.2 | 0.55 | 62.5 | 0.253 | 63 | 66 |
| DTP3 | Drivers/Vaccination | 0.09 | 0.4 | 0.24 | 68.1 | 0.712 | 51 | 72 |
| HepB3 | Drivers/Vaccination | 0.06 | 0.2 | 0.31 | 56.0 | 0.847 | 48 | 60 |
| Hib3 | Drivers/Vaccination | 0.03 | 0.1 | 0.34 | 49.2 | 0.94 | 45 | 53 |
| Pol3 | Drivers/Vaccination | 0.36 | 1.5 | 0.24 | 68.0 | 0.132 | 49 | 72 |
| Measles | Drivers/Vaccination | 0.29 | 1.2 | 0.25 | 69.2 | 0.236 | 53 | 73 |
| RCV1 | Drivers/Vaccination | 0.27 | 1.0 | 0.27 | 58.4 | 0.336 | 43 | 62 |
| Nursing | Drivers/Workforce | 0.76 | 2.1 | 0.36 | 38.0 | **0.041** | 35 | 42 |
| Physicians | Drivers/Workforce | 0.47 | 1.5 | 0.31 | 51.0 | 0.135 | 44 | 55 |

lmer(Regulation ~ Sign + Baseline + (1|income))

**S22 Table. De-escalation Merged Model Comparison Results.**
Merged model comparison for de-escalation to investigate the importance of action for declining proportion, including all DPSE categories. df: residual degrees of freedom. AICc: Akaike information criterion corrected for small sample sizes. Delta AICc: Difference between the AICc of the best model and the given model. Weight is the AICc weight.

| Model Formula | df | logLik | AICc | Delta AICc | weight |
|---|---|---|---|---|---|
| Declining proportion ~ Action * DPSE + Baseline Mean | 186 | -179.69 | 378.36 | 0 | 1 |
| Declining proportion ~ Action + DPSE + Baseline Mean | 189 | -192.48 | 397.40 | 19.04 | 0 |
| Declining proportion ~ Baseline Mean | 193 | -255.20 | 514.47 | 136.11 | 0 |
| Declining proportion ~ Action + Baseline Mean | 192 | -255.14 | 516.40 | 138.04 | 0 |

**S23 Table. De-escalation Merged Model Results.**
Merged model results for de-escalation to investigate the importance of action. Model results derived from the merged model with the main formula as in S4 Table and reported for all the categories.

| Indicators | Odds Ratio | CI | P value |
|---|---|---|---|
| Action (DRIVERS) | 0.62 | 0.41 – 0.92 | **0.018** |
| Action (USE-DRIVERS) | 4.23 | 2.26 – 8.11 | **<0.001** |
| Action (RESISTANCE-DRIVERS) | 2.25 | 1.11 – 4.65 | **0.026** |
| Action (DRI-DRIVERS) | 13.53 | 1.87 – 317.75 | **0.035** |

**S24 Table. Model selection table including variables for best selected models and null models.**

Model selection summary tables included first 5 best selected models and the null model to determine the most important variables affecting Antibiotic Resistance (ABR) linear trend in 16 years. Two model types are included in the table. First model type used linear trend as a response variable and determining the change of ABR indicators between two intervals (2008-2016 vs 2000-2008). Second model type used categorical trend as a response variable, and it refers to the positive or negative change. Note that linear trend models run with different datasets including data from i. all countries, ii. High Income Countries, iii. Low- and Middle-Income Countries according to the income column. Overall, all models run with a specific data subset and named accordingly (see Fig 4 capture for model names). DPSEA refers to the indicators from the framework (see methods), Baseline refers to the Mean of DPSE Indicator in Baseline (2000−2008), income refers to the high income of low and middle income. For country names matching with ISO3 codes see S1 Table, for Tier 2 indicators and other variable abbreviations see S2 Table. df =degrees of freedom. AICc: Akaike information criterion corrected for small sample sizes. Delta AICc: Difference between the AICc of the best model and the given model. Action variables are indicated in *italics*.

| Model Name | Variables | df | logLik | AICc | delta | weight | income |
|---|---|---|---|---|---|---|---|
| **DPSEA** | Linear Trend ~ *General* + Baseline + DPSE * Workforce | 12 | -39.912 | 106.022 | 0.000 | 0.314 | ALL |
| **DPSEA** | Linear Trend ~ *Action* + Baseline + DPSE * Workforce | 12 | -40.212 | 106.621 | 0.599 | 0.233 | ALL |
| **DPSEA** | Linear Trend ~ Baseline + DPSE * Workforce | 11 | -41.480 | 106.806 | 0.785 | 0.212 | ALL |
| **DPSEA** | Linear Trend ~ *Monitoring and Surveillance* + Baseline + DPSE * Workforce | 12 | -40.800 | 107.798 | 1.776 | 0.129 | ALL |
| **DPSEA** | Linear Trend ~ Vaccination + Baseline + DPSE * Workforce | 12 | -42.785 | 111.767 | 5.745 | 0.018 | ALL |
| **DPSEA** | Linear Trend ~ 1 | 3 | -71.821 | 149.801 | 43.779 | 0.000 | ALL |
| **DPSEA.noDr** | Linear Trend ~ DPSE + *General* | 7 | -52.507 | 119.614 | 0.000 | 0.420 | ALL |
| **DPSEA.noDr** | Linear Trend ~ DPSE + *Action* | 7 | -53.329 | 121.257 | 1.643 | 0.185 | ALL |
| **DPSEA.noDr** | Linear Trend ~ DPSE | 6 | -54.974 | 122.394 | 2.781 | 0.105 | ALL |
| **DPSEA.noDr** | Linear Trend ~ DPSE + *Monitoring and Surveillance* | 7 | -54.244 | 123.087 | 3.473 | 0.074 | ALL |
| **DPSEA.noDr** | Linear Trend ~ DPSE + *General* + Baseline | 8 | -54.337 | 125.448 | 5.834 | 0.023 | ALL |
| **DPSEA.noDr** | Linear Trend ~ 1 | 3 | -78.520 | 163.165 | 43.551 | 0.000 | ALL |
| **aP.noDr** | Linear Trend ~ DPSE + *General* | 7 | -54.390 | 123.410 | 0.000 | 0.394 | ALL |
| **aP.noDr** | Linear Trend ~ DPSE + *Action* | 7 | -55.026 | 124.682 | 1.272 | 0.209 | ALL |
| **aP.noDr** | Linear Trend ~ DPSE | 6 | -56.727 | 125.924 | 2.514 | 0.112 | ALL |
| **aP.noDr** | Linear Trend ~ DPSE + *Monitoring and Surveillance* | 7 | -55.771 | 126.172 | 2.762 | 0.099 | ALL |
| **aP.noDr** | Linear Trend ~ DPSE + *General* + *Action* | 8 | -56.214 | 129.242 | 5.832 | 0.021 | ALL |
| **aP.noDr** | Linear Trend ~ 1 | 3 | -78.269 | 162.670 | 39.260 | 0.000 | ALL |
| **aS.noDr** | Linear Trend ~ DPSE + *General* | 7 | -44.454 | 103.720 | 0.000 | 0.345 | ALL |
| **aS.noDr** | Linear Trend ~ DPSE | 6 | -45.663 | 103.929 | 0.209 | 0.311 | ALL |
| **aS.noDr** | Linear Trend ~ DPSE + *Action* | 7 | -45.650 | 106.113 | 2.392 | 0.104 | ALL |
| **aS.noDr** | Linear Trend ~ DPSE + *Monitoring and Surveillance* | 7 | -46.370 | 107.551 | 3.831 | 0.051 | ALL |

| **Model Name** | **Variables** | **df** | **logLik** | **AICc** | **delta** | **weight** | **income** |
|---|---|---|---|---|---|---|---|
| **aS.noDr** | Linear Trend ~ *Awareness and Education* + DPSE + *General* | 8 | -45.499 | 108.050 | 4.330 | 0.040 | ALL |
| **aS.noDr** | Linear Trend ~ 1 | 3 | -56.134 | 118.437 | 14.717 | 0.000 | ALL |
| **aE.noDr** | Linear Trend ~ DPSE | 6 | -42.937 | 98.546 | 0.000 | 0.473 | ALL |
| **aE.noDr** | Linear Trend ~ DPSE + *General* | 7 | -42.679 | 100.260 | 1.715 | 0.201 | ALL |
| **aE.noDr** | Linear Trend ~ DPSE + *Action* | 7 | -43.425 | 101.754 | 3.208 | 0.095 | ALL |
| **aE.noDr** | Linear Trend ~ DPSE + *Monitoring and Surveillance* | 7 | -43.622 | 102.147 | 3.601 | 0.078 | ALL |
| **aE.noDr** | Linear Trend ~ *Awareness and Education* + DPSE | 7 | -44.668 | 104.240 | 5.694 | 0.027 | ALL |
| **aE.noDr** | Linear Trend ~ 1 | 3 | -50.749 | 107.686 | 9.140 | 0.005 | ALL |
| **aP** | Linear Trend ~ DPSE * Workforce | 10 | -46.859 | 115.300 | 0.000 | 0.223 | ALL |
| **aP** | Linear Trend ~ DPSE | 6 | -51.456 | 115.500 | 0.200 | 0.201 | ALL |
| **aP** | Linear Trend ~ DPSE + *General* | 7 | -50.387 | 115.562 | 0.262 | 0.195 | ALL |
| **aP** | Linear Trend ~ DPSE + *Action* | 7 | -51.183 | 117.154 | 1.854 | 0.088 | ALL |
| **aP** | Linear Trend ~ DPSE + *Monitoring and Surveillance* | 7 | -52.167 | 119.122 | 3.822 | 0.033 | ALL |
| **aP** | Linear Trend ~ 1 | 3 | -71.261 | 148.686 | 33.386 | 0.000 | ALL |
| **aS** | Linear Trend ~ DPSE | 6 | -43.015 | 98.853 | 0.000 | 0.296 | ALL |
| **aS** | Linear Trend ~ DPSE * Sanitation | 10 | -38.625 | 99.494 | 0.641 | 0.215 | ALL |
| **aS** | Linear Trend ~ DPSE + *General* | 7 | -42.719 | 100.547 | 1.694 | 0.127 | ALL |
| **aS** | Linear Trend ~ DPSE + *Action* | 7 | -43.710 | 102.529 | 3.676 | 0.047 | ALL |
| **aS** | Linear Trend ~ DPSE * Workforce | 10 | -40.240 | 102.724 | 3.871 | 0.043 | ALL |
| **aS** | Linear Trend ~ 1 | 3 | -50.850 | 107.929 | 9.075 | 0.003 | ALL |
| **aE** | Linear Trend ~ DPSE | 6 | -39.877 | 92.667 | 0.000 | 0.313 | ALL |
| **aE** | Linear Trend ~ DPSE * Workforce | 10 | -35.385 | 93.270 | 0.603 | 0.231 | ALL |
| **aE** | Linear Trend ~ DPSE * Sanitation | 10 | -36.625 | 95.750 | 3.083 | 0.067 | ALL |
| **aE** | Linear Trend ~ DPSE + *General* | 7 | -40.344 | 95.919 | 3.252 | 0.061 | ALL |
| **aE** | Linear Trend ~ DPSE + *Action* | 7 | -40.895 | 97.021 | 4.354 | 0.035 | ALL |
| **aE** | Linear Trend ~ 1 | 3 | -45.471 | 97.195 | 4.528 | 0.032 | ALL |
| **Dr** | Linear Trend ~ Baseline | 5 | -9.837 | 29.899 | 0.000 | 0.790 | ALL |
| **Dr** | Linear Trend ~ *Action* + Baseline | 6 | -11.505 | 35.324 | 5.425 | 0.052 | ALL |
| **Dr** | Linear Trend ~ *General* + Baseline | 6 | -11.620 | 35.555 | 5.656 | 0.047 | ALL |
| **Dr** | Linear Trend ~ *Awareness and Education* + Baseline | 6 | -12.053 | 36.422 | 6.523 | 0.030 | ALL |
| **Dr** | Linear Trend ~ income + Baseline | 6 | -12.148 | 36.611 | 6.713 | 0.028 | ALL |
| **Dr** | Linear Trend ~ 1 | 4 | -16.607 | 41.362 | 11.463 | 0.003 | ALL |
| **P** | Linear Trend ~ *Monitoring and Surveillance* | 5 | -125.120 | 260.614 | 0.000 | 0.134 | ALL |
| **P** | Linear Trend ~ Workforce | 5 | -125.162 | 260.699 | 0.085 | 0.128 | ALL |
| **P** | Linear Trend ~ *Action* | 5 | -125.799 | 261.973 | 1.359 | 0.068 | ALL |
| **P** | Linear Trend ~ 1 | 4 | -126.877 | 262.002 | 1.388 | 0.067 | ALL |
| **P** | Linear Trend ~ income | 5 | -125.877 | 262.130 | 1.515 | 0.063 | ALL |
| **P** | Linear Trend ~ 1 | 4 | -126.877 | 262.002 | 1.388 | 0.067 | ALL |
| **S** | Linear Trend ~ income + Baseline | 6 | -53.321 | 120.228 | 0.000 | 0.083 | ALL |
| **S** | Linear Trend ~ *Monitoring and Surveillance* + Baseline | 6 | -53.491 | 120.566 | 0.338 | 0.070 | ALL |
| **S** | Linear Trend ~ Baseline | 5 | -54.894 | 120.900 | 0.672 | 0.059 | ALL |

| Model Name | Variables | df | logLik | AICc | delta | weight | income |
|---|---|---|---|---|---|---|---|
| **S** | Linear Trend ~ Infection + Mean Temperature + Baseline | 7 | -52.563 | 121.279 | 1.052 | 0.049 | ALL |
| **S** | Linear Trend ~ Mean Temperature + Baseline | 6 | -53.871 | 121.326 | 1.099 | 0.048 | ALL |
| **S** | Linear Trend ~ 1 | 4 | -57.515 | 123.757 | 3.530 | 0.014 | ALL |
| **E** | Linear Trend ~ income + Mean Temperature + *Action* | 5 | 7.656 | 0.142 | 0.000 | 0.368 | ALL |
| **E** | Linear Trend ~ *General* + income + Mean Temperature | 5 | 6.901 | 1.652 | 1.510 | 0.173 | ALL |
| **E** | Linear Trend ~ *Awareness and Education* + income + Mean Temperature | 5 | 6.054 | 3.346 | 3.205 | 0.074 | ALL |
| **E** | Linear Trend ~ income + Workforce + Baseline | 5 | 5.353 | 4.749 | 4.607 | 0.037 | ALL |
| **E** | Linear Trend ~ income + Mean Temperature + *Monitoring and Surveillance* | 5 | 5.285 | 4.884 | 4.742 | 0.034 | ALL |
| **E** | Linear Trend ~ 1 | 2 | -7.159 | 19.174 | 19.032 | 0.000 | ALL |
| **DPS** | Linear Trend ~ DPSE | 5 | -42.065 | 94.584 | 0.000 | 0.243 | ALL |
| **DPS** | Linear Trend ~ DPSE + *General* | 6 | -41.273 | 95.188 | 0.604 | 0.179 | ALL |
| **DPS** | Linear Trend ~ DPSE + *Action* | 6 | -42.150 | 96.942 | 2.358 | 0.075 | ALL |
| **DPS** | Linear Trend ~ DPSE * Workforce | 8 | -40.105 | 97.327 | 2.743 | 0.062 | ALL |
| **DPS** | Linear Trend ~ DPSE + *Action* + Baseline | 7 | -41.402 | 97.665 | 3.081 | 0.052 | ALL |
| **DPS** | Linear Trend ~ 1 | 3 | -64.124 | 134.428 | 39.844 | 0.000 | ALL |
| **PSE** | Linear Trend ~ *Monitoring and Surveillance* | 4 | -47.422 | 103.283 | 0.000 | 0.204 | ALL |
| **PSE** | Linear Trend ~ *Action* | 4 | -48.224 | 104.888 | 1.605 | 0.092 | ALL |
| **PSE** | Linear Trend ~ income | 4 | -48.881 | 106.202 | 2.919 | 0.047 | ALL |
| **PSE** | Linear Trend ~ 1 | 3 | -50.434 | 107.129 | 3.846 | 0.030 | ALL |
| **PSE** | Linear Trend ~ *General* | 4 | -49.358 | 107.155 | 3.872 | 0.029 | ALL |
| **PSE** | Linear Trend ~ 1 | 3 | -50.434 | 107.129 | 3.846 | 0.030 | ALL |
| **DP** | Linear Trend ~ DPSE * Workforce | 6 | -21.956 | 56.712 | 0.000 | 0.258 | ALL |
| **DP** | Linear Trend ~ DPSE * *Monitoring and Surveillance* | 6 | -22.670 | 58.139 | 1.427 | 0.126 | ALL |
| **DP** | Linear Trend ~ DPSE | 4 | -24.923 | 58.219 | 1.507 | 0.121 | ALL |
| **DP** | Linear Trend ~ DPSE + *Monitoring and Surveillance* + Baseline | 6 | -22.782 | 58.363 | 1.651 | 0.113 | ALL |
| **DP** | Linear Trend ~ DPSE + *Action* + Baseline | 6 | -23.561 | 59.922 | 3.210 | 0.052 | ALL |
| **DP** | Linear Trend ~ 1 | 3 | -50.092 | 106.405 | 49.693 | 0.000 | ALL |
| **PS** | Linear Trend ~ Baseline * income | 6 | -36.503 | 86.172 | 0.000 | 0.425 | ALL |
| **PS** | Linear Trend ~ *Monitoring and Surveillance* | 4 | -40.499 | 89.538 | 3.366 | 0.079 | ALL |
| **PS** | Linear Trend ~ income | 4 | -41.170 | 90.881 | 4.709 | 0.040 | ALL |
| **PS** | Linear Trend ~ Baseline + income | 5 | -40.237 | 91.297 | 5.124 | 0.033 | ALL |
| **PS** | Linear Trend ~ *Monitoring and Surveillance* + Baseline | 5 | -40.307 | 91.435 | 5.263 | 0.031 | ALL |
| **PS** | Linear Trend ~ 1 | 3 | -44.120 | 94.560 | 8.387 | 0.006 | ALL |
| **SE** | Linear Trend ~ Infection + Mean Temperature + Workforce | 6 | -17.852 | 50.249 | 0.000 | 0.083 | ALL |
| **SE** | Linear Trend ~ Sanitation * income | 6 | -18.165 | 50.875 | 0.625 | 0.061 | ALL |
| **SE** | Linear Trend ~ 1 | 3 | -22.129 | 50.925 | 0.676 | 0.059 | ALL |

| **Model Name** | **Variables** | **df** | **logLik** | **AICc** | **delta** | **weight** | **income** |
|---|---|---|---|---|---|---|---|
| **SE** | Linear Trend ~ Baseline * income | 6 | -18.258 | 51.061 | 0.812 | 0.055 | ALL |
| **SE** | Linear Trend ~ Infection + Workforce | 5 | -19.746 | 51.257 | 1.008 | 0.050 | ALL |
| **SE** | Linear Trend ~ 1 | 3 | -22.129 | 50.925 | 0.676 | 0.059 | ALL |
| **DPSEA** | Linear Trend ~ DPSE + *Monitoring and Surveillance* | 7 | -20.925 | 57.138 | 0.000 | 0.083 | HIC |
| **DPSEA** | Linear Trend ~ *General* + Mean Temperature + DPSE * Workforce | 12 | -14.971 | 57.746 | 0.608 | 0.061 | HIC |
| **DPSEA** | Linear Trend ~ *Monitoring and Surveillance* + DPSE * Workforce | 11 | -16.400 | 57.980 | 0.842 | 0.055 | HIC |
| **DPSEA** | Linear Trend ~ DPSE | 6 | -22.563 | 58.081 | 0.942 | 0.052 | HIC |
| **DPSEA** | Linear Trend ~ Mean Temperature + *Action* + DPSE * Workforce | 12 | -15.399 | 58.604 | 1.466 | 0.040 | HIC |
| **DPSEA** | Linear Trend ~ 1 | 3 | -28.843 | 63.951 | 6.813 | 0.003 | HIC |
| **DPSEA.noDr** | Linear Trend ~ DPSE + *Monitoring and Surveillance* | 7 | -18.484 | 51.958 | 0.000 | 0.479 | HIC |
| **DPSEA.noDr** | Linear Trend ~ DPSE + *General* | 7 | -20.233 | 55.458 | 3.499 | 0.083 | HIC |
| **DPSEA.noDr** | Linear Trend ~ DPSE + *Action* | 7 | -20.444 | 55.879 | 3.921 | 0.067 | HIC |
| **DPSEA.noDr** | Linear Trend ~ DPSE | 6 | -21.759 | 56.255 | 4.297 | 0.056 | HIC |
| **DPSEA.noDr** | Linear Trend ~ DPSE * *Monitoring and Surveillance* | 10 | -17.210 | 56.419 | 4.461 | 0.051 | HIC |
| **DPSEA.noDr** | Linear Trend ~ 1 | 3 | -31.466 | 69.138 | 17.179 | 0.000 | HIC |
| **aP.noDr** | Linear Trend ~ DPSE + *Monitoring and Surveillance* | 7 | -20.371 | 55.788 | 0.000 | 0.478 | HIC |
| **aP.noDr** | Linear Trend ~ DPSE + *General* | 7 | -22.203 | 59.453 | 3.665 | 0.077 | HIC |
| **aP.noDr** | Linear Trend ~ DPSE | 6 | -23.458 | 59.695 | 3.906 | 0.068 | HIC |
| **aP.noDr** | Linear Trend ~ DPSE + *Action* | 7 | -22.366 | 59.779 | 3.991 | 0.065 | HIC |
| **aP.noDr** | Linear Trend ~ DPSE + *General* + Mean Temperature | 8 | -21.403 | 60.164 | 4.376 | 0.054 | HIC |
| **aP.noDr** | Linear Trend ~ 1 | 3 | -32.479 | 71.174 | 15.386 | 0.000 | HIC |
| **aS.noDr** | Linear Trend ~ DPSE | 6 | -16.013 | 44.850 | 0.000 | 0.295 | HIC |
| **aS.noDr** | Linear Trend ~ DPSE + *General* | 7 | -15.157 | 45.423 | 0.573 | 0.222 | HIC |
| **aS.noDr** | Linear Trend ~ DPSE + *Monitoring and Surveillance* | 7 | -15.597 | 46.304 | 1.454 | 0.143 | HIC |
| **aS.noDr** | Linear Trend ~ DPSE + *Action* | 7 | -15.988 | 47.085 | 2.235 | 0.097 | HIC |
| **aS.noDr** | Linear Trend ~ DPSE + Baseline | 7 | -16.755 | 48.619 | 3.769 | 0.045 | HIC |
| **aS.noDr** | Linear Trend ~ 1 | 3 | -23.229 | 52.686 | 7.836 | 0.006 | HIC |
| **aE.noDr** | Linear Trend ~ DPSE | 6 | -17.860 | 48.596 | 0.000 | 0.322 | HIC |
| **aE.noDr** | Linear Trend ~ DPSE + *General* | 7 | -17.271 | 49.722 | 1.126 | 0.184 | HIC |
| **aE.noDr** | Linear Trend ~ DPSE + *Monitoring and Surveillance* | 7 | -17.493 | 50.165 | 1.569 | 0.147 | HIC |
| **aE.noDr** | Linear Trend ~ DPSE + *Action* | 7 | -18.018 | 51.216 | 2.620 | 0.087 | HIC |
| **aE.noDr** | Linear Trend ~ DPSE + Baseline | 7 | -18.792 | 52.762 | 4.166 | 0.040 | HIC |
| **aE.noDr** | Linear Trend ~ 1 | 3 | -24.501 | 55.245 | 6.649 | 0.012 | HIC |
| **aP** | Linear Trend ~ DPSE * Workforce | 10 | -18.012 | 58.774 | 0.000 | 0.190 | HIC |
| **aP** | Linear Trend ~ DPSE + *Monitoring and Surveillance* | 7 | -21.976 | 59.300 | 0.526 | 0.146 | HIC |
| **aP** | Linear Trend ~ DPSE | 6 | -23.598 | 60.196 | 1.422 | 0.093 | HIC |
| **aP** | Linear Trend ~ DPSE + *Monitoring and Surveillance* + Workforce | 8 | -21.730 | 61.217 | 2.443 | 0.056 | HIC |
| **aP** | Linear Trend ~ DPSE + Infection + *Monitoring and Surveillance* | 8 | -21.777 | 61.311 | 2.537 | 0.053 | HIC |
| **aP** | Linear Trend ~ 1 | 3 | -29.328 | 64.933 | 6.159 | 0.009 | HIC |

| Model Name | Variables | df | logLik | AICc | delta | weight | income |
|---|---|---|---|---|---|---|---|
| **aS** | Linear Trend ~ DPSE | 6 | -17.385 | 47.876 | 0.000 | 0.120 | HIC |
| **aS** | Linear Trend ~ 1 | 3 | -21.007 | 48.319 | 0.443 | 0.096 | HIC |
| **aS** | Linear Trend ~ DPSE * Infection | 10 | -13.264 | 49.583 | 1.707 | 0.051 | HIC |
| **aS** | Linear Trend ~ DPSE + Infection | 7 | -17.149 | 49.790 | 1.915 | 0.046 | HIC |
| **aS** | Linear Trend ~ Infection | 4 | -20.769 | 50.051 | 2.175 | 0.040 | HIC |
| **aS** | Linear Trend ~ 1 | 3 | -21.007 | 48.319 | 0.443 | 0.096 | HIC |
| **aE** | Linear Trend ~ 1 | 3 | -21.685 | 49.690 | 0.000 | 0.100 | HIC |
| **aE** | Linear Trend ~ DPSE | 6 | -18.520 | 50.207 | 0.517 | 0.077 | HIC |
| **aE** | Linear Trend ~ Workforce | 4 | -21.122 | 50.784 | 1.094 | 0.058 | HIC |
| **aE** | Linear Trend ~ Infection | 4 | -21.388 | 51.317 | 1.627 | 0.044 | HIC |
| **aE** | Linear Trend ~ *Monitoring and Surveillance* + Workforce | 5 | -20.292 | 51.406 | 1.716 | 0.043 | HIC |
| **aE** | Linear Trend ~ 1 | 3 | -21.685 | 49.690 | 0.000 | 0.100 | HIC |
| **Dr** | Linear Trend ~ Baseline | 5 | 26.282 | -42.114 | 0.000 | 0.587 | HIC |
| **Dr** | Linear Trend ~ *Action* + Baseline | 6 | 26.156 | -39.676 | 2.438 | 0.174 | HIC |
| **Dr** | Linear Trend ~ *Monitoring and Surveillance* + Baseline | 6 | 25.471 | -38.305 | 3.809 | 0.087 | HIC |
| **Dr** | Linear Trend ~ 1 | 4 | 22.677 | -37.056 | 5.058 | 0.047 | HIC |
| **Dr** | Linear Trend ~ *Awareness and Education* + Baseline | 6 | 24.562 | -36.489 | 5.625 | 0.035 | HIC |
| **Dr** | Linear Trend ~ 1 | 4 | 22.677 | -37.056 | 5.058 | 0.047 | HIC |
| **P** | Linear Trend ~ *Monitoring and Surveillance* | 5 | -53.675 | 118.119 | 0.000 | 0.154 | HIC |
| **P** | Linear Trend ~ Mean Temperature | 5 | -54.189 | 119.148 | 1.029 | 0.092 | HIC |
| **P** | Linear Trend ~ Infection + *Monitoring and Surveillance* | 6 | -53.037 | 119.166 | 1.047 | 0.091 | HIC |
| **P** | Linear Trend ~ Infection + Workforce | 6 | -53.714 | 120.518 | 2.399 | 0.046 | HIC |
| **P** | Linear Trend ~ Workforce | 5 | -54.975 | 120.718 | 2.599 | 0.042 | HIC |
| **P** | Linear Trend ~ 1 | 4 | -57.569 | 123.645 | 5.526 | 0.010 | HIC |
| **S** | Linear Trend ~ Mean Temperature + Workforce + Baseline | 7 | -48.134 | 112.601 | 0.000 | 0.133 | HIC |
| **S** | Linear Trend ~ Infection + Baseline | 6 | -49.926 | 113.567 | 0.967 | 0.082 | HIC |
| **S** | Linear Trend ~ Baseline | 5 | -51.259 | 113.718 | 1.117 | 0.076 | HIC |
| **S** | Linear Trend ~ Infection + *Monitoring and Surveillance* + Baseline | 7 | -49.056 | 114.445 | 1.844 | 0.053 | HIC |
| **S** | Linear Trend ~ Mean Temperature + Baseline | 6 | -50.514 | 114.742 | 2.141 | 0.046 | HIC |
| **S** | Linear Trend ~ 1 | 4 | -53.550 | 115.884 | 3.284 | 0.026 | HIC |
| **E** | Linear Trend ~ Mean Temperature + *Action* + Workforce | 5 | 11.959 | -7.919 | 0.000 | 0.650 | HIC |
| **E** | Linear Trend ~ Mean Temperature + *Action* + Baseline | 5 | 9.132 | -2.265 | 5.654 | 0.038 | HIC |
| **E** | Linear Trend ~ Mean Temperature + *Action* | 4 | 6.721 | -1.806 | 6.113 | 0.031 | HIC |
| **E** | Linear Trend ~ *General* + Mean Temperature + Workforce | 5 | 8.753 | -1.506 | 6.413 | 0.026 | HIC |
| **E** | Linear Trend ~ Mean Temperature + *Monitoring and Surveillance* + Workforce | 5 | 8.440 | -0.880 | 7.039 | 0.019 | HIC |
| **E** | Linear Trend ~ 1 | 2 | -1.235 | 7.394 | 15.312 | 0.000 | HIC |

| Model Name | Variables | df | logLik | AICc | delta | weight | income |
|---|---|---|---|---|---|---|---|
| **DPS** | Linear Trend ~ DPSE * Workforce | 8 | -16.709 | 51.475 | 0.000 | 0.207 | HIC |
| **DPS** | Linear Trend ~ DPSE | 5 | -20.646 | 52.115 | 0.640 | 0.150 | HIC |
| **DPS** | Linear Trend ~ DPSE + *Monitoring and Surveillance* | 6 | -19.923 | 53.013 | 1.538 | 0.096 | HIC |
| **DPS** | Linear Trend ~ DPSE + Infection | 6 | -20.759 | 54.685 | 3.210 | 0.042 | HIC |
| **DPS** | Linear Trend ~ DPSE + Infection + *Monitoring and Surveillance* | 7 | -19.686 | 54.950 | 3.475 | 0.036 | HIC |
| **DPS** | Linear Trend ~ 1 | 3 | -24.602 | 55.524 | 4.049 | 0.027 | HIC |
| **PSE** | Linear Trend ~ Mean Temperature | 4 | -22.094 | 52.855 | 0.000 | 0.093 | HIC |
| **PSE** | Linear Trend ~ *Monitoring and Surveillance* | 4 | -22.286 | 53.239 | 0.384 | 0.077 | HIC |
| **PSE** | Linear Trend ~ Mean Temperature + *Action* + Workforce | 6 | -19.982 | 53.412 | 0.557 | 0.071 | HIC |
| **PSE** | Linear Trend ~ *General* + Mean Temperature + Workforce | 6 | -19.995 | 53.439 | 0.584 | 0.070 | HIC |
| **PSE** | Linear Trend ~ Infection + *Monitoring and Surveillance* | 5 | -21.363 | 53.743 | 0.888 | 0.060 | HIC |
| **PSE** | Linear Trend ~ 1 | 3 | -25.166 | 56.725 | 3.870 | 0.013 | HIC |
| **DP** | Linear Trend ~ DPSE * *Monitoring and Surveillance* | 6 | 4.630 | 4.454 | 0.000 | 0.505 | HIC |
| **DP** | Linear Trend ~ DPSE + *Monitoring and Surveillance* | 5 | 2.065 | 7.070 | 2.616 | 0.137 | HIC |
| **DP** | Linear Trend ~ DPSE * Mean Temperature | 6 | 3.094 | 7.525 | 3.072 | 0.109 | HIC |
| **DP** | Linear Trend ~ DPSE * Workforce | 6 | 2.718 | 8.278 | 3.825 | 0.075 | HIC |
| **DP** | Linear Trend ~ DPSE + Infection + *Monitoring and Surveillance* | 6 | 1.806 | 10.103 | 5.649 | 0.030 | HIC |
| **DP** | Linear Trend ~ 1 | 3 | -9.326 | 25.114 | 20.660 | 0.000 | HIC |
| **PS** | Linear Trend ~ Mean Temperature | 4 | -20.747 | 50.404 | 0.000 | 0.105 | HIC |
| **PS** | Linear Trend ~ *Monitoring and Surveillance* | 4 | -20.837 | 50.583 | 0.179 | 0.096 | HIC |
| **PS** | Linear Trend ~ Infection + *Monitoring and Surveillance* | 5 | -19.859 | 51.113 | 0.709 | 0.073 | HIC |
| **PS** | Linear Trend ~ Infection + Mean Temperature | 5 | -20.441 | 52.277 | 1.873 | 0.041 | HIC |
| **PS** | Linear Trend ~ 1 | 3 | -22.953 | 52.439 | 2.035 | 0.038 | HIC |
| **PS** | Linear Trend ~ 1 | 3 | -22.953 | 52.439 | 2.035 | 0.038 | HIC |
| **SE** | Linear Trend ~ Workforce + Baseline | 5 | -15.131 | 42.198 | 0.000 | 0.054 | HIC |
| **SE** | Linear Trend ~ Mean Temperature + Workforce | 5 | -15.294 | 42.524 | 0.326 | 0.046 | HIC |
| **SE** | Linear Trend ~ 1 | 3 | -17.998 | 42.723 | 0.526 | 0.042 | HIC |
| **SE** | Linear Trend ~ DPSE * Infection | 6 | -14.010 | 42.821 | 0.623 | 0.040 | HIC |
| **SE** | Linear Trend ~ Infection + Mean Temperature + Workforce | 6 | -14.137 | 43.075 | 0.877 | 0.035 | HIC |
| **SE** | Linear Trend ~ 1 | 3 | -17.998 | 42.723 | 0.526 | 0.042 | HIC |
| **DPSEA** | Linear Trend ~ DPSE * Baseline | 9 | -6.856 | 35.312 | 0.000 | 0.289 | LMIC |
| **DPSEA** | Linear Trend ~ DPSE + Baseline | 7 | -10.080 | 36.314 | 1.002 | 0.175 | LMIC |
| **DPSEA** | Linear Trend ~ DPSE + Workforce + Baseline | 8 | -9.803 | 38.430 | 3.118 | 0.061 | LMIC |
| **DPSEA** | Linear Trend ~ Workforce + Baseline + DPSE * Sanitation | 11 | -5.594 | 38.688 | 3.376 | 0.053 | LMIC |

| Model Name | Variables | df | logLik | AICc | delta | weight | income |
|---|---|---|---|---|---|---|---|
| **DPSEA** | Linear Trend ~ Workforce + DPSE * Baseline | 10 | -7.360 | 39.210 | 3.897 | 0.041 | LMIC |
| **DPSEA** | Linear Trend ~ 1 | 3 | -39.742 | 85.913 | 50.601 | 0.000 | LMIC |
| **DPSEA.noDr** | Linear Trend ~ DPSE * Baseline | 10 | -14.246 | 51.983 | 0.000 | 0.700 | LMIC |
| **DPSEA.noDr** | Linear Trend ~ *Monitoring and Surveillance* + DPSE * Baseline | 11 | -15.219 | 56.696 | 4.713 | 0.066 | LMIC |
| **DPSEA.noDr** | Linear Trend ~ Baseline + DPSE * *Monitoring and Surveillance* | 11 | -15.566 | 57.390 | 5.407 | 0.047 | LMIC |
| **DPSEA.noDr** | Linear Trend ~ *Action* + DPSE * Baseline | 11 | -15.591 | 57.441 | 5.458 | 0.046 | LMIC |
| **DPSEA.noDr** | Linear Trend ~ *General* + DPSE * Baseline | 11 | -16.171 | 58.601 | 6.618 | 0.026 | LMIC |
| **DPSEA.noDr** | Linear Trend ~ 1 | 3 | -43.699 | 93.740 | 41.757 | 0.000 | LMIC |
| **aP.noDr** | Linear Trend ~ DPSE * Baseline | 10 | -15.008 | 53.682 | 0.000 | 0.945 | LMIC |
| **aP.noDr** | Linear Trend ~ DPSE + Baseline | 7 | -22.174 | 60.127 | 6.444 | 0.038 | LMIC |
| **aP.noDr** | Linear Trend ~ DPSE + *Action* + Baseline | 8 | -22.726 | 63.775 | 10.093 | 0.006 | LMIC |
| **aP.noDr** | Linear Trend ~ DPSE + *Monitoring and Surveillance* + Baseline | 8 | -23.341 | 65.005 | 11.323 | 0.003 | LMIC |
| **aP.noDr** | Linear Trend ~ DPSE + *General* + Baseline | 8 | -23.568 | 65.459 | 11.776 | 0.003 | LMIC |
| **aP.noDr** | Linear Trend ~ 1 | 3 | -42.820 | 91.999 | 38.317 | 0.000 | LMIC |
| **aS.noDr** | Linear Trend ~ DPSE * Baseline | 10 | -10.103 | 48.668 | 0.000 | 0.755 | LMIC |
| **aS.noDr** | Linear Trend ~ DPSE | 6 | -18.966 | 52.732 | 4.064 | 0.099 | LMIC |
| **aS.noDr** | Linear Trend ~ DPSE + Baseline | 7 | -17.745 | 53.352 | 4.684 | 0.073 | LMIC |
| **aS.noDr** | Linear Trend ~ DPSE + *General* | 7 | -19.883 | 57.628 | 8.960 | 0.009 | LMIC |
| **aS.noDr** | Linear Trend ~ *Awareness and Education* + DPSE | 7 | -19.915 | 57.692 | 9.024 | 0.008 | LMIC |
| **aS.noDr** | Linear Trend ~ 1 | 3 | -26.605 | 59.937 | 11.269 | 0.003 | LMIC |
| **aE.noDr** | Linear Trend ~ DPSE | 6 | -16.354 | 48.526 | 0.000 | 0.361 | LMIC |
| **aE.noDr** | Linear Trend ~ 1 | 3 | -21.386 | 49.733 | 1.206 | 0.198 | LMIC |
| **aE.noDr** | Linear Trend ~ DPSE + Baseline | 7 | -16.324 | 51.980 | 3.454 | 0.064 | LMIC |
| **aE.noDr** | Linear Trend ~ Baseline | 4 | -21.484 | 52.635 | 4.109 | 0.046 | LMIC |
| **aE.noDr** | Linear Trend ~ DPSE + *Monitoring and Surveillance* | 7 | -17.137 | 53.608 | 5.082 | 0.028 | LMIC |
| **aE.noDr** | Linear Trend ~ 1 | 3 | -21.386 | 49.733 | 1.206 | 0.198 | LMIC |
| **aP** | Linear Trend ~ DPSE * Baseline | 9 | -7.245 | 36.163 | 0.000 | 0.450 | LMIC |
| **aP** | Linear Trend ~ DPSE + Baseline | 7 | -10.325 | 36.846 | 0.683 | 0.320 | LMIC |
| **aP** | Linear Trend ~ DPSE + Workforce + Baseline | 8 | -10.135 | 39.149 | 2.987 | 0.101 | LMIC |
| **aP** | Linear Trend ~ DPSE + *Action* + Baseline | 8 | -11.538 | 41.956 | 5.794 | 0.025 | LMIC |
| **aP** | Linear Trend ~ DPSE + *General* + Baseline | 8 | -11.686 | 42.252 | 6.089 | 0.021 | LMIC |
| **aP** | Linear Trend ~ 1 | 3 | -39.203 | 84.842 | 48.679 | 0.000 | LMIC |
| **aS** | Linear Trend ~ DPSE * Baseline | 8 | 2.740 | 18.990 | 0.000 | 0.329 | LMIC |
| **aS** | Linear Trend ~ DPSE + Baseline | 6 | -2.180 | 20.781 | 1.792 | 0.134 | LMIC |
| **aS** | Linear Trend ~ DPSE + Workforce + Baseline | 7 | -0.626 | 21.474 | 2.484 | 0.095 | LMIC |
| **aS** | Linear Trend ~ DPSE + *General* + Baseline | 7 | -0.720 | 21.662 | 2.672 | 0.087 | LMIC |
| **aS** | Linear Trend ~ DPSE + *Action* + Baseline | 7 | -1.080 | 22.383 | 3.393 | 0.060 | LMIC |

| Model Name | Variables | df | logLik | AICc | delta | weight | income |
|---|---|---|---|---|---|---|---|
| **aS** | Linear Trend ~ 1 | 2 | -21.149 | 46.819 | 27.829 | 0.000 | LMIC |
| **aE** | Linear Trend ~ DPSE + Baseline | 6 | -1.744 | 21.950 | 0.000 | 0.231 | LMIC |
| **aE** | Linear Trend ~ DPSE + Workforce + Baseline | 7 | 0.434 | 22.465 | 0.514 | 0.178 | LMIC |
| **aE** | Linear Trend ~ *Awareness and Education* + DPSE + Baseline | 7 | 0.130 | 23.074 | 1.124 | 0.132 | LMIC |
| **aE** | Linear Trend ~ DPSE * Baseline | 7 | -0.104 | 23.542 | 1.591 | 0.104 | LMIC |
| **aE** | Linear Trend ~ DPSE + Animal Production + Baseline | 7 | -0.475 | 24.282 | 2.332 | 0.072 | LMIC |
| **aE** | Linear Trend ~ 1 | 2 | -16.632 | 37.969 | 16.019 | 0.000 | LMIC |
| **Dr** | Linear Trend ~ 1 | 4 | -28.779 | 65.866 | 0.000 | 0.547 | LMIC |
| **Dr** | Linear Trend ~ Baseline | 5 | -28.884 | 68.233 | 2.367 | 0.168 | LMIC |
| **Dr** | Linear Trend ~ *General* | 5 | -29.748 | 69.961 | 4.095 | 0.071 | LMIC |
| **Dr** | Linear Trend ~ *Action* | 5 | -30.311 | 71.088 | 5.222 | 0.040 | LMIC |
| **Dr** | Linear Trend ~ *Awareness and Education* | 5 | -30.875 | 72.216 | 6.350 | 0.023 | LMIC |
| **Dr** | Linear Trend ~ 1 | 4 | -28.779 | 65.866 | 0.000 | 0.547 | LMIC |
| **P** | Linear Trend ~ 1 | 4 | -66.595 | 141.710 | 0.000 | 0.342 | LMIC |
| **P** | Linear Trend ~ Sanitation | 5 | -66.919 | 144.627 | 2.917 | 0.080 | LMIC |
| **P** | Linear Trend ~ *Awareness and Education* | 5 | -67.456 | 145.701 | 3.990 | 0.047 | LMIC |
| **P** | Linear Trend ~ *General* | 5 | -67.559 | 145.908 | 4.198 | 0.042 | LMIC |
| **P** | Linear Trend ~ *Action* | 5 | -67.564 | 145.918 | 4.208 | 0.042 | LMIC |
| **P** | Linear Trend ~ 1 | 4 | -66.595 | 141.710 | 0.000 | 0.342 | LMIC |
| **DPS** | Linear Trend ~ DPSE * Baseline | 8 | -6.856 | 32.592 | 0.000 | 0.503 | LMIC |
| **DPS** | Linear Trend ~ DPSE + Baseline | 6 | -10.080 | 33.776 | 1.184 | 0.278 | LMIC |
| **DPS** | Linear Trend ~ DPSE + Workforce + Baseline | 7 | -9.803 | 35.803 | 3.210 | 0.101 | LMIC |
| **DPS** | Linear Trend ~ DPSE + *Action* + Baseline | 7 | -11.222 | 38.640 | 6.048 | 0.024 | LMIC |
| **DPS** | Linear Trend ~ DPSE + *General* + Baseline | 7 | -11.380 | 38.956 | 6.364 | 0.021 | LMIC |
| **DPS** | Linear Trend ~ 1 | 3 | -37.062 | 80.560 | 47.968 | 0.000 | LMIC |
| **PSE** | Linear Trend ~ DPSE + Baseline | 6 | -11.667 | 38.834 | 0.000 | 0.533 | LMIC |
| **PSE** | Linear Trend ~ DPSE * Baseline | 7 | -11.137 | 41.143 | 2.309 | 0.168 | LMIC |
| **PSE** | Linear Trend ~ DPSE + Sanitation + Baseline | 7 | -12.188 | 43.246 | 4.413 | 0.059 | LMIC |
| **PSE** | Linear Trend ~ *Awareness and Education* + DPSE + Baseline | 7 | -12.506 | 43.881 | 5.047 | 0.043 | LMIC |
| **PSE** | Linear Trend ~ DPSE + Workforce + Baseline | 7 | -12.756 | 44.381 | 5.547 | 0.033 | LMIC |
| **PSE** | Linear Trend ~ 1 | 3 | -21.942 | 50.774 | 11.940 | 0.001 | LMIC |
| **DP** | Linear Trend ~ DPSE * Baseline | 6 | -7.860 | 29.434 | 0.000 | 0.337 | LMIC |
| **DP** | Linear Trend ~ DPSE + Baseline | 5 | -9.182 | 29.565 | 0.130 | 0.316 | LMIC |
| **DP** | Linear Trend ~ DPSE + Workforce + Baseline | 6 | -8.309 | 30.331 | 0.897 | 0.215 | LMIC |
| **DP** | Linear Trend ~ DPSE + *Action* + Baseline | 6 | -10.561 | 34.836 | 5.402 | 0.023 | LMIC |
| **DP** | Linear Trend ~ DPSE + Sanitation + Baseline | 6 | -10.593 | 34.900 | 5.466 | 0.022 | LMIC |
| **DP** | Linear Trend ~ 1 | 3 | -35.811 | 78.084 | 48.650 | 0.000 | LMIC |
| **PS** | Linear Trend ~ Baseline | 4 | -9.237 | 28.075 | 0.000 | 0.518 | LMIC |

| **Model Name** | **Variables** | **df** | **logLik** | **AICc** | **delta** | **weight** | **income** |
|---|---|---|---|---|---|---|---|
| **PS** | Linear Trend ~ Sanitation + Baseline | 5 | -9.767 | 32.034 | 3.959 | 0.072 | LMIC |
| **PS** | Linear Trend ~ *Awareness and Education* + Baseline | 5 | -10.081 | 32.662 | 4.587 | 0.052 | LMIC |
| **PS** | Linear Trend ~ Workforce + Baseline | 5 | -10.307 | 33.115 | 5.040 | 0.042 | LMIC |
| **PS** | Linear Trend ~ Infection + Baseline | 5 | -10.347 | 33.194 | 5.119 | 0.040 | LMIC |
| **PS** | Linear Trend ~ 1 | 3 | -17.831 | 42.585 | 14.511 | 0.000 | LMIC |
| **DPSEA** | Categorical Trend ~ DPSE * Gini + DPSE * Mean Temperature | 13 | -33.291 | 95.163 | 0.000 | 0.602 | ALL |
| **DPSEA** | Categorical Trend ~ DPSE * Mean Temperature + DPSE * Workforce | 13 | -36.107 | 100.795 | 5.633 | 0.036 | ALL |
| **DPSEA** | Categorical Trend ~ Workforce + DPSE * Mean Temperature | 10 | -40.197 | 101.921 | 6.759 | 0.021 | ALL |
| **DPSEA** | Categorical Trend ~ *General* + Workforce + DPSE * Mean Temperature | 11 | -39.562 | 102.969 | 7.807 | 0.012 | ALL |
| **DPSEA** | Categorical Trend ~ GDP + Workforce + DPSE * Mean Temperature | 11 | -39.655 | 103.155 | 7.993 | 0.011 | ALL |
| **DPSEA** | Categorical Trend ~ 1 | 2 | -107.357 | 218.793 | 123.631 | 0.000 | ALL |
| **DPSEA.noDr** | Categorical Trend ~ Animal Production + *Action* + DPSE * Mean Temperature | 11 | -56.943 | 137.328 | 0.000 | 0.514 | ALL |
| **DPSEA.noDr** | Categorical Trend ~ *General* + Animal Production + DPSE * Mean Temperature | 11 | -57.691 | 138.824 | 1.496 | 0.243 | ALL |
| **DPSEA.noDr** | Categorical Trend ~ *Awareness and Education* + Animal Production + DPSE * Mean Temperature | 11 | -59.994 | 143.431 | 6.103 | 0.024 | ALL |
| **DPSEA.noDr** | Categorical Trend ~ *General* + DPSE * Mean Temperature | 10 | -61.421 | 144.037 | 6.709 | 0.018 | ALL |
| **DPSEA.noDr** | Categorical Trend ~ *General* + Baseline + DPSE * Mean Temperature | 11 | -60.511 | 144.465 | 7.137 | 0.014 | ALL |
| **DPSEA.noDr** | Categorical Trend ~ 1 | 2 | -135.161 | 274.385 | 137.057 | 0.000 | ALL |
| **aP.noDr** | Categorical Trend ~ DPSE + *General* + Animal Production | 7 | -64.701 | 144.032 | 0.000 | 0.218 | ALL |
| **aP.noDr** | Categorical Trend ~ DPSE * Mean Temperature | 9 | -62.586 | 144.195 | 0.163 | 0.200 | ALL |
| **aP.noDr** | Categorical Trend ~ DPSE * *Monitoring and Surveillance* | 9 | -63.573 | 146.169 | 2.137 | 0.075 | ALL |
| **aP.noDr** | Categorical Trend ~ DPSE + *General* | 6 | -66.951 | 146.371 | 2.339 | 0.068 | ALL |
| **aP.noDr** | Categorical Trend ~ DPSE + Animal Production + *Action* | 7 | -66.036 | 146.701 | 2.669 | 0.057 | ALL |
| **aP.noDr** | Categorical Trend ~ 1 | 2 | -128.882 | 261.830 | 117.798 | 0.000 | ALL |
| **aS.noDr** | Categorical Trend ~ DPSE + *General* + Animal Production | 7 | -51.992 | 118.796 | 0.000 | 0.397 | ALL |

| Model Name | Variables | df | logLik | AICc | delta | weight | income |
|---|---|---|---|---|---|---|---|
| **aS.noDr** | Categorical Trend ~ DPSE + Animal Production + *Action* | 7 | -52.756 | 120.324 | 1.528 | 0.185 | ALL |
| **aS.noDr** | Categorical Trend ~ *Awareness and Education* + DPSE + Animal Production | 7 | -53.625 | 122.061 | 3.265 | 0.078 | ALL |
| **aS.noDr** | Categorical Trend ~ DPSE + *General* | 6 | -55.090 | 122.784 | 3.988 | 0.054 | ALL |
| **aS.noDr** | Categorical Trend ~ *Awareness and Education* + DPSE + *General* | 7 | -54.748 | 124.308 | 5.512 | 0.025 | ALL |
| **aS.noDr** | Categorical Trend ~ 1 | 2 | -100.980 | 206.044 | 87.248 | 0.000 | ALL |
| **aE.noDr** | Categorical Trend ~ DPSE + *General* + Animal Production | 7 | -48.671 | 112.246 | 0.000 | 0.292 | ALL |
| **aE.noDr** | Categorical Trend ~ DPSE + Animal Production + *Action* | 7 | -49.031 | 112.965 | 0.719 | 0.204 | ALL |
| **aE.noDr** | Categorical Trend ~ *Awareness and Education* + DPSE + Animal Production | 7 | -49.630 | 114.163 | 1.917 | 0.112 | ALL |
| **aE.noDr** | Categorical Trend ~ DPSE + *General* | 6 | -51.549 | 115.769 | 3.523 | 0.050 | ALL |
| **aE.noDr** | Categorical Trend ~ *Awareness and Education* + DPSE + *General* | 7 | -51.208 | 117.319 | 5.073 | 0.023 | ALL |
| **aE.noDr** | Categorical Trend ~ 1 | 2 | -91.080 | 186.253 | 74.007 | 0.000 | ALL |
| **aP** | Categorical Trend ~ DPSE * Mean Temperature | 9 | -34.041 | 87.367 | 0.000 | 0.997 | ALL |
| **aP** | Categorical Trend ~ DPSE * Sanitation | 9 | -40.224 | 99.733 | 12.366 | 0.002 | ALL |
| **aP** | Categorical Trend ~ DPSE * Workforce | 9 | -41.540 | 102.366 | 14.998 | 0.001 | ALL |
| **aP** | Categorical Trend ~ DPSE * Gini | 9 | -41.746 | 102.778 | 15.410 | 0.000 | ALL |
| **aP** | Categorical Trend ~ DPSE * Baseline | 9 | -42.375 | 104.037 | 16.670 | 0.000 | ALL |
| **aP** | Categorical Trend ~ 1 | 2 | -103.852 | 211.786 | 124.419 | 0.000 | ALL |
| **aS** | Categorical Trend ~ DPSE + *Action* + Workforce | 7 | -36.487 | 88.083 | 0.000 | 0.219 | ALL |
| **aS** | Categorical Trend ~ DPSE + *General* + Workforce | 7 | -36.683 | 88.475 | 0.393 | 0.180 | ALL |
| **aS** | Categorical Trend ~ DPSE + *General* + Animal Production | 7 | -37.003 | 89.115 | 1.032 | 0.131 | ALL |
| **aS** | Categorical Trend ~ DPSE + Animal Production + *Action* | 7 | -38.329 | 91.767 | 3.685 | 0.035 | ALL |
| **aS** | Categorical Trend ~ DPSE + *General* | 6 | -39.615 | 92.053 | 3.971 | 0.030 | ALL |
| **aS** | Categorical Trend ~ 1 | 2 | -74.774 | 153.660 | 65.578 | 0.000 | ALL |
| **aE** | Categorical Trend ~ DPSE * Mean Temperature | 9 | -24.729 | 69.480 | 0.000 | 0.989 | ALL |
| **aE** | Categorical Trend ~ DPSE * Sanitation | 9 | -30.653 | 81.329 | 11.849 | 0.003 | ALL |
| **aE** | Categorical Trend ~ DPSE + *Action* + Workforce | 7 | -33.254 | 81.738 | 12.259 | 0.002 | ALL |
| **aE** | Categorical Trend ~ DPSE + *General* + Workforce | 7 | -34.057 | 83.344 | 13.865 | 0.001 | ALL |

| Model Name | Variables | df | logLik | AICc | delta | weight | income |
|---|---|---|---|---|---|---|---|
| **aE** | Categorical Trend ~ DPSE + *General* + Animal Production | 7 | -34.167 | 83.565 | 14.085 | 0.001 | ALL |
| **aE** | Categorical Trend ~ 1 | 2 | -67.705 | 139.535 | 70.055 | 0.000 | ALL |
| **Dr** | Categorical Trend ~ GDP + Baseline | 4 | -132.757 | 273.663 | 0.000 | 0.065 | ALL |
| **Dr** | Categorical Trend ~ GDP + Mean Temperature + Baseline | 5 | -132.004 | 274.233 | 0.570 | 0.049 | ALL |
| **Dr** | Categorical Trend ~ *General* * income | 5 | -132.019 | 274.262 | 0.599 | 0.048 | ALL |
| **Dr** | Categorical Trend ~ GDP | 3 | -134.323 | 274.736 | 1.073 | 0.038 | ALL |
| **Dr** | Categorical Trend ~ GDP + Mean Temperature | 4 | -133.386 | 274.921 | 1.258 | 0.034 | ALL |
| **Dr** | Categorical Trend ~ 1 | 2 | -142.735 | 289.515 | 15.852 | 0.000 | ALL |
| **P** | Categorical Trend ~ *Monitoring and Surveillance* + Baseline | 5 | -70.903 | 152.182 | 0.000 | 0.081 | ALL |
| **P** | Categorical Trend ~ Infection + *Monitoring and Surveillance* + Baseline | 6 | -70.199 | 152.926 | 0.745 | 0.056 | ALL |
| **P** | Categorical Trend ~ *Action* + Baseline | 5 | -71.307 | 152.990 | 0.808 | 0.054 | ALL |
| **P** | Categorical Trend ~ Mean Temperature + *Monitoring and Surveillance* + Baseline | 6 | -70.423 | 153.375 | 1.193 | 0.045 | ALL |
| **P** | Categorical Trend ~ *Monitoring and Surveillance* + Workforce + Baseline | 6 | -70.578 | 153.684 | 1.503 | 0.038 | ALL |
| **P** | Categorical Trend ~ 1 | 3 | -83.958 | 174.063 | 21.882 | 0.000 | ALL |
| **S** | Categorical Trend ~ GDP + *Monitoring and Surveillance* + Baseline | 6 | -27.018 | 67.621 | 0.000 | 0.087 | ALL |
| **S** | Categorical Trend ~ *Monitoring and Surveillance* + Baseline | 5 | -28.903 | 68.916 | 1.295 | 0.046 | ALL |
| **S** | Categorical Trend ~ Infection + *Monitoring and Surveillance* + Baseline | 6 | -27.896 | 69.377 | 1.756 | 0.036 | ALL |
| **S** | Categorical Trend ~ *Monitoring and Surveillance* + Vaccination + Baseline | 6 | -27.916 | 69.417 | 1.796 | 0.035 | ALL |
| **S** | Categorical Trend ~ *Awareness and Education* + Mean Temperature + Baseline | 6 | -28.090 | 69.764 | 2.143 | 0.030 | ALL |
| **S** | Categorical Trend ~ 1 | 3 | -41.555 | 89.538 | 21.917 | 0.000 | ALL |
| **E** | Categorical Trend ~ *Awareness and Education* + Sanitation | 4 | -1.910 | 15.152 | 0.000 | 0.033 | ALL |
| **E** | Categorical Trend ~ *Awareness and Education* + income | 4 | -1.910 | 15.152 | 0.000 | 0.033 | ALL |
| **E** | Categorical Trend ~ *Awareness and Education* * income | 4 | -1.910 | 15.152 | 0.000 | 0.033 | ALL |
| **E** | Categorical Trend ~ *Awareness and Education* + income + Population Density | 5 | 0.000 | 15.455 | 0.302 | 0.029 | ALL |

| Model Name | Variables | df | logLik | AICc | delta | weight | income |
|---|---|---|---|---|---|---|---|
| **E** | Categorical Trend ~ *Awareness and Education* + income + Animal Production | 5 | 0.000 | 15.455 | 0.302 | 0.029 | ALL |
| **E** | Categorical Trend ~ 1 | 2 | -7.922 | 20.701 | 5.549 | 0.002 | ALL |
| **DPS** | Categorical Trend ~ DPSE * Mean Temperature | 7 | -38.065 | 90.992 | 0.000 | 0.097 | ALL |
| **DPS** | Categorical Trend ~ DPSE | 4 | -42.536 | 93.373 | 2.381 | 0.030 | ALL |
| **DPS** | Categorical Trend ~ DPSE + *General* | 5 | -41.507 | 93.470 | 2.478 | 0.028 | ALL |
| **DPS** | Categorical Trend ~ DPSE + *Action* | 5 | -41.651 | 93.758 | 2.766 | 0.024 | ALL |
| **DPS** | Categorical Trend ~ *Awareness and Education* + DPSE | 5 | -41.711 | 93.877 | 2.885 | 0.023 | ALL |
| **DPS** | Categorical Trend ~ 1 | 2 | -95.524 | 195.137 | 104.145 | 0.000 | ALL |
| **PSE** | Categorical Trend ~ DPSE * Sanitation | 7 | -25.576 | 66.424 | 0.000 | 0.686 | ALL |
| **PSE** | Categorical Trend ~ DPSE * Workforce | 7 | -26.711 | 68.695 | 2.271 | 0.220 | ALL |
| **PSE** | Categorical Trend ~ DPSE * Gini | 7 | -27.928 | 71.130 | 4.706 | 0.065 | ALL |
| **PSE** | Categorical Trend ~ DPSE * Mean Temperature | 7 | -28.946 | 73.165 | 6.741 | 0.024 | ALL |
| **PSE** | Categorical Trend ~ DPSE * Baseline | 7 | -31.982 | 79.237 | 12.813 | 0.001 | ALL |
| **PSE** | Categorical Trend ~ 1 | 2 | -49.352 | 102.832 | 36.408 | 0.000 | ALL |
| **DP** | Categorical Trend ~ DPSE | 3 | -18.732 | 43.685 | 0.000 | 0.055 | ALL |
| **DP** | Categorical Trend ~ DPSE + *General* | 4 | -18.561 | 45.497 | 1.811 | 0.022 | ALL |
| **DP** | Categorical Trend ~ *Awareness and Education* + DPSE | 4 | -18.617 | 45.609 | 1.923 | 0.021 | ALL |
| **DP** | Categorical Trend ~ DPSE + *Action* | 4 | -18.618 | 45.609 | 1.924 | 0.021 | ALL |
| **DP** | Categorical Trend ~ DPSE + Workforce | 4 | -18.635 | 45.644 | 1.958 | 0.021 | ALL |
| **DP** | Categorical Trend ~ 1 | 2 | -77.561 | 159.232 | 115.547 | 0.000 | ALL |
| **PS** | Categorical Trend ~ DPSE + *Monitoring and Surveillance* + Workforce | 5 | -22.292 | 55.405 | 0.000 | 0.067 | ALL |
| **PS** | Categorical Trend ~ DPSE + *Action* + Workforce | 5 | -22.761 | 56.344 | 0.939 | 0.042 | ALL |
| **PS** | Categorical Trend ~ DPSE + Animal Production + *Action* | 5 | -22.779 | 56.380 | 0.974 | 0.041 | ALL |
| **PS** | Categorical Trend ~ DPSE | 3 | -25.194 | 56.708 | 1.303 | 0.035 | ALL |
| **PS** | Categorical Trend ~ DPSE * *General* | 5 | -23.232 | 57.286 | 1.881 | 0.026 | ALL |
| **PS** | Categorical Trend ~ 1 | 2 | -42.092 | 88.341 | 32.936 | 0.000 | ALL |
| **SE** | Categorical Trend ~ Mean Temperature + *Action* + Workforce | 5 | -16.861 | 45.487 | 0.000 | 0.098 | ALL |
| **SE** | Categorical Trend ~ *Action* + Workforce + Baseline | 5 | -17.204 | 46.172 | 0.685 | 0.070 | ALL |
| **SE** | Categorical Trend ~ *General* + Workforce + Baseline | 5 | -17.208 | 46.180 | 0.693 | 0.070 | ALL |
| **SE** | Categorical Trend ~ *General* + Mean Temperature + Workforce | 5 | -17.356 | 46.476 | 0.989 | 0.060 | ALL |

| Model Name | Variables | df | logLik | AICc | delta | weight | income |
|---|---|---|---|---|---|---|---|
| **SE** | Categorical Trend ~ DPSE + *Action* + Workforce | 5 | -17.457 | 46.678 | 1.191 | 0.054 | ALL |
| **SE** | Categorical Trend ~ 1 | 2 | -25.898 | 56.120 | 10.633 | 0.000 | ALL |
